# Associations between IL-6 and trajectories of depressive symptoms across the life course: Evidence from ALSPAC and UK Biobank cohorts

**DOI:** 10.1101/2024.04.26.24306425

**Authors:** A. J. Edmondson-Stait, E. Davyson, X. Shen, M. J. Adams, G. M. Khandaker, V. E. Miron, A. M. McIntosh, S. M. Lawrie, A. S. F. Kwong, H. C. Whalley

**Affiliations:** Translational Neuroscience PhD Programme, Centre for Clinical Brain Sciences, University of Edinburgh,UK.; Centre for Clinical Brain Sciences, University of Edinburgh, UK; MRC Integrative Epidemiology Unit, Population Health Sciences, Bristol Medical School, University of Bristol, UK; Centre for Academic Mental Health, Population Health Sciences, Bristol Medical School, University of Bristol, UK; NIHR Bristol Biomedical Research Centre, Bristol, UK; BARLO Multiple Sclerosis Centre, Keenan Research Centre for Biomedical Science at St. Michael’s Hospital, Toronto M5B 1T8, Canada; Department of Immunology, University of Toronto, Toronto M5S 1A8, Canada; UK Dementia Research Institute at the University of Edinburgh, Edinburgh BioQuarter, Edinburgh EH16 4TJ, UK; Generation Scotland, Institute of Genetics and Cancer, University of Edinburgh, UK

## Abstract

Peripheral inflammatory markers, including serum IL-6, are associated with depression, but less is known about how these markers associate with depression at different stages of the life-course. We examined associations between serum IL-6 levels at baseline and subsequent depression symptom trajectories in two longitudinal cohorts: ALSPAC (age 10-28y;N=4,835) and UK Biobank (39- 86y;N=39,613) using multi-level growth curve modelling. Models were adjusted for sex, BMI and socioeconomic factors. Depressive symptoms were measured using the Short Moods and Feelings Questionnaire (SMFQ) in ALSPAC (max timepoints=11) and the Patient Health Questionnaire-2 (PHQ-2) in UK Biobank (max timepoints=8). Higher baseline IL-6 was associated with worse depression symptom trajectories in both cohorts (largest effect size: 0.046 (ALSPAC, age 16y)). These associations were stronger in the younger ALSPAC cohort, where additionally higher IL-6 at age 9 years was associated with worse depression symptoms trajectories in females compared to males. Weaker sex differences were observed in the older cohort, UK Biobank. These findings suggest that systemic inflammation may influence the severity and course of depressive symptoms across the life course, which is apparent regardless of age and differences in measures and number of time points between these large, population-based cohorts.

## 2. Introduction

There is substantial evidence to suggest low grade inflammation, as reflected by elevated levels of circulating inflammatory markers in the blood and cerebrospinal fluid (CSF), may contribute to the aetiology of depression ^1–4^. Neuroimaging and post-mortem brain studies have shown increased markers of neuroinflammation in individuals with depression compared to controls ^4–6^. Longitudinal studies have shown increased blood IL-6 levels in childhood associate with depressive and psychotic symptoms in early adulthood ^7–9^. Increased inflammatory markers have been also shown to associate with worse depressive symptom severity, including in a Mendelian randomisation (MR) study that found a potential causal association of IL-6 with suicidal thoughts ^10, 11^. Causal evidence also comes from RCTs showing anti-inflammatory treatment for chronic inflammatory conditions improves depressive symptoms independent of improvement in physical symptoms and other MR studies suggesting putative causality of IL-6 on major depressive disorder ^12, 13^.

Depression affects individuals across the entire life course with an onset typically occurring between ages 20 to 30 years ^14, 15^. However, there are few studies investigating the effect of baseline inflammation on the longitudinal patterns of depressive symptoms over the life course. A study using latent class analysis in the ALSPAC cohort showed that serum IL-6 levels at age 9 years associated with a trajectory group of persistently worse depressive symptoms from ages 10 to 19 years ^16^. A study also using latent class analysis in the NESDA cohort (age 18 to 65 years at baseline) found increased inflammatory blood markers associated with an atypical depression subgroup ^17^. Over a 6-year follow-up this subgroup had higher BMI and rate of metabolic syndrome compared to a melancholic depression subgroup and controls ^18^. However, none of these studies examine the effects of inflammation over a larger period of the life course.

Examining the effects of inflammation on depression across the life course provides insight into the heterogeneity and underlying mechanisms of depression at specific developmental stages, aiding the development of biologically based stratification.

Depression is highly heterogeneous and there is increasing cross-sectional evidence that an inflammatory subgroup of depression exists, associated with worse depressive symptom severity ^10, 19^. Therefore, it is crucial to examine whether increased inflammation associates with increased depression symptom severity over different stages of the life course. One such method for understanding the longitudinal relationships between inflammation and depression is trajectory analysis ^20^. Briefly, this method assesses the patterns of change in depressive symptoms over time (trajectories) for individuals or groups of individuals from repeated assessments of depression symptoms ^20^. This then facilitates the investigation into risk factors that may influence the course of these trajectories and whether these effects persist over time.

Additionally, it is known there is a sex difference in both depression and inflammation ^20–23^. Evidence suggests these sex differences extend into differences in inflammatory-associated depression, especially in adolescence but with inconsistent findings in later life ^24–27^. There is a need to understand the sex differences more fully at different stages of the life course, whilst assessing repeated measures of subsequent depression.

Here, we used multi-level growth curve modelling to investigate the effects of IL-6 on subsequent trajectories of depressive symptoms in two longitudinal cohorts: ALSPAC (age 10-28y) and UK Biobank (39-86y). Specifically, we tested whether increased baseline measures of serum IL-6 are associated with worse trajectories of depressive symptoms and if this effect is seen consistently across two different cohorts spanning early and later life. Individuals were divided into groups based on IL-6 tertiles, with the bottom third tertile group consisting of people with lower levels of IL-6 and the top third tertile group consisting of people with higher levels of IL-6, as has been studied previously ^28, 29^. We also tested if there was a sex difference in the relationship between IL-6 and subsequent trajectories of depressive symptoms, by stratifying analysis by sex. Finally, due to the difficulty in interpreting the effects of polynomial trajectory models, we also calculated the mean depressive scores for each IL-6 tertile trajectory and assessed the differences in depressive scores between the top and bottom third IL-6 tertile trajectories of depressive symptoms, at different ages.

## 3. Materials and methods

### 3.1 Study Sample

#### 3.1.1 ALSPAC Cohort

ALSPAC is an ongoing, longitudinal, prospective, population-based study in South- West England investigating the impact of various exposures on health and developmental outcomes ^30–32^. Initially 14,541 pregnant mothers with an estimated delivery date between April 1991 and December 1992 were recruited. This resulted in 14,092 live births and 13,988 children still alive after one year. When the oldest children were approximately 7 years of age, an attempt was made to bolster the initial sample with eligible cases who had failed to join the study originally. The total sample size for analyses using any data collected after the age of seven is therefore 15,447 pregnancies, of which 14,901 children were alive at 1 year of age. Further details of this study cohort are described in the cohort profile publications ^30–32^.

Demographics of ALSPAC participants used within the current study are shown in Table 1.

**Table 1.** Demographic table of ALSPAC participants.

|  | IL-6 Tertile Group |  |  |
| --- | --- | --- | --- |
|  | Bottom Third | Middle Third | Top Third |
| n | 1616 | 1602 | 1617 |
| Sex = Males (%) | 943 ( 58.4) | 807 ( 50.4) | 666 ( 41.2) |
| BMI (log transformed) (mean (SD)) | 2.81 (0.11) | 2.85 (0.14) | 2.90 (0.17) |
| Maternal education at birth (%) |  |  |  |
| A-level/Degree | 710 ( 43.9) | 646 ( 40.3) | 636 ( 39.3) |
| CSE/O-level/Vocational | 753 ( 46.6) | 789 ( 49.3) | 780 ( 48.2) |
| NA | 153 ( 9.5) | 167 ( 10.4) | 201 ( 12.4) |
| Currently taking any medication (%) |  |  |  |
| FALSE | 1426 ( 88.2) | 1373 ( 85.7) | 1341 ( 82.9) |
| TRUE | 187 ( 11.6) | 227 ( 14.2) | 272 ( 16.8) |
| NA | 3 ( 0.2) | 2 ( 0.1) | 4 ( 0.2) |
| Townsend Deprivation Index quintile (%) |  |  |  |
| 1 | 478 ( 29.6) | 399 ( 24.9) | 399 ( 24.7) |
| 2 | 219 ( 13.6) | 207 ( 12.9) | 198 ( 12.2) |
| 3 | 245 ( 15.2) | 219 ( 13.7) | 235 ( 14.5) |
| 4 | 222 ( 13.7) | 261 ( 16.3) | 271 ( 16.8) |
| 5 | 74 ( 4.6) | 93 ( 5.8) | 95 ( 5.9) |
| NA | 378 ( 23.4) | 423 ( 26.4) | 419 ( 25.9) |
| IL-6 (Inverse Normal Transformed, Z-score) (mean (SD)) | -1.09 (0.53) | 0.00 (0.25) | 1.09 (0.53) |
| IL-6 (raw value, pg/ml) | 0.39 (0.13) | 0.82 (0.15) | 2.48 (1.95) |
| SMFQ (Time Point 1) (mean (SD)) | 3.87 (3.32) | 3.96 (3.50) | 4.10 (3.55) |
| SMFQ (Time Point 2) (mean (SD)) | 3.79 (3.75) | 3.71 (3.56) | 4.26 (4.04) |
| SMFQ (Time Point 3) (mean (SD)) | 4.46 (4.02) | 4.73 (4.42) | 5.38 (4.74) |
| SMFQ (Time Point 4) (mean (SD)) | 5.38 (5.23) | 5.54 (5.43) | 6.32 (5.89) |
| SMFQ (Time Point 5) (mean (SD)) | 5.99 (4.90) | 6.56 (5.41) | 6.87 (5.40) |
| SMFQ (Time Point 6) (mean (SD)) | 6.12 (5.39) | 6.19 (5.54) | 7.36 (6.21) |
| SMFQ (Time Point 7) (mean (SD)) | 5.09 (4.78) | 5.26 (5.26) | 5.68 (5.49) |
| SMFQ (Time Point 8) (mean (SD)) | 5.76 (5.13) | 5.79 (5.25) | 6.25 (5.58) |
| SMFQ (Time Point 9) (mean (SD)) | 6.51 (5.74) | 6.27 (5.57) | 6.99 (6.01) |
| SMFQ (Time Point 10) (mean (SD)) | 6.19 (5.97) | 6.24 (5.97) | 7.12 (6.60) |
| SMFQ (Time Point 11) (mean (SD)) | 6.43 (5.89) | 6.01 (5.72) | 7.01 (6.27) |
| Age (Time Point 1) (mean (SD)) | 10.62 (0.24) | 10.64 (0.25) | 10.63 (0.24) |
| Age (Time Point 2) (mean (SD)) | 12.81 (0.22) | 12.80 (0.22) | 12.80 (0.21) |
| Age (Time Point 3) (mean (SD)) | 13.82 (0.19) | 13.83 (0.21) | 13.83 (0.21) |
| Age (Time Point 4) (mean (SD)) | 16.69 (0.24) | 16.68 (0.25) | 16.68 (0.23) |
| Age (Time Point 5) (mean (SD)) | 17.79 (0.38) | 17.83 (0.39) | 17.83 (0.39) |
| Age (Time Point 6) (mean (SD)) | 18.66 (0.48) | 18.67 (0.49) | 18.64 (0.49) |
| Age (Time Point 7) (mean (SD)) | 21.96 (0.50) | 21.95 (0.53) | 21.96 (0.51) |
| Age (Time Point 8) (mean (SD)) | 22.88 (0.49) | 22.90 (0.53) | 22.90 (0.52) |
| Age (Time Point 9) (mean (SD)) | 23.88 (0.48) | 23.88 (0.53) | 23.86 (0.51) |
| Age (Time Point 10) (mean (SD)) | 25.78 (0.48) | 25.77 (0.52) | 25.76 (0.51) |
| Age (Time Point 11) (mean (SD)) | 28.38 (0.52) | 28.38 (0.54) | 28.37 (0.53) |

#### 3.1.2 UK Biobank Cohort

UK Biobank is a large, population-based, prospective study, aiming to investigate contributing factors to a wide range of health-related outcomes ^33^. UK Biobank consists of over 500,000 participants, aged 40-69 years when recruited between 2006 and 2010 over 22 assessment centres throughout the UK (http://www.ukbiobank.ac.uk/).

Data collection occurred at both in-person assessment visits and remote online follow-up questionnaires. In-person assessment visits included an initial assessment visit (2006-2010), first repeat assessment visit (2012-2013), an imaging visit (2014+) and a repeat imaging visit (2019+) ^33, 34^. Online follow-up questionnaires included assessments such as mental health (2016-2017), experiences of pain (2019), health and well-being (2022+) and mental well-being (2022+). Demographics of UK Biobank participants used within the current study are shown in Table 2.

**Table 2.** Demographic table of UK Biobank participants.

|  | IL-6 Tertile Group |  |  |
| --- | --- | --- | --- |
|  | Bottom Third | Middle Third | Top Third |
| n | 13776 | 13640 | 12197 |
| Sex = Male (%) | 6162 ( 44.7) | 6291 ( 46.1) | 5737 ( 47.0) |
| BMI (log transformed) (mean (SD)) | 3.22 (0.13) | 3.30 (0.14) | 3.37 (0.17) |
| BMI (categorised) (%) |  |  |  |
| Underweight | 105 ( 0.8) | 50 ( 0.4) | 36 ( 0.3) |
| Healthy | 6894 ( 50.0) | 4033 ( 29.6) | 2291 ( 18.8) |
| Overweight | 5507 ( 40.0) | 6563 ( 48.1) | 4979 ( 40.8) |
| Obese | 1216 ( 8.8) | 2856 ( 20.9) | 4351 ( 35.7) |
| Morbidly Obese | 18 ( 0.1) | 98 ( 0.7) | 461 ( 3.8) |
| Missing | 36 ( 0.3) | 40 ( 0.3) | 79 ( 0.6) |
| Inflammatory condition = TRUE (%) | 1573 ( 11.4) | 2095 ( 15.4) | 2674 ( 21.9) |
| Taking inflammatory medication = TRUE (%) | 3138 ( 22.8) | 3692 ( 27.1) | 3822 ( 31.3) |
| Smoking status (%) |  |  |  |
| Previous | 4412 ( 32.0) | 4780 ( 35.0) | 4477 ( 36.7) |
| Current | 1196 ( 8.7) | 1352 ( 9.9) | 1612 ( 13.2) |
| Never | 8124 ( 59.0) | 7470 ( 54.8) | 6060 ( 49.7) |
| NA | 44 ( 0.3) | 38 ( 0.3) | 48 ( 0.4) |
| Townsend Deprivation Index (mean (SD)) | -1.56 (2.99) | -1.33 (3.07) | -0.90 (3.27) |
| IL-6 (Inverse Normal Transformed, Z-score) (mean (SD)) | -1.09 (0.53) | 0.00 (0.25) | 1.04 (0.50) |
| IL-6 (raw value, Olink Normalised) (mean (SD)) | -0.70 (0.30) | -0.01 (0.17) | 0.97 (0.77) |
| PHQ-2 (Initial time point) (mean (SD)) | 0.53 (1.03) | 0.56 (1.08) | 0.63 (1.16) |
| PHQ-2 (Repeat time point) (mean (SD)) | 0.46 (0.96) | 0.41 (0.86) | 0.45 (1.00) |
| PHQ-2 (Imaging time point) (mean (SD)) | 0.41 (0.96) | 0.43 (0.97) | 0.45 (0.95) |
| PHQ-2 (Repeat Imaging time point) (mean (SD)) | 0.32 (0.78) | 0.46 (0.96) | 0.39 (0.89) |
| PHQ-2 (Mental health time point) (mean (SD)) | 0.50 (1.03) | 0.52 (1.06) | 0.57 (1.14) |
| PHQ-2 (Pain time point) (mean (SD)) | 0.54 (1.05) | 0.59 (1.12) | 0.68 (1.21) |
| PHQ-2 (Health & Well-being time point) (mean (SD)) | 0.46 (1.01) | 0.48 (1.06) | 0.56 (1.19) |
| PHQ-2 (Mental well-being time point) (mean (SD)) | 0.52 (1.07) | 0.56 (1.12) | 0.64 (1.22) |
| Age (Initial time point) (mean (SD)) | 54.31 (8.13) | 57.30 (7.94) | 58.53 (7.60) |
| Age (Repeat time point) (mean (SD)) | 60.05 (7.38) | 61.87 (7.11) | 62.96 (6.61) |
| Age (Imaging time point) (mean (SD)) | 63.34 (7.82) | 65.78 (7.69) | 66.00 (7.61) |
| Age (Repeat Imaging time point) (mean (SD)) | 63.84 (7.40) | 66.71 (7.37) | 65.32 (6.76) |
| Age (Mental health time point) (mean (SD)) | 62.25 (7.84) | 64.77 (7.63) | 65.67 (7.33) |
| Age (Pain time point) (mean (SD)) | 64.96 (7.75) | 67.66 (7.57) | 68.35 (7.31) |
| Age (Health & Well-being time point)<br>(mean (SD)) | 67.78 (7.64) | 70.26 (7.47) | 70.93 (7.28) |
| Age (Mental well-being time point)<br>(mean (SD)) | 67.98 (7.54) | 70.40 (7.42) | 71.07 (7.22) |

### 3.2 Measures of depressive symptoms

#### 3.2.1 ALSPAC Cohort

The Short Mood and Feelings Questionnaire (SMFQ) was used to assess self-reported depressive symptoms at 11 time points between the ages of 10 to 28 years (Supplementary Figure 1, Supplementary Table 1). The SMFQ was administered via mail/email or in clinics. There were four clinic time points (ages 10, 12, 14 and 18 years) and seven remote self-reported (mail) time points (ages 17, 19, 22, 23, 24, 26 and 28 years). The SMFQ is a 13-item questionnaire that measures the presence of depressive symptoms within the last two weeks ^35^. The SMFQ has been used in clinical populations to assess depressive symptoms ^36^ and has been shown to predict clinical depression in ALSPAC ^35^. Supplementary Table 2 lists the SMFQ items. Each item response is scored from 0 to 2 (0 = “not true”, 1 = “sometimes”, 2 = “true”), where the total summed score ranges from 0 to 26 and where a higher score corresponds to worse depressive symptoms. The mean number of time points per participant was 6.12 (median = 6, mode =3).

#### 3.2.2 UK Biobank Cohort

Depressive symptoms were assessed at 8 time points (four in-person and four online follow-up questionnaire assessments) using questions from the Patient Health Questionnaire-2 (PHQ-2) which reflect depressed mood and anhedonia (Supplementary Table 4) ^37^. PHQ-2 has previously been shown to be a valid screening tool for detecting depression ^38–40^. The mean, standard deviation, min, max and interquartile range of ages at each time point is described in Supplementary Table 3 and Supplementary Figure 2. The mean number of time points per participant was 2.56 (median = 2, mode = 1).

### 3.3 Measures of blood serum IL-6

#### 3.3.1 ALSPAC Cohort

Blood samples were collected at age 9 years (mean age:9.86 years; SD:0.31) and high sensitivity serum CRP and IL-6 were measured in 5,059 participants. Details of laboratory methods are described in detail previously ^7^. Individuals with serum CRP ≥ 10mg/L (N = 60) were excluded from the main analysis to minimise confounding by chronic inflammatory condition or acute infection ^41^, consistent with previous studies ^7, 29^. The final sample used for analysis consisted of 4,999 participants.

#### 3.3.2 UK Biobank Cohort

Proteomic data was extracted by Olink by analysing blood samples collected at the initial assessment from a subset of UK Biobank participants (N = 54,219) (https://biobank.ctsu.ox.ac.uk/crystal/ukb/docs/Olink_proteomics_data.pdf), ^42^. This subset of participants consisted of 46,595 randomly selected participants from the initial assessment visit, 6,376 participants selected for the UKB-PPP study and 1,268 participants who participated in a COVID-19 repeat-imaging study at multiple visits^42^. 2,923 unique proteins were measured using Olink Explore 3072 Proximity Extension Assay. This including IL-6 protein which was measured in 44,076 participants. Further details on Olink proteomics data are described by UK Biobank here: https://biobank.ndph.ox.ac.uk/showcase/ukb/docs/Olink_proteomics_data.pdf, https://biobank.ndph.ox.ac.uk/showcase/ukb/docs/Olink_1536_B0_to_B7_Analysis_Report.pdf,

https://biobank.ndph.ox.ac.uk/showcase/ukb/docs/Olink_1536_B0_to_B7_Normaliza tion.pdf, https://biobank.ndph.ox.ac.uk/showcase/ukb/docs/Olink_1536_B0_to_B7_FAQ.pdf, https://biobank.ndph.ox.ac.uk/showcase/ukb/docs/PPP_Phase_1_QC_dataset_companion_doc.pdf. Individuals with CRP ≥ 10 mg/L (N = 1,758) were excluded from the main analysis, to minimize confounding by acute infection and keep analysis consistent to ALSPAC analysis. Details of blood sampling processing for CRP are described by UK Biobank here: https://biobank.ndph.ox.ac.uk/ukb/ukb/docs/haematology.pdf, https://biobank.ndph.ox.ac.uk/showcase/showcase/docs/serum_biochemistry.pdf, https://www.ukbiobank.ac.uk/media/oiudpjqa/bcm023_ukb_biomarker_panel_website_v1-0-aug-2015-edit-2018.pdf. The final sample used for analysis consisted of 40,069 participants (mean age for baseline IL-6 measurement: 56.6 years; SD:8.10).

### 3.4 Statistical analysis

#### 3.4.1 Deriving trajectories of depressive symptoms

Multi-level growth curve modelling was conducted in R, using the “lme4” package, to create population averaged trajectories of depression ^43^. Multi-level growth curve modelling was conducted in R, using the “lme4” package, to create population averaged trajectories of depression (Bates et al., 2015). Briefly, multi-level growth curve modelling clusters repeated measures within individuals. Unlike traditional linear regression, which treats each observation as independent, multi-level growth curve modelling recognises that repeated measures within the same individual are likely to be correlated, which reduces bias. Furthermore, multi-level growth curve modelling enables the exploration of individual trajectories of change over time. By allowing for random effects at both the individual level and the group level, this approach can capture not only mean population trends across the entire sample but also variations in trajectories among different individuals or groups.

Age was mean centred to 17.9 years in ALSPAC and 63 years in UK Biobank (the mean age of all assessments in each cohort) in order to improve model interpretation. Continuous covariate variables were Z-score scaled.

We assessed both linear and non-linear (quadratic, cubic and quartic) models. The fit of the model was assessed using Bayesian Information Criterion (BIC) and likelihood ratio test (LRT). A quartic model fitted the ALSPAC data best and a quadratic model fitted the UK Biobank data best (Supplementary Tables 5-6).

The models included repeated measures per participant of SMFQ scores for ALSPAC and PHQ-2 scores for UK Biobank and age at which the depression questionnaire was completed. In ALSPAC the intercept and four polynomial age terms were able to vary across individuals to capture each individual’s unique trajectory (ie. random intercept and random slopes model). In UK Biobank the intercept and only linear age terms were able to vary across individuals (ie. random intercept and random linear slope model). The model did not converge when also including a random quadratic slope term or when trying a cubic model.

To examine how IL-6 associated with changes in depressive symptoms, we split participants into IL-6 tertile groups ^7, 29^. The models included fixed effects of IL-6 tertile and an interaction of IL-6 tertiles with each of the fixed-effect age polynomial terms.

#### 3.4.2 Calculating mean depressive symptom scores

To assess the association between IL-6 tertile groups and development of symptoms over time, we created a population trajectory for each IL-6 tertile group. We then calculated the mean depressive symptoms scores at ages 10, 13, 16, 19, 22, 25 and 28 years in ALSAPC and ages 40, 50, 60, 70 and 80 years in UK Biobank for each of these trajectories, in the fully adjusted models. These ages were chosen to reduce the number of multiple tests performed whilst still capturing potentially important developmental changes over time. We then calculated the differences in mean depressive symptoms scores at each of these ages of the IL-6 tertile groups in a pair-wise manner. Further information on how these scores and their differences were calculated for the trajectories is presented elsewhere ^20^. Briefly, the depressive symptom scores were calculated for each IL-6 tertile group trajectory. The delta method (which incorporates the estimate, standard errors and confidence intervals) was then used to compare these two scores (ie. upper vs lower tertile, lower vs middle tertile, upper vs middle tertile in turn) revealing a mean difference in scores that are derived estimates from each trajectory. Differences in scores were transformed to Z-scores to compare results between ALSPAC and UK Biobank (detailed in Supplementary Methods). P-values were adjusted for multiple testing using the false discovery rate (FDR). The number of multiple tests were the number of different time points used to calculate scores for (ALSPAC: n tests = 7; UK Biobank: n tests = 5).

#### 3.4.3 Confounders

Confounders used in the ALSPAC models were the same as described in Edmondson-Stait et al. (2022). Three main models were used, the first was an unadjusted model with no covariates added, the second was adjusted for sex only and the third fully adjusted model further included covarying for log-transformed BMI (at age 9 years) and maternal education as a marker of socioeconomic status ^44, 45^. Maternal education was coded as a binary variable as either “CSE/O-level/Vocational education” or “A-level/degree level of education”. Sex was coded as a binary variable as either “Male” or “Female”. BMI (age 9 years) was calculated by dividing weight (kg) by squared height (meters). Distributions and participant counts of these variables are shown in Supplementary Figure 3.

Confounders used in the UK Biobank models were similar to those used in the ALSPAC cohort. Three main models were used, the first was an minimally adjusted model with covariates for protein batch and assessment centre at the initial assessment added, the second was additionally adjusted for sex, and the third fully adjusted model included further covarying for log-transformed BMI (at the time of blood sample collection), smoking status and Townsend deprivation index as a marker of socioeconomic status as these have been previously shown to associate with inflammation or psychiatric disorders ^46, 47^. Distributions and participant counts of these variables are shown in Supplementary Figure 4.

#### 3.4.4 Missing data

Missing outcome data in the trajectories analysis were addressed using full information maximum likelihood estimation (FIML), as part of the “lmer” function from the “lme4” package in R ^43, 48^. Briefly, this assumes that the probability of an individual missing a measure of depressive symptoms does not depend on their underlying depressive symptoms score at that occasion, given their observed depressive symptoms trajectory at other occasions. We included individuals into the analysis if they had at least one measurement of depression symptoms in order to maximise power ^49^.

#### 3.4.5 Sensitivity analyses

Sensitivity analyses involved investigating the impact of sex, tertile categorisation of IL-6, the impact of anti-inflammatory medication and impact of attrition. Previous studies have shown trajectories of depression are different for males and females ^20,21^. Therefore, we created a new variable that split the IL-6 tertiles by sex: female & bottom third IL-6 tertile, female & middle third IL-6 tertile, female & top third IL-6 tertile, male & bottom third IL-6 tertile, male & middle third IL-6 tertile and male & top third IL-6 tertile. The models were then run splitting the trajectories on this sex-split IL-6 tertile variable and analysed the same way as in the main analysis. To assess the effect of tertile categorisation of IL-6, we ran the analysis using a continuous measure of IL-6 (which was inverse normal transformed to achieve normal distribution and Z-score scaled) and compared the model estimates. The impact of inflammatory medication was assessed by removing individuals that might be taking medication that affects inflammation (ALSPAC N removed = 695; UK Biobank N removed = 10,652). In ALSPAC, the only measure available for medication at age 9 years (when IL-6 was measured) was a general variable of “Currently taking medication?”, therefore this may include medications that do not impact inflammation. In UK Biobank, anyone taking anti-inflammatory medications were removed (Supplementary Material; Supplementary Table 7). To assess attrition, we used linear regression to test for associations between IL-6 tertile on the number of questionnaires completed. In UK Biobank, the two imaging time points were excluded in this count as only a subset of individuals were invited to attend these appointments.

Additionally, we assessed the differences in markers of socioeconomic status used in ALSPAC and UK Biobank. To ensure consistency with our previous ALSPAC study we used maternal education as a marker of socioeconomic status ^8^. However, Townsend deprivation index was used as a marker of socioeconomic status in UK Biobank as no measure of maternal education was available. Therefore, we also conducted a sensitivity analysis in ALSPAC using Townsend deprivation index quintiles as a covariate in place of maternal education allowing a comparison of results between ALSPAC and UK Biobank (Supplementary Material, Supplementary Figure 5).

In UK Biobank various other sensitivity analyses were performed. Other factors that may affect inflammation included inflammatory conditions and high BMI. Individuals with an inflammatory condition were identified and removed (N removed = 6,342), using definitions of 49 conditions ^50^ (Supplementary Table 8). Individuals with BMI ≥ 40 were also removed (N removed = 577), as inflammation associates with high BMI (Supplementary Material) ^3^. Finally, to assess attrition due to death we removed people who had died after the initial baseline appointment. Further details are in the Supplementary Material. Similar analysis was not conducted in ALSPAC due to this cohort being a younger age.

## 4. Results

### 4.1 Sample characteristics of ALSPAC

A total of 4,999 individuals had serum IL-6 data and CRP < 10 mg/L, of these 4,835 had at least one measurement of depressive symptoms (measured by the SMFQ). Sample characteristics of this sample are shown in Table 1, split by IL-6 tertile.

### 4.2 Sample characteristics of UK Biobank

A total of 40,069 individuals had IL-6 data and CRP < 10 mg/L, of these 39,613 had at least one measurement of depressive symptoms (measured by PHQ-2; 18,958 had depressive symptoms measured only at the initial assessment). Sample characteristics of this sample are shown in Table 2, split by IL-6 tertile. IL-6 was measured in the initial assessment in which participant ages ranged from age 39-70 years (mean: 56.6 y; SD:8.10). The mean ages varied for participants in each IL-6 tertile (bottom third: mean: 54.31y, SD: 8.13; middle third: mean: 57.30y, SD: 7.94; top third: mean: 58.53y, SD: 7.60).

### 4.3 Associations of baseline IL-6 with subsequent depressive symptoms trajectories in ALSPAC

In the fully adjusted model (sex, BMI and maternal education), the overall pattern of depressive symptom trajectories increased from ages 10 to 20 years, followed by a plateau from 22 years onwards. The top third IL-6 tertile group had a higher trajectory compared to both the middle and bottom third IL-6 tertile groups, indicating increased depressive symptoms across this period (Figure 1A, Supplementary Table 9). However, confidence intervals overlapped across all IL-6 tertile group trajectories. Model estimates for all models (unadjusted, sex adjusted and fully adjusted are presented in Supplementary Table 10.

**Figure 1.**
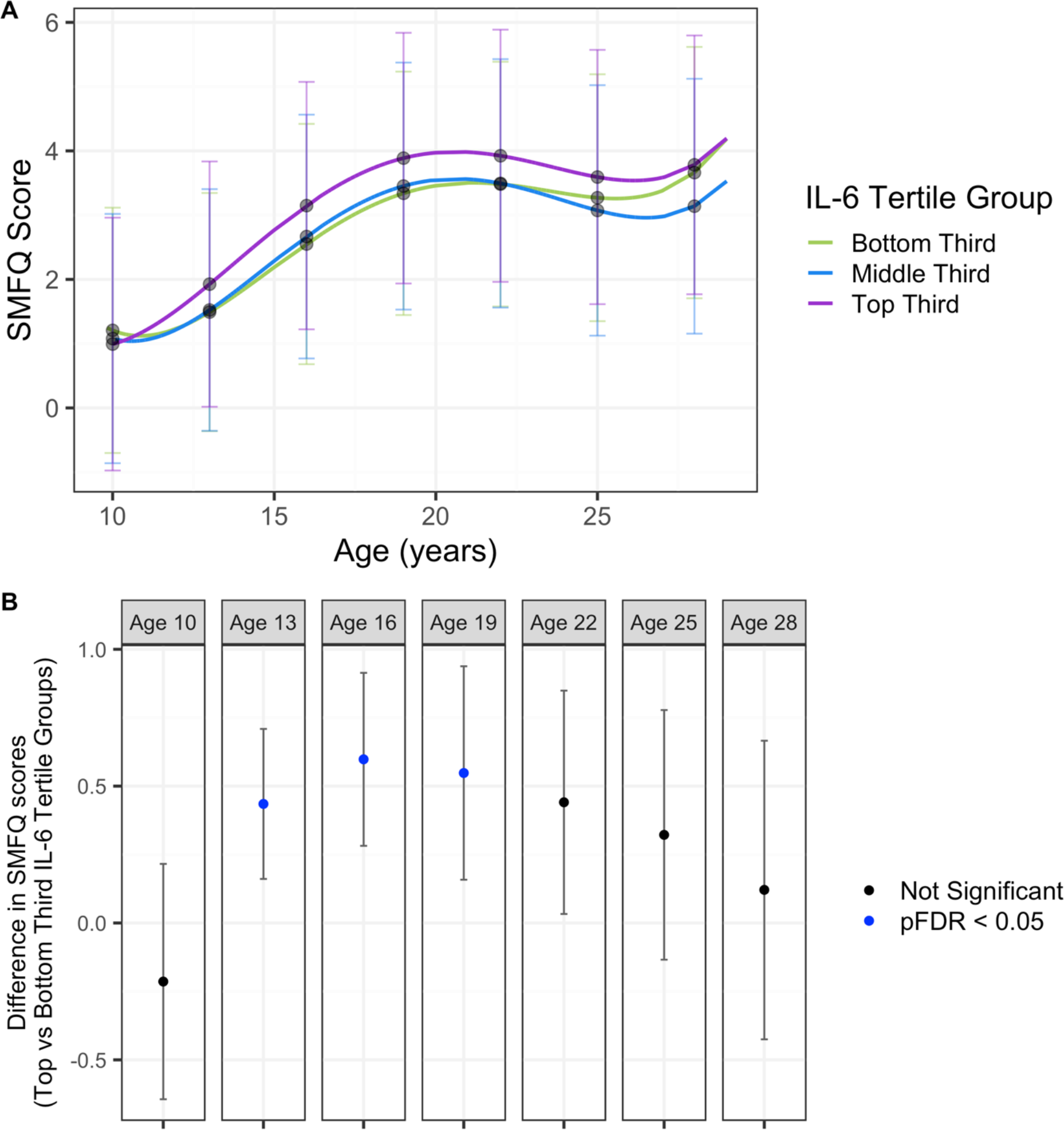
A. Depression trajectories in ALSPAC split by IL-6 tertile groups. B. Differences in depression scores in ALSPAC between top third and bottom third IL-6 tertiles. Results from the fully adjusted model. Mean depressive scores were calculated from the depression trajectories in each IL-6 tertile at ages 10, 13, 16, 19, 22, 25 and 28 years. Differences between the top third and bottom third IL-6 tertile trajectories was calculated using the delta method. P-values are corrected for multiple correction (FDR).

Given the difficulty in interpreting non-linear trajectories (i.e., positive linear polynomial terms, and negative quadratic polynomial terms), we report the mean differences at various ages across youth development between the bottom third and top third IL-6 tertile groups. There was evidence for a difference in depressive symptom scores between the top third and bottom third IL-6 tertiles across adolescence at ages 13 (SMFQ Score^diff^=0.435, 95%CI 0.161, 0.709, pFDR=0.0130), 16 years (SMFQ Score^diff^=0.598, 95%CI 0.282, 0.914, pFDR=0.0015) and 19 years (SMFQ Score^diff^=0.548, 95%CI 0.158, 0.938, pFDR=0.0414), but not the other ages tested (10, 22, 25 and 28 years) (Figure 1B,Table 3).

**Table 3.** Estimated differences in depression scores between IL-6 tertile top and bottom third trajectories at ages 10, 13, 16, 19, 22, 25 and 28 years, in ALSPAC. Results from the fully adjusted model.

| IL6 tertile: Top vs Bottom Third Age (years) | Difference (Z-score) | Difference (raw score) | 95% CI (raw score) | P (uncorrected) | P (FDR) |
| --- | --- | --- | --- | --- | --- |
| 10 | -0.012 | -0.214 | -0.644 - 0.216 | 0.3297 | 1 |
| 13 | 0.039 | 0.435 | 0.161 - 0.709 | 0.0019 | 0.0130 |
| 16 | 0.046 | 0.598 | 0.282 - 0.914 | 0.0002 | 0.0015 |
| 19 | 0.034 | 0.548 | 0.158 - 0.938 | 0.0059 | 0.0414 |
| 22 | 0.026 | 0.441 | 0.033 - 0.849 | 0.034 | 0.2383 |
| 25 | 0.017 | 0.322 | -0.134 - 0.778 | 0.1666 | 1 |
| 28 | 0.005 | 0.121 | -0.425 - 0.666 | 0.6649 | 1 |

There was evidence for differences in trajectories between females and males when splitting each IL-6 tertile group by sex. Depression trajectories and mean depressive scores across all IL-6 tertiles were worse in females compared to males (Figure 2A, Table 4, Supplementary Table 11). Females in the top third IL-6 tertile group generally had worse trajectories. There was evidence for a difference in depressive symptom scores between the top third and bottom third IL-6 tertiles in females, but not males, at ages 13 and 16 years (Figure 2B, Table 4, Supplementary Table 12).

**Figure 2.**
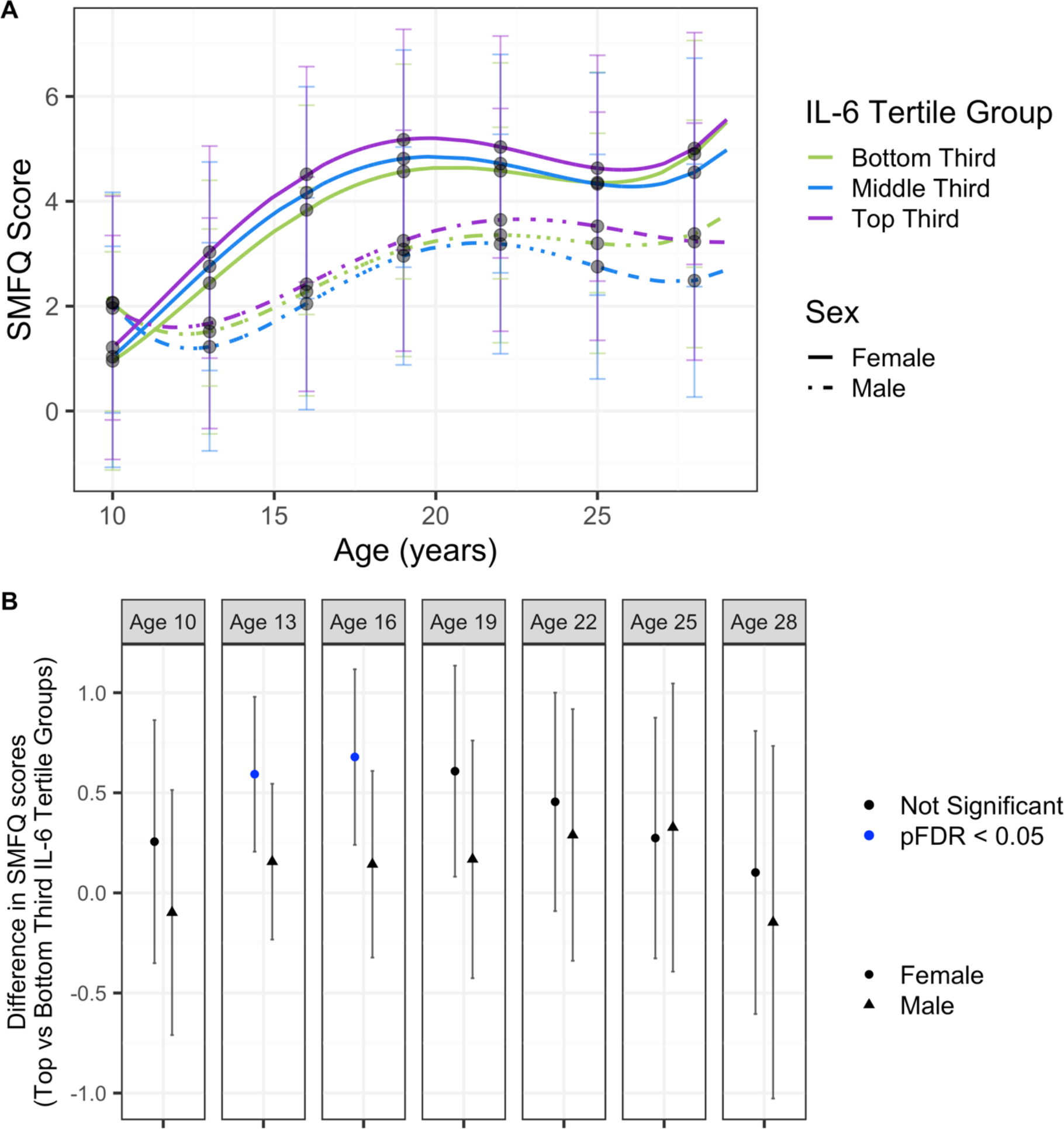
A. Depression trajectories in ALSPAC split by sex and IL-6 tertile groups. B. Differences in depression scores in ALSPAC between top third and bottom third IL-6 tertiles, in males and females separately. Results from the fully adjusted model. Mean depressive scores were calculated from the depression trajectories in each IL-6 tertile split by sex at ages 10, 13, 16, 19, 22, 25 and 28 years. Differences between the top third and bottom third IL-6 tertile trajectories was calculated using the delta method. P-values are corrected for multiple correction (FDR).

**Table 4.** Estimated differences in depression scores between IL-6 tertile top and bottom third trajectories at ages 10, 13, 16, 19, 22, 25 and 28 years, in ALSPAC, split by sex. Results from the fully adjusted model.

|  | IL6 tertile: Top vs Bottom Third Age (years) | Difference (Z-score) | Difference (raw score) | 95% CI (raw score) | P (uncorrected) | P (FDR) |
| --- | --- | --- | --- | --- | --- | --- |
| Females | 10 | 0.01 | 0.256 | -0.351 - 0.863 | 0.4086 | 1 |
|  | 13 | 0.038 | 0.593 | 0.206 - 0.979 | 0.0027 | 0.0187 |
|  | 16 | 0.038 | 0.679 | 0.24 - 1.117 | 0.0024 | 0.0168 |
|  | 19 | 0.028 | 0.608 | 0.081 - 1.135 | 0.0237 | 0.1662 |
|  | 22 | 0.02 | 0.455 | -0.091 - 1 | 0.1024 | 0.7169 |
|  | 25 | 0.011 | 0.274 | -0.327 - 0.875 | 0.3717 | 1 |
|  | 28 | 0.004 | 0.102 | -0.605 - 0.809 | 0.7778 | 1 |
| Males | 10 | -0.004 | -0.098 | -0.71 - 0.514 | 0.7538 | 1 |
|  | 13 | 0.01 | 0.156 | -0.233 - 0.545 | 0.4318 | 1 |
|  | 16 | 0.008 | 0.143 | -0.323 - 0.609 | 0.5466 | 1 |
|  | 19 | 0.007 | 0.168 | -0.426 - 0.761 | 0.5803 | 1 |
|  | 22 | 0.011 | 0.289 | -0.339 - 0.918 | 0.3668 | 1 |
|  | 25 | 0.011 | 0.327 | -0.393 - 1.046 | 0.3736 | 1 |
|  | 28 | -0.004 | -0.147 | -1.027 - 0.734 | 0.7437 | 1 |

Similar results were also seen when using a continuous measure of IL-6, when individuals taking any medication were removed and when using Townsend deprivation index quintiles in place of maternal education (Supplementary Tables 13- 15). These results were unlikely to be bias by attrition as IL-6 tertiles did not associate with the number of completed questionnaires (Supplementary Table 16, Supplementary Figure 6).

### 4.4 Associations of baseline IL-6 with subsequent depressive symptoms trajectories in UK Biobank

In the fully adjusted model (sex, BMI, batch, assessment centre, Townsend deprivation index and smoking status), the overall pattern of depressive symptom trajectories decreased from age 40 years until mid 60 years where they begin to increase. The top third IL-6 tertile group had a higher trajectory compared to both the middle and bottom third IL-6 tertile groups, indicating increased depressive symptoms across this period (Figure 3A, Supplementary Table 17). However, confidence intervals overlapped across all IL-6 tertile group trajectories. Model estimates for all models (unadjusted, sex adjusted and fully adjusted are presented in Supplementary Table 18.

**Figure 3.**
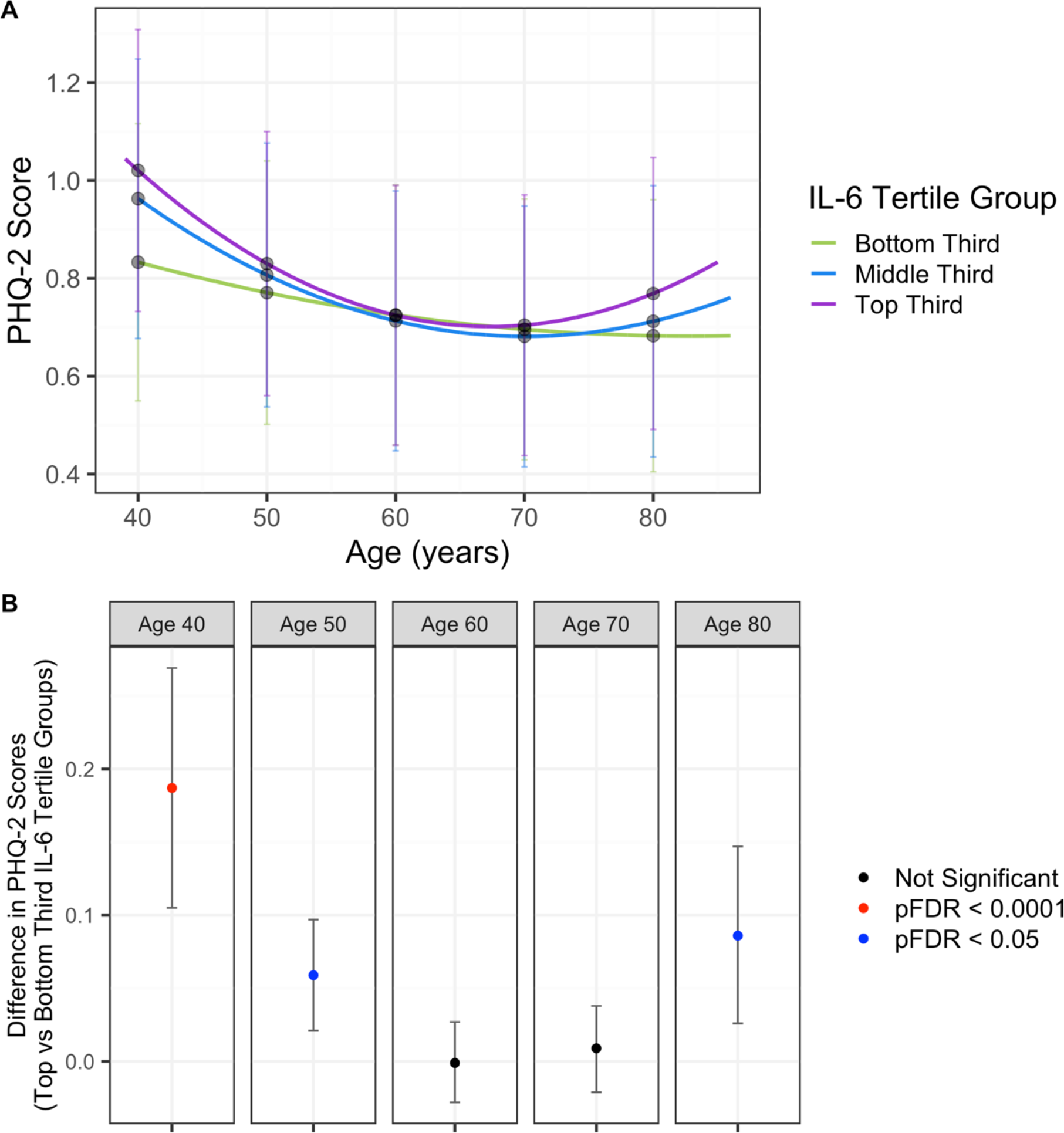
A. Depression trajectories in UK Biobank split by IL-6 tertile groups. **B. Differences in depression scores in UK Biobank between top third and bottom third IL-6 tertiles.** Results from the fully adjusted model. Mean depressive scores were calculated from the depression trajectories in each IL-6 tertile at ages 40, 50, 60, 70 and 80 years. Differences between the top third and bottom third IL-6 tertile trajectories was calculated using the delta method. P-values are corrected for multiple correction (FDR).

There was evidence for a difference in depressive symptom scores between the top third and bottom third IL-6 tertile groups at ages 40 (PHQ-2 Score^diff^=0.187, 95%CI 0.105, 0.269, pFDR<0.0001), 50 (PHQ-2 Score^diff^=0.059, 95%CI 0.021, 0.097, pFDR=0.0022) and 80 years (PHQ-2 Score^diff^=0.086, 95%CI 0.026, 0.147, pFDR=0.0257), but not the other ages tested (Figure 3B, Table 5).

**Table 5.**
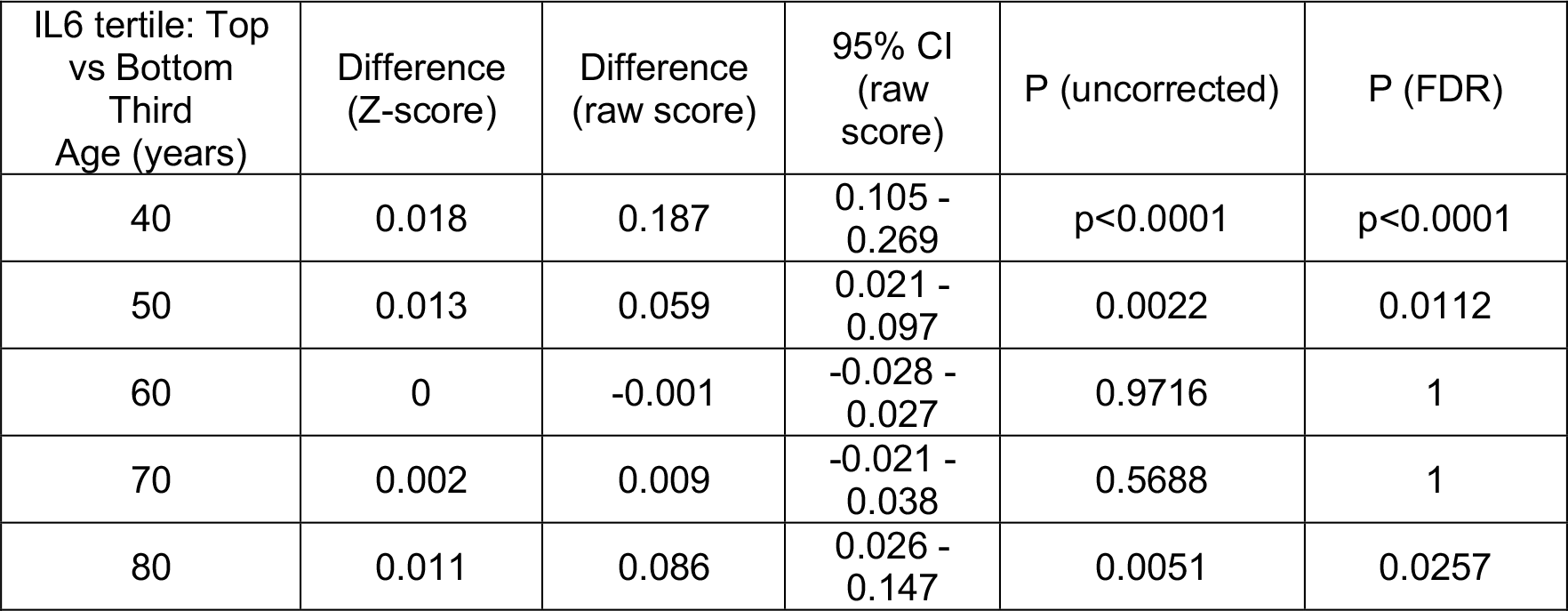
Estimated differences in depression scores between different IL-6 tertile trajectories at ages 40, 50, 60, 70 and 80 years, in UK Biobank. Results from the fully adjusted model.

| IL6 tertile: Top vs Bottom Third Age (years) | Difference (Z-score) | Difference (raw score) | 95% CI (raw score) | P (uncorrected) | P (FDR) |
| --- | --- | --- | --- | --- | --- |
| 40 | 0.018 | 0.187 | 0.105 - 0.269 | p<0.0001 | p<0.0001 |
| 50 | 0.013 | 0.059 | 0.021 - 0.097 | 0.0022 | 0.0112 |
| 60 | 0 | -0.001 | -0.028 - 0.027 | 0.9716 | 1 |
| 70 | 0.002 | 0.009 | -0.021 - 0.038 | 0.5688 | 1 |
| 80 | 0.011 | 0.086 | 0.026 - 0.147 | 0.0051 | 0.0257 |

There was evidence for differences between females and males when splitting each IL-6 tertile group by sex. Depression trajectories and mean depressive scores across all IL-6 tertiles were generally worse in females compared to males (Figure 4A, Supplementary Table 19). There was evidence for a difference in depressive symptom scores between the top third and bottom third IL-6 tertiles in both males and females at age 40 years, in males only at age 50 years, but not for ages 60, 70 or 80 years (Figure 4B, Table 6, Supplementary Table 20).

**Figure 4.**
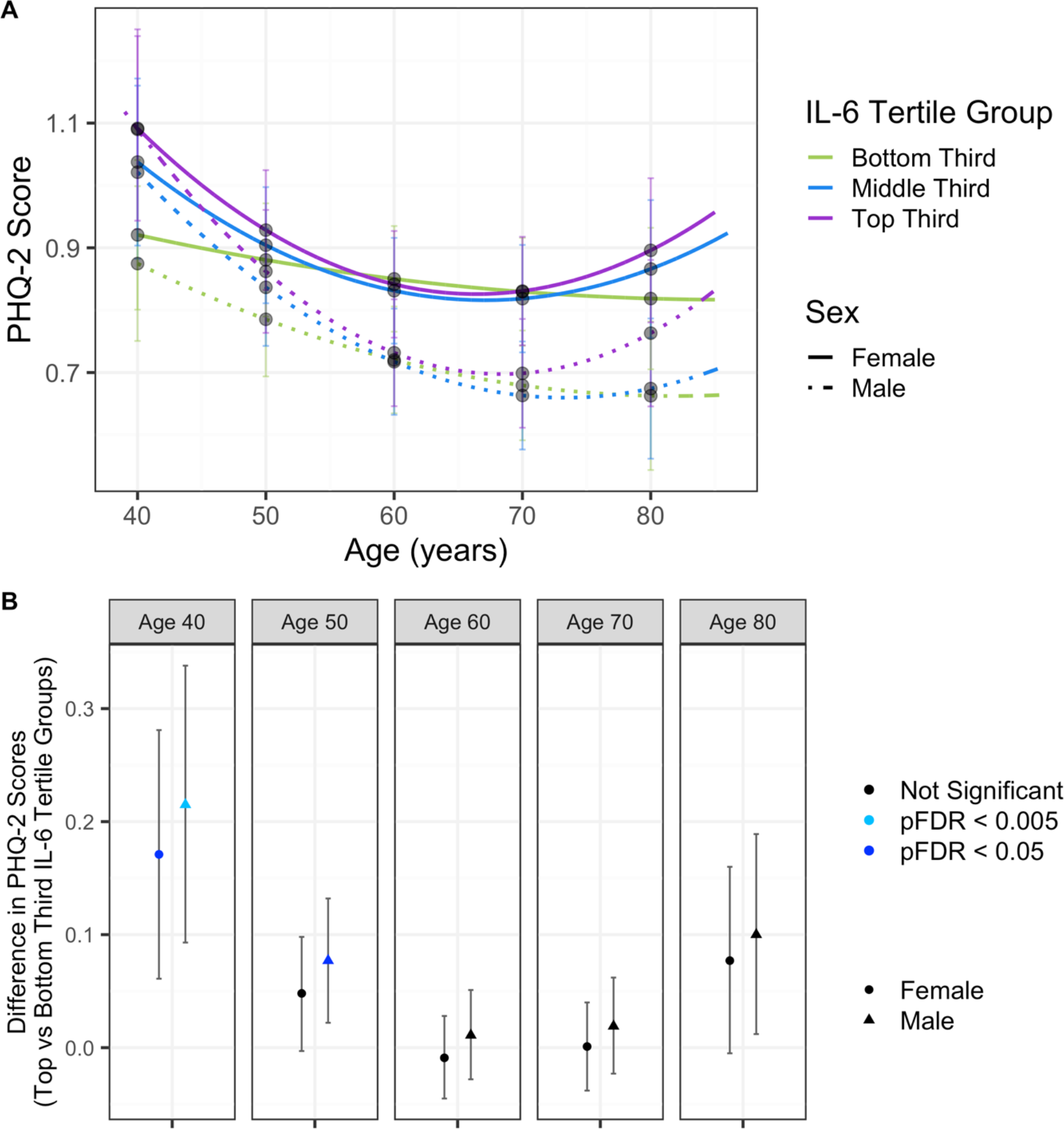
A. Depression trajectories in UK Biobank split by sex and IL-6 tertile groups. B. Differences in depression scores in UK Biobank between top third and bottom third IL-6 tertiles, in males and females separately. Results from the fully adjusted model. Mean depressive scores were calculated from the depression trajectories in each IL-6 tertile split by sex at ages 40, 50, 60, 70 and 80 years. Differences between the top third and bottom third IL-6 tertile trajectories was calculated using the delta method. P-values are corrected for multiple correction (FDR).

**Table 6.** Estimated differences in depression scores between different IL-6 tertile trajectories at ages 40, 50, 60, 70 and 80 years, in UK Biobank, split by sex. Results from the fully adjusted model.

|  | IL6 tertile:<br>Top vs<br>Bottom<br>Third<br>Age<br>(years) | Difference<br>(Z-score) | Difference<br>(raw score) | 95% CI<br>(raw<br>score) | P (uncorrected) | P (FDR) |
| --- | --- | --- | --- | --- | --- | --- |
| Females | 40 | 0.013 | 0.171 | 0.061 -<br>0.281 | 0.0024 | 0.012 |
|  | 50 | 0.008 | 0.048 | -0.003 -<br>0.098 | 0.0634 | 0.3168 |
|  | 60 | -0.002 | -0.009 | -0.045 -<br>0.028 | 0.6386 | 1 |
|  | 70 | 0 | 0.001 | -0.038 -<br>0.04 | 0.9563 | 1 |
|  | 80 | 0.008 | 0.077 | -0.005 -<br>0.16 | 0.0655 | 0.3276 |
| Males | 40 | 0.014 | 0.215 | 0.093 -<br>0.338 | 0.0006 | 0.0028 |
|  | 50 | 0.011 | 0.077 | 0.022 -<br>0.132 | 0.0065 | 0.0324 |
|  | 60 | 0.002 | 0.011 | -0.028 -<br>0.051 | 0.565 | 1 |
|  | 70 | 0.004 | 0.019 | -0.023 -<br>0.062 | 0.3693 | 1 |
|  | 80 | 0.009 | 0.1 | 0.012 -<br>0.189 | 0.0257 | 0.1283 |

Similar results were also seen when using a continuous measure of IL-6, when individuals taking anti-inflammatory medication were removed, when individuals with inflammatory conditions were removed, when individuals with BMI ≤ 40 were removed and when subsetting to only participants that were alive after the initial appointment (Supplementary Tables 21-25). However, these results may have been biased by attrition as IL-6 tertiles were associated with the number of completed questionnaires in the sample of participants that remained alive after the initial assessment (Supplementary Table 26, Supplementary Figures 7-8).

## 5. Discussion

Longitudinal trajectories of depressive symptoms were modelled to investigate the effects of baseline IL-6 on depressive symptoms in two cohorts spanning different stages of the life course (ALSPAC and UK Biobank). Higher IL-6 was associated with worse trajectories of depression symptoms across the life course. This relationship was stronger the younger cohort (ALSPAC), compared to the older cohort (UK Biobank). Sex differences were also consistent in both cohorts but stronger in the younger cohort (ALSPAC), where the association between higher IL-6 and worse depression trajectories were stronger in females compared to males.

The main strengths of this study are the use of two large-scale population cohorts with prospectively collected data and repeated measures of depression symptoms at 11 assessments across ages 9 to 28 years in ALSPAC and 8 assessments across ages 40 to 80 years in UK Biobank. This permitted the investigation of identifying the ages where increased IL-6 associated with worse depression trajectories and whether these effects were persistent across different stages of the life course.

Similar relationships were observed between IL-6 and depression trajectories in two different cohorts despite their heterogeneity and no overlap in ages. Also presented is an alternative way of interpreting trajectory results by looking at mean differences in scores at ages, which has only recently been developed in the field of longitudinal epidemiology ^20, 51^.

The overall pattern of depression trajectories in ALSPAC was consistent with previous studies of the same cohort ^20, 52^. Depressive symptoms increased from ages 10 to 20 years, followed by a plateau from 22 years onwards. Depression trajectories have been modelled in other cohorts and show a similar pattern to the UK Biobank results in this current study ^22^, whereby symptoms decrease from age 40 years until mid 60 years where they begin to increase again. There was evidence that people with higher IL-6 (ie. in the top third IL-6 tertile group) had worse trajectories than those with lower IL-6 (ie. in bottom third and middle third IL-6 tertile groups), with the greatest difference in mean depressive symptom scores between the top third and bottom third IL-6 tertile groups observed at ages 13, 16 and 19 years in ALSPAC and 40, 50 and 80 years in UK Biobank. The Z-scores of the mean differences for these ages were also comparable between ALSPAC and UK Biobank, with slightly larger mean differences in ALSPAC (0.039-0.046) compared to UK Biobank (0.011-0.018). This suggests that at various points across the life course, higher IL-6 associates with worse depression symptom trajectories, with a relatively greater impact of IL-6 on depressive symptoms in younger compared to older people. This could be due to inflammation coinciding with a vulnerable period of neurodevelopment in younger individuals ^53, 54^. Additionally, it may be that other environmental and social factors having more prominent effects on depression at later stages of the life course ^55^. It should also be noted that there is a difference in IL-6 assay method used in ALSPAC and UK Biobank. ALSPAC used ELISA which is commonly used for assessing IL-6 measures. Whereas UK Biobank used Olink which is a high-throughput method and may not be as accurate as ELISA. Additionally, these measures are on different scales. ALSPAC IL-6 data is provided as raw pg/ml measurements whereas UK Biobank is provided after they apply an in-house normalisation method which involves a log2 transformation.

The findings in this study are in line with previous studies. Previous studies have shown higher IL-6 is associated with depressive symptoms in both a cross-sectional and cross-lag relationship ^3, 7, 13, 16, 29, 56^. The study presented in chapter 4 showed IL- 6 was associated with the total number of depressive episodes, representing increased burden of depression in ALSPAC ^8^. Cross-sectionally, an inflammatory subgroup of depression has been shown to associate with depression severity ^10^.

Another study using latent class analysis in ALSPAC showed that baseline serum IL- 6 levels was associated with a trajectory group of persistently worse depressive symptoms from ages 10 to 19 years ^16^. This current study complemented and extended these studies in numerous ways. Firstly, by extending the age range investigated in ALSPAC to also include a period of early adulthood (up to 28 years) in which development of psychiatric disorders can occur. Secondly, analysis was conducted in cohorts of both younger (ALSPAC) and older (UK Biobank) age with prospectively collected data. Thirdly, population level trajectories of depressive symptoms were assessed, rather than more probabilistic groups of individuals from latent class analysis.

There was also strong evidence of sex specific effects of IL-6 on depression trajectories in ALSPAC and weaker evidence in UK Biobank. Previous studies have shown that females have worse depression trajectories than males in ALSPAC ^20–22^. Here, in addition to showing this, sex-differences in depression trajectories were also shown to persist into older adulthood (39-86 years). This is consistent with findings in other cohorts ^22^. In ALSPAC, there was evidence that the difference in depressive scores between the top third and bottom third IL-6 tertile at ages 13 and 16 years was greater in females than in males. Similar findings have been reported elsewhere, showing that IL-6 associates with more severe depression in female but not male adolescents ^24^. This could be due to hormonal changes that occur during pubertal development ^57^. Female sex hormones, such as oestrogen have also been shown to have effects on the immune system, though depending on context can have both anti or pro-inflammatory effects ^58^. However, there may also be methodological explanations for this sex difference, such as there are a greater number of females than males in ALSPAC. Whereas in UK Biobank the differences in scores between the top third and bottom third IL-6 tertile remained in both males and females for ages 40 years but occurred only in males at age 50 years and diminished at age 80 years. This could be attributed to a variety of explanations including that in general inflammation increases with age ^59^. In UK Biobank the ages of participants at the initial assessment when IL-6 was measured varied from age 39 to 70 years. Whereas in ALSPAC IL-6 was measured at age 9 years. This led to differences in the mean ages for UK Biobank participants for each IL-6 tertile group, with an older mean age for each tertile as IL-6 increases. However, age was included in the models. Future studies should assess the relationship of inflammatory markers measured at the same age on depression trajectories in older individuals to strengthen the findings in this current study.

However, extensive sensitivity analyses from both cohorts showed these findings persisted when controlling for factors that typically affect depression and inflammation. In ALSPAC, these findings were robust against adjusting for covariates sex, BMI and socioeconomic markers (maternal education or Townsend deprivation index quintiles). Sensitivity analyses removing individuals taking any medication also resulted in similar model coefficients. In UK Biobank, these findings were robust against adjusting for covariates sex, BMI, Townsend deprivation index and smoking status.

The UKB-PPP study was enriched for people with ill health and therefore the sample used for the UK Biobank analysis may be bias to this ^42^. This was accounted for in the sensitivity analyses removing individuals with an inflammatory condition, with BMI ≥ 40 or who were taking anti-inflammatory medication which showed similar effects to the main analysis. In both ALSPAC and UK Biobank, using a continuous measure of IL-6 showed similar results to using IL-6 tertile groups.

However, it should be noted that these effect sizes are small, and the confidence intervals are wide (despite the large sample sizes). This could be due to some limitations of the study. Both ALSPAC and UK Biobank are population-based cohorts rather than clinical cohorts. Further, UK Biobank participants are more likely to be female and living in less socioeconomically deprived areas than the general population ^60^. Additionally, although the age range of participants in UK Biobank is 39 to 86 years, the mean participants age is between 57 and 70 years for each time point (Supplementary Table 3). This contributes to higher confidence intervals in the distal ages, and therefore results should be interpreted with this in consideration.

There are also other limitations to consider in this study. Both ALSPAC and UK Biobank suffer from attrition and in UK Biobank 48% only had one measurement of depressive symptoms. These individuals were retained in the analysis as they contribute to the relationship between IL-6 and depression, and a key advantage of multi-level models is that it uses FIML to account for missing outcome data.

However, if the data is not missing at random then this method would be biased. We conducted sensitivity tests and found that IL-6 tertile group associated with the number of times a participant completed a questionnaire associated in UK Biobank but not in ALSPAC. In UK Biobank this sensitivity analysis was done in participants that remained alive after the initial assessment, due to this cohort being an older sample, and excluded the two imaging appointments. This suggests that there is likely some bias between IL-6 tertile group and subsequent attrition in UK Biobank, but not in ALSPAC.

In conclusion, the findings in this study suggest that high IL-6 associates with worse depression symptom trajectories observed at different stages of the life course, with stronger associations in younger individuals. On further analysis of sex differences, this association was stronger in females, compared to males in early adolescence. Whereas weaker sex differences were observed in later life. Future studies could also investigate the trajectories of different depression subtypes, such as atypical depression, and whether inflammatory proteins influence their trajectories across the life course.

## Acknowledgements

This research was funded by the Wellcome Trust (Grant No. 108890/Z/15/Z). For the purpose of open access, the authors have applied a CC BY public copyright licence to any Author Accepted Manuscript version arising from this submission.

The UK Medical Research Council and Wellcome (Grant No.: 217065/Z/19/Z) and the University of Bristol provide core support for ALSPAC. This publication is the work of the authors and Amelia Edmondson-Stait and Alex Kwong will serve as guarantors for the contents of this paper. A comprehensive list of grants funding is available on the ALSPAC website: (http://www.bristol.ac.uk/alspac/external/documents/grant-acknowledgements.pdf); This research was specifically funded by the MRC (MR/M006727/1 and GO701503/85179), Wellcome Trust (08426812/Z/07/Z)], Wellcome Trust and MRC (092731), NIH (PD301198-SC101645).

UK Biobank Data acquisition and analyses were conducted using the UK Biobank Resource under approved project #4844.

ED was supported by the UK Research and Innovation (Grant No. EP/S02431X/1), UK Research and Innovation Centre for Doctoral Training in Biomedical AI at the University of Edinburgh, School of Informatics. ASFK is funded by a Wellcome Early Career Award (Grant ref: 227063/Z/23/Z). AMM is supported by Wellcome Trust Investigator Award (Grant ref: 220857/Z/20/Z).

GMK acknowledges funding support from the UK Medical Research Council (MRC) via the Integrative Epidemiology Unit (IEU) at the University of Bristol (MC_UU_00032/06), and additional funding from the Wellcome Trust (201486/Z/16/Z and 201486/B/16/Z), the MRC (MR/W014416/1; MR/S037675/1; and The CHECKPOINT Hub, APP4735-GTEE-2024), and the UK National Institute of Health and Care Research (NIHR) Bristol Biomedical Research Centre (NIHR 203315). The views expressed are those of the authors and not necessarily those of the UK NIHR or the Department of Health and Social Care.

We are extremely grateful to all the families who took part in both the ALSPAC and UK Biobank studies, the whole ALSPAC and UK Biobank teams, which includes interviewers, computer and laboratory technicians, clerical workers, research scientists, volunteers, managers, receptionists, midwifes and nurses.

## Conflict of Interest

None to declare.

## Data availability

The data used in the present study is available from UK Biobank and ALSPAC with restrictions applied. Data were used under license and thus not publicly available. Access to the UK Biobank data can be requested through a standard protocol (https://www.ukbiobank.ac.uk/register-apply/). The ALSPAC study website contains details of all data available: http://www.bristol.ac.uk/alspac/researchers/our-data. Code used for analysis will be made publicly available on GitHub (www.github.com/ameliaES/) upon publication of this article.

## Notes

### Competing Interest Statement

The authors have declared no competing interest.

### Author Declarations

Ethical approval for UK Biobank analyses was granted by the North West Multi-centre Research Ethics Committee as a Research Tissue Bank approval (reference: 11/NW/0382). Ethical approval for Avon Longitudinal Study of Parents And Children was obtained from the Avon Longitudinal Study of Parents And Children Ethics and Law Committee and the Local Research Ethics Committees.

