## Supplementary Material for "Associations between IL-6 and trajectories of depressive symptoms across the life course: Evidence from ALSPAC and UK Biobank cohorts"

### Supplementary Methods

### ALSPAC Cohort

Study data [1-3] were collected and managed using REDCap electronic data capture tools hosted at the University of Bristol [4]. REDCap (Research Electronic Data Capture) is a secure, web-based software platform designed to support data capture for research studies. Please note that the study website contains details of all the data that is available through a fully searchable data dictionary and variable search tool: http://www.bristol.ac.uk/alspac/researchers/our-data/. Ethical approval for the study was obtained from the ALSPAC Ethics and Law Committee and the Local Research Ethics Committees. Consent for biological samples has been collected in accordance with the Human Tissue Act (2004). Informed consent for the use of data collected via questionnaires and clinics was obtained from participants following the recommendations of the ALSPAC Ethics and Law Committee at the time.

### Z-score calculation

Z-scores of the differences between depression questionnaire scores were calculated to compare results between ALSPAC and UK Biobank. This was done in R by calculating the difference in scores / ((upper 95% CI – lower 95% CI) / 3.1999 ) * sqrt(sample size).

### Sensitivity analysis using Townsend deprivation index quintiles in ALSPAC

The socioeconomic variables used for ALSPAC and UK Biobank were different in the main analysis. In UK Biobank we used Townsend deprivation index as a continuous score of material deprivation calculated from using data on non-car ownership, non-home ownership, unemployment and overcrowding within geographic regions [5]. A sensitivity analysis was performed in ALSPAC where Townsend deprivation index quintiles were included as a covariate in place of maternal education so results across ALSPAC and UK Biobank could be compared.

Townsend deprivation index quintiles were measured at three timepoints in ALSPAC (age 4, 6 and 16 years). The majority of individuals did not change in the quintile they belonged to across these three time points (Supplementary Figure 5). The variable measured at the 6-year time point was used as the covariate in the sensitivity analysis as this was closest to the time point at which blood samples were taken for IL-6 measurements.

### Categorised definitions for BMI and BMI ≥ 40 BMI sensitivity analysis

BMI was categorised into the following groups in UK Biobank: BMI < 18.5 = underweight, BMI 18.5 – 24.9 = healthy weight, BMI 25 – 29.9 = overweight, BMI 30-39.9 = Obese and BMI ≥ 40 = morbidly obese [6]. Numbers of individuals in each group are shown in the demographic table (Table 2). Individuals with BMI ≥ 40 were removed in a sensitivity analysis to ensure any inflammation due to high BMI was not affecting the results. Similar analysis was not conducted in ALSPAC as these BMI category definitions are for adults.

### Sensitivity analysis of alive UK Biobank participants and number of completed questionnaires covariate

To assess whether the number of questionnaires someone had completed affected the results we first removed people who had died (N removed = 2,815) after the initial baseline appointment and secondly included a covariate for the number of questionnaires someone had completed (excluding the two imaging time points as only a subset of individuals were invited to attend these appointments).

### Medication Sensitivity Analysis

In both cohorts we removed individuals that might be taking medication that affects inflammation (ALSPAC N removed = 695; UK Biobank N removed = 10,652). In ALSPAC, the only measure available for medication at age 9 years (when IL-6 was measured) was a general variable of “Currently taking medication?”, therefore this may include medications that do not impact inflammation. In UK Biobank, anyone taking anti-inflammatory medications were removed (Supplementary Table 7).

### Supplementary Figures

Supplementary Figure 1. Box plot of ages of individuals at each appointment time point in ALSPAC.

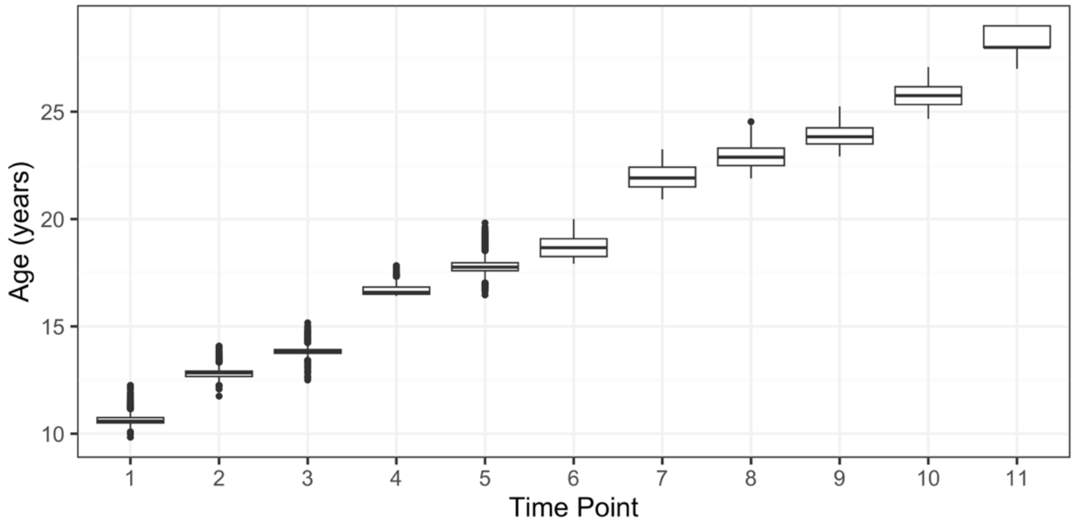

Supplementary Figure 2. Box plot of ages of individuals at each appointment time point in UK Biobank.

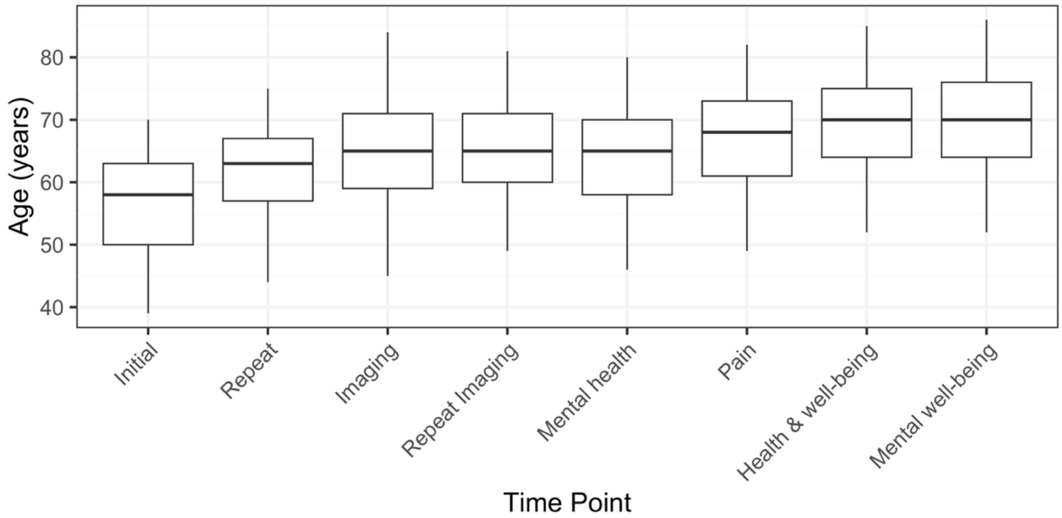


Supplementary Figure 3. Plots of covariates per IL-6 tertile group in ALSPAC. A. Number of females and males, B. Maternal education, C. BMI, D. log transformed BMI. Abbreviations: A-level/Degree, A/D; CSE/O-level/Vocational Qualification, C/O/N.

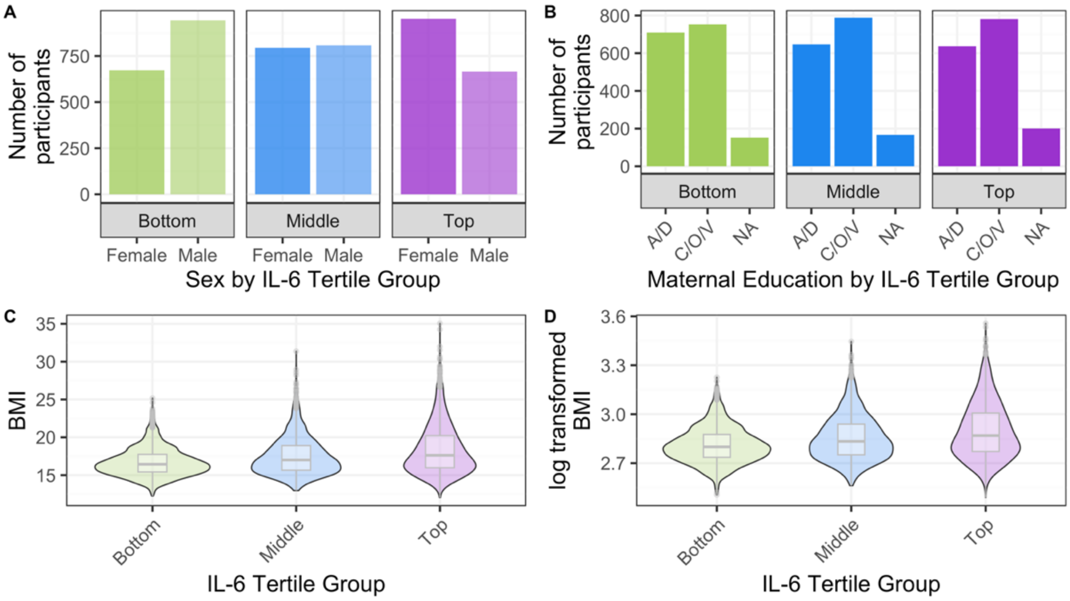


Supplementary Figure 4. Plots of covariates per IL-6 tertile group in UK Biobank. A. Number of females and males, B. Townsend deprivation index, C. Smoking status, D. BMI, E. log transformed BMI.

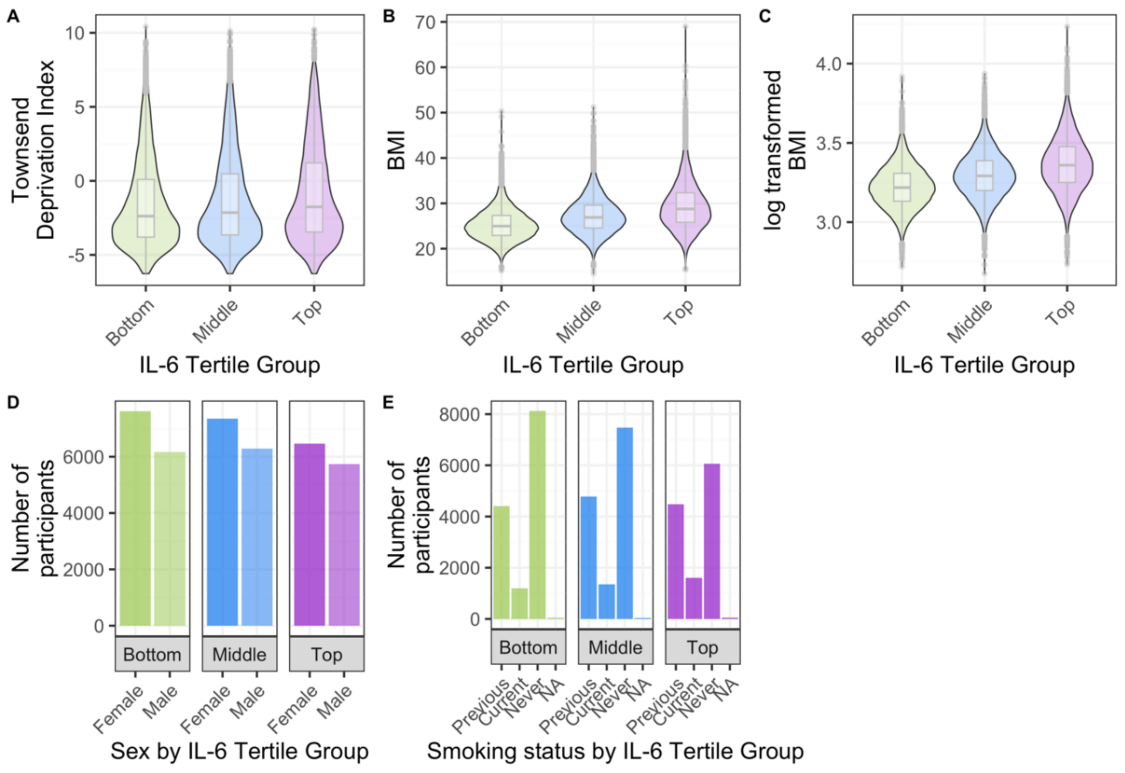
A
Townsend Deprivation Index
IL-6 Tertile Group
Bottom
Middle
Top
B
BMI
C
log transformed BMI
D
Number of participants
Female
Male
Sex by IL-6 Tertile Group
E
Previous
Current
Never
NA
Smoking status by IL-6 Tertile Group

Supplementary Figure 5. Alluvial plot of Townsend deprivation index quintile in ALSPAC for three time points.

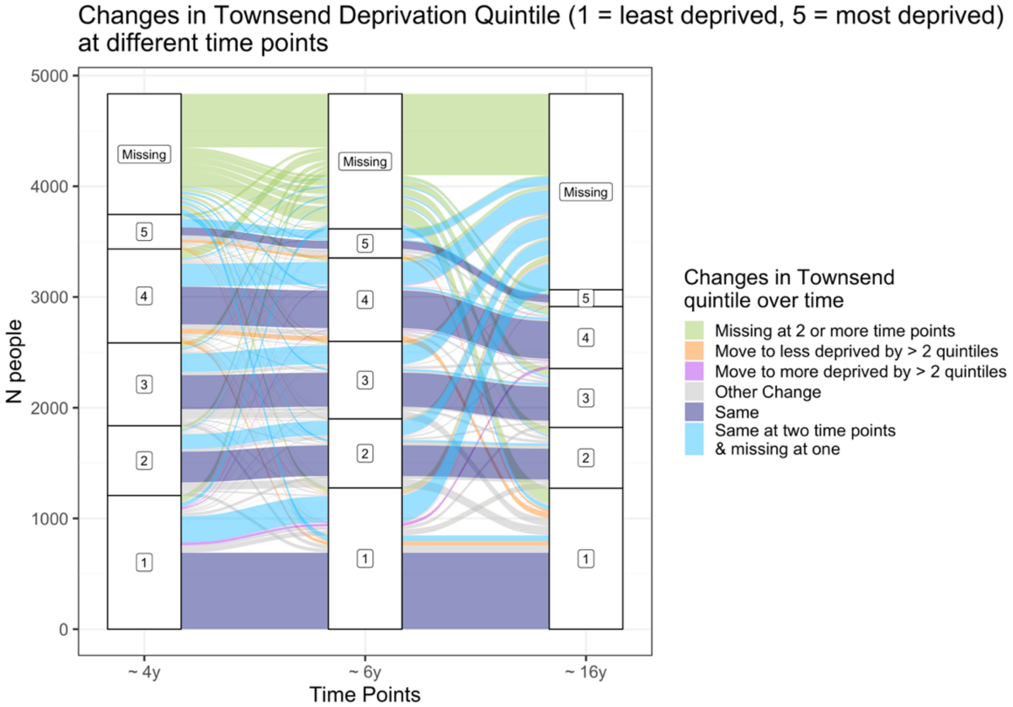


Supplementary Figure 6. Number of completed SMFQ questionnaires in ALSPAC. Coloured by IL-6 tertile.

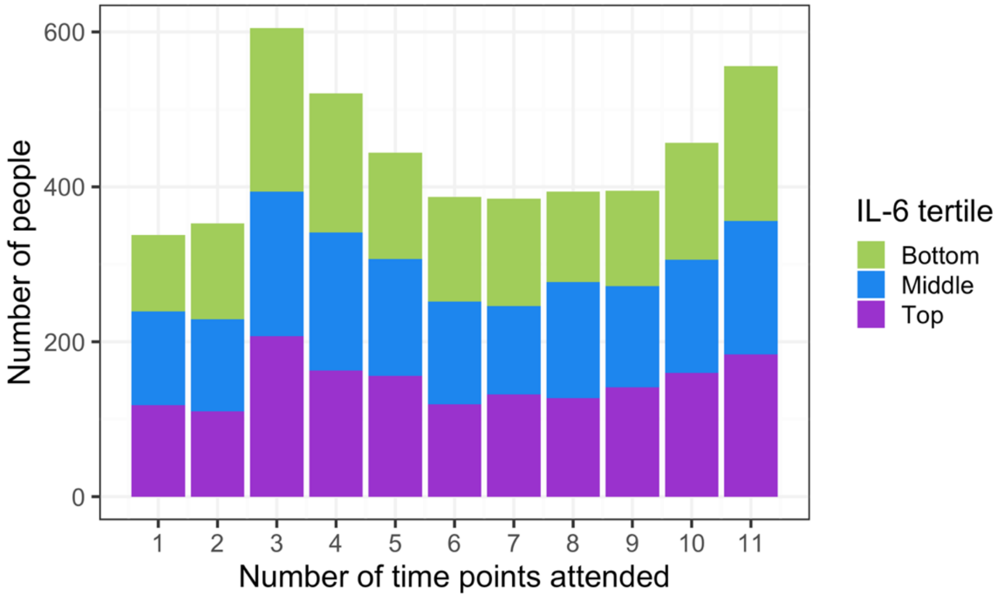


**Supplementary Figure 7. Number of completed PHQ-2 questionnaires in UKB.** Coloured by IL-6 tertile. Participants subset to those that remained alive after the initial assessment. Excluding the two imaging appointments as only a subset of participants were invited to these appointments.

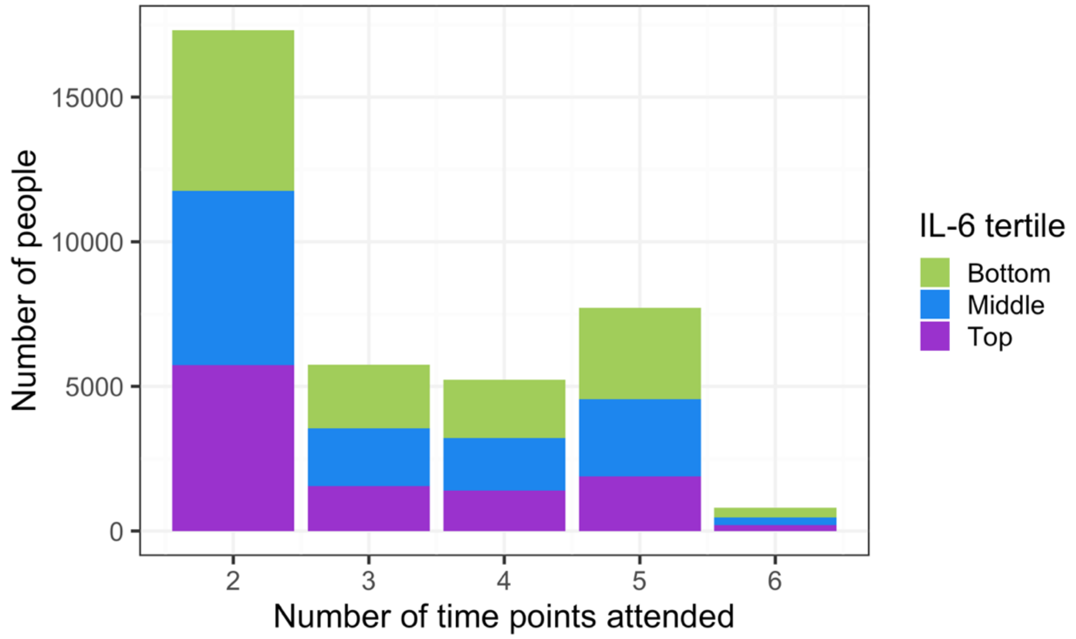


Supplementary Figure 8. Number of completed PHQ-2 questionnaires and if person was dead or alive in UK Biobank.

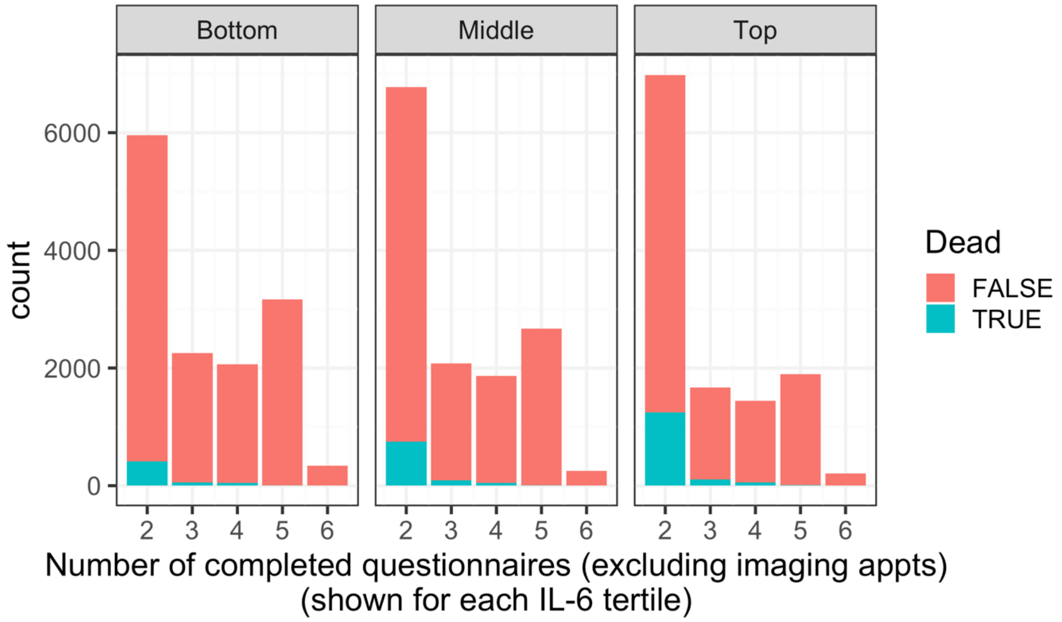

Supplementary Tables

Supplementary Table 1. Descriptive statistics of SMFQ responses and ages at each time point in ALSPAC

| Time Point | dep_mean | dep_SD | dep_min | dep_max | dep_IQR | dep_N | dep_N_missing | age_mean | age_SD | age_min | age_max | age_IQR |
|---|---|---|---|---|---|---|---|---|---|---|---|---|
| 1 | 3.98 | 3.459 | 0 | 23 | 5 | 4458 | 377 | 10.629 | 0.241 | 9.833 | 12.25 | 0.25 |
| 2 | 3.92 | 3.798 | 0 | 25 | 4 | 4052 | 783 | 12.803 | 0.215 | 11.75 | 14.083 | 0.25 |
| 3 | 4.857 | 4.417 | 0 | 26 | 5 | 3674 | 1161 | 13.826 | 0.202 | 12.5 | 15.167 | 0.167 |
| 4 | 5.746 | 5.54 | 0 | 26 | 6 | 2732 | 2103 | 16.683 | 0.237 | 16.417 | 17.833 | 0.333 |
| 5 | 6.473 | 5.25 | 0 | 26 | 6 | 2585 | 2250 | 17.821 | 0.385 | 16.463 | 19.822 | 0.37 |
| 6 | 6.561 | 5.753 | 0 | 26 | 7 | 1831 | 3004 | 18.658 | 0.488 | 17.917 | 20 | 0.833 |
| 7 | 5.35 | 5.19 | 0 | 26 | 7 | 1818 | 3017 | 21.959 | 0.517 | 20.917 | 23.25 | 0.917 |
| 8 | 5.934 | 5.327 | 0 | 26 | 6 | 2062 | 2773 | 22.893 | 0.512 | 21.895 | 24.534 | 0.812 |
| 9 | 6.582 | 5.779 | 0 | 26 | 8 | 2117 | 2718 | 23.872 | 0.507 | 22.917 | 25.25 | 0.75 |
| 10 | 6.532 | 6.209 | 0 | 26 | 8 | 2081 | 2754 | 25.768 | 0.505 | 24.667 | 27.083 | 0.833 |
| 11 | 6.492 | 5.982 | 0 | 26 | 7 | 2163 | 2672 | 28.379 | 0.531 | 27 | 29 | 1 |

Supplementary Table 2. Questions in the SMFQ.

| Question Number | SMFQ Question: |
|---|---|
| 1 | I felt miserable or unhappy |
| 2 | I didn't enjoy anything at all |
| 3 | I felt so tired I just sat around and did nothing |
| 4 | I was very restless |
| 5 | I felt I was no good anymore |
| 6 | I cried a lot |
| 7 | I found it hard to think properly or concentrate |
| 8 | I hated myself |
| 9 | I was a bad person |
| 10 | I felt lonely |
| 11 | I thought nobody really loved me |
| 12 | I thought I could never be as good as others |
| 13 | I did everything wrong |

Supplementary Table 3. Descriptive statistics of PHQ-2 responses and ages at each time point in UK Biobank

| Time Point | dep_mean | dep_SD | dep_min | dep_max | dep_IQR | dep_N | dep_N_missing | age_mean | age_SD | age_min | age_max | age_IQR |
|---|---|---|---|---|---|---|---|---|---|---|---|---|
| Initial | 0.569 | 1.092 | 0 | 6 | 1 | 39435 | 178 | 56.64 | 8.102 | 39 | 70 | 13 |
| Repeat | 0.443 | 0.94 | 0 | 6 | 0 | 1653 | 37960 | 61.447 | 7.184 | 44 | 75 | 10 |
| Imaging | 0.427 | 0.958 | 0 | 6 | 0 | 4760 | 34853 | 64.769 | 7.829 | 45 | 84 | 12 |
| Repeat Imaging | 0.387 | 0.868 | 0 | 6 | 0 | 331 | 39282 | 65.147 | 7.336 | 49 | 81 | 11 |
| Mental health | 0.524 | 1.07 | 0 | 6 | 1 | 12611 | 27002 | 64.016 | 7.775 | 46 | 80 | 12 |
| Pain | 0.592 | 1.119 | 0 | 6 | 1 | 13263 | 26350 | 66.761 | 7.72 | 49 | 82 | 12 |
| Health & well-being | 0.493 | 1.074 | 0 | 6 | 0 | 15502 | 24111 | 69.425 | 7.619 | 52 | 85 | 11 |
| Mental well-being | 0.564 | 1.125 | 0 | 6 | 1 | 13717 | 25896 | 69.591 | 7.538 | 52 | 86 | 12 |

**Supplementary Table 4. PHQ-9 items available at each time point.** Data available is shown as "T" for "TRUE" and data not available is shown as "F" for "FALSE" in the table.

| PHQ-9: | In-person assessment | | | | Online-follow up quesionnaire | | | |
|---|---|---|---|---|---|---|---|---|
| Over the last 2 weeks, how often have you been bothered by any of the following problems? | Initial | First repeat | Imaging | Repeat imaging | Mental Health | Pain | Health and well-being | Mental well-being |
| Little interest or pleasure in doing things | T | T | T | T | T | T | T | T |
| Feeling down, depressed or hopeless | T | T | T | T | T | T | T | T |
| Trouble falling or staying asleep, or sleeping too much | F | F | F | F | T | T | F | T |
| Feeling tired or having little energy | T | T | T | T | T | T | F | T |
| Poor appetite or overeating | F | F | F | F | T | T | F | T |
| Feeling bad about yourself or that you are a failure or have let yourself or your family down | F | F | F | F | T | T | F | T |
| Trouble concentrating on things, such as reading the newspaper or watching television | F | F | F | F | T | T | F | T |
| Being so restless that it is hard to sit still | T | T | T | T | T | F | F | T |
| Thoughts that you would be better off dead or of hurting yourself in some way | F | F | F | F | T | T | F | T |

Supplementary Table 5. Estimates of linear, quadratic, cubic and quartic models to assess best model fit in ALSPAC

| Parameter | Estimate (Linear) | SE (Linear) | p-value (Linear) | N (Linear) | Estimate (Quadratic) | SE (Quadratic) | p-value (Quadratic) | N (Quadratic) | Estimate (Cubic) | SE (Cubic) | p-value (Cubic) | N (Cubic) | Estimate (Quartic) | SE (Quartic) | p-value (Quartic) | N (Quartic) |
|---|---|---|---|---|---|---|---|---|---|---|---|---|---|---|---|---|
| Intercept | 5.4271 | 0.0523 | <0.0001 | 4835 | 5.8704 | 0.068 | <0.0001 | 4835 | 5.8866 | 0.0684 | <0.0001 | 4835 | 6.1495 | 0.0745 | <0.0001 | 4835 |
| age | 0.1603 | 0.0062 | <0.0001 | 4835 | 0.1844 | 0.0064 | <0.0001 | 4835 | 0.1623 | 0.0107 | <0.0001 | 4835 | 0.2311 | 0.0128 | <0.0001 | 4835 |
| age^2 (acceleration) | - | - | - | - | <0.0001 | 0.001 | <0.0001 | 4835 | <0.0001 | 0.0011 | <0.0001 | 4835 | <0.0001 | 0.0027 | <0.0001 | 4835 |
| age^3 (cubic change) | - | - | - | - | - | - | - | - | 0.0004 | 0.0002 | 0.0097 | 4835 | <0.0001 | 0.0003 | <0.0001 | 4835 |
| age^4 (quartic change) | - | - | - | - | - | - | - | - | - | - | - | - | 0.0003 | <0.0001 | <0.0001 | 4835 |
| Intercept variance | 9.104 | 3.0173 | - | - | 14.0229 | 3.7447 | - | - | 14.0559 | 3.7491 | - | - | 14.6395 | 3.8262 | - | - |
| Age (slope) variance | 0.0641 | 0.2532 | - | - | 0.0724 | 0.269 | - | - | 0.0726 | 0.2694 | - | - | 0.0749 | 0.2736 | - | - |
| Intercept/age covariance | 0.5271 | 0.69 | - | - | 0.7067 | 0.7015 | - | - | 0.7079 | 0.7008 | - | - | 0.7364 | 0.7033 | - | - |
| Quadratic variance | - | - | - | - | 0.0009 | 0.0304 | - | - | 0.0009 | 0.0305 | - | - | 0.001 | 0.0317 | - | - |
| Intercept/Quadratic covariance | - | - | - | - | -0.0907 | -0.7958 | - | - | -0.0911 | -0.7959 | - | - | -0.0971 | -0.8001 | - | - |
| Age (slope)/Quadratic covariance | - | - | - | - | -0.0022 | -0.2731 | - | - | -0.0023 | -0.2739 | - | - | -0.0026 | -0.2942 | - | - |
| Residual variance | 14.4745 | 3.8045 | - | - | 13.5098 | 3.6756 | - | - | 13.4971 | 3.6738 | - | - | 13.3474 | 3.6534 | - | - |
| Deviance | 171265.9 | | | | 170363.493 | | | | 170356.8 | | | | 170266 | | | |
| AIC | 171277.9 | | | | 170383.493 | | | | 170378.8 | | | | 170290 | | | |
| BIC | 171327.6 | | | | 170466.432 | | | | 170470.1 | | | | 170389.5 | | | |

Supplementary Table 6. Estimates of linear and quadratic models to assess best model fit in UK Biobank

| Parameter | Estimate (Linear) | SE (Linear) | p-value (Linear) | N (Linear) | Estimate (Quadratic) | SE (Quadratic) | p-value (Quadratic) | N (Quadratic) |
|---|---|---|---|---|---|---|---|---|
| Intercept | 0.5624 | 0.0048 | <0.0001 | 39613 | 0.5464 | 0.0053 | <0.0001 | 39613 |
| age | <0.0001 | 0.0004 | <0.0001 | 39613 | <0.0001 | 0.0004 | <0.0001 | 39613 |
| age^2 (acceleration) | - | - | - | - | 0.0002 | <0.0001 | <0.0001 | 39613 |
| Intercept variance | 0.5191 | 0.7205 | - | - | 0.5202 | 0.7212 | - | - |
| Age (slope) variance | 0.0011 | 0.0335 | - | - | 0.0011 | 0.0334 | - | - |
| Intercept/age covariance | -0.003 | -0.1259 | - | - | -0.0028 | -0.1148 | - | - |
| Residual variance | 0.5897 | 0.7679 | - | - | 0.5892 | 0.7676 | - | - |
| Deviance | 283639.0953 | | | | 283584.1488 | | | |
| AIC | 283651.0953 | | | | 283598.1488 | | | |
| BIC | 283708.2487 | | | | 283664.8278 | | | |

Supplementary Table 7. Counts of each anti-inflammatory medication taken in UK Biobank.

| Medication | Count |
|---|---|
| Aspirin 75mg Tablet | 239 |
| Nu-Seals Aspirin 75mg E/C Tablet | 0 |
| Aspirin | 5316 |
| Aspirin+Methocarbamol 325mg/400mg Tablet | 0 |
| Ibuprofen | 0 |
| Indomethacin | 0 |
| Naproxen | 0 |
| Mefenamic Acid | 0 |
| Aspirin+Metoclopramide 325mg/5mg Effervescent Tablet | 0 |
| Ibuprofen+Codeine Phosphate | 0 |
| Diclofenac Sodium+Misoprostol | 0 |
| Naproxen+Misoprostol | 4 |
| Aspirin+Cyclizine Hydrochloride 500mg/25mg Tablet | 4799 |
| Aspirin+Glycine 500mg/133mg Dispersible Tablet | 1 |
| Aspirin+Codeine 300mg/8mg Tablet | 0 |
| Aspirin+Codeine | 5 |
| Diclofenac | 3 |
| Propionic Acid-Ibuprofen | 1 |
| Ibuprofen+Menthol 5%/3% Gel | 9 |
| Deep Relief Ibuprofen Gel | 0 |
| Anadin Ibuprofen 200mg Tablet | 0 |
| Ibuprofen Product | 4 |
| Indomethacin Product | 240 |
| Aspirin+Papaveretum 500mg/7.71mg Dispersible Tablet | 0 |
| Isosorbide Mononitrate+Aspirin | 19 |
| Dipyridamole+Aspirin | 779 |
| Celecoxib | 35 |
| Etoricoxib | 42 |
| Ibuprofen+Pseudoephedrine Hydrochloride | 24 |
| Lemsip Flu 12hr Ibuprofen+Pseudoephedrine Capsule | 23 |
| Care Ibuprofen 10% Gel | 1 |

Supplementary Table 8. Counts and types of inflammatory conditions in UK Biobank.

| Condition | N |
|---|---|
| Diabetes | 1652 |
| Eczema/Dermatitis | 1002 |
| Gout | 515 |
| Emphysema/Chronic Bronchitis | 482 |
| Psoriasis | 428 |
| Rheumatoid Arthritis | 386 |
| Spine Arthritis/Spondylitis | 304 |
| Arthritis | 299 |
| Type 2 Diabetes | 257 |
| Chronic Sinusitis | 212 |
| Tuberculosis | 203 |
| Ulcerative Colitis | 203 |
| Crohns Disease | 131 |
| Multiple Sclerosis | 130 |
| Helicobacter Pylori | 120 |
| Bronchiectasis | 96 |
| Ankylosing Spondylitis | 79 |
| Non-Hodgkins Lymphoma | 70 |
| Polymyalgia Rheumatica | 66 |
| Psoriatic Arthropathy | 57 |
| Non-Infective Hepatitis | 44 |
| Hodgkins Lymphoma/Hodgkins Disease | 37 |
| Hepatitis | 35 |
| Sjogren's Syndrome/Sicca Syndrome | 32 |
| HIV/AIDS | 31 |
| Type 1 Diabetes | 31 |
| Systemic Lupus Erythematosis/Sle | 28 |
| Polio/Poliomyelitis | 21 |
| Lichen Sclerosis | 19 |
| Acute Infective Polyneuritis/Guillain/Barre Syndrome | 18 |
| Vasculitis | 17 |
| Leukaemia | 16 |
| Hepatitis C | 13 |
| Scleroderma/Systemic Sclerosis | 13 |
| Connective Tissue Disorder | 12 |
| Hepatitis B | 12 |
| Nephritis | 12 |
| Plantar Fascitis | 12 |
| Optic Neuritis | 11 |
| Fibrosing Alveolitis/Unspecified Alveolitis | 9 |
| Giant Cell/Temporal Arteritis | 9 |
| Inflammatory Bowel Disease | 9 |
| Myositis/Myopathy | 8 |
| Wegners Granulmatosis | 8 |
| Pemphigoid/Pemphigus | 7 |
| Primary Biliary Cirrhosis | 7 |
| Lymphoma | 6 |
| Pelvic Inflammatory Disease | 6 |
| Polymyositis | 5 |
| Acute Myeloid Leukaemia | 3 |
| Sclerosing Cholangitis | 3 |
| Dermatomyositis | 2 |
| Glomerulnephritis | 2 |
| Microscopic Polyarteritis | 1 |
| None | 33271 |

Supplementary Table 9. Estimated depression scores for each IL-6 tertile trajectory at ages 10, 13, 16, 19, 22, 25 and 28 years, in ALSPAC.

| IL-6 Tertile Group | age | estimate | 95% CI |
|---|---|---|---|
| SMFQ Score [IL-6 tertile = Bottom ] | Age 10 | 1.207 | -0.702 - 3.116 |
| SMFQ Score [IL-6 tertile = Middle ] | Age 10 | 1.079 | -0.86 - 3.018 |
| SMFQ Score [IL-6 tertile = Top ] | Age 10 | 0.993 | -0.974 - 2.961 |
| SMFQ Score [IL-6 tertile = Bottom ] | Age 13 | 1.491 | -0.362 - 3.345 |
| SMFQ Score [IL-6 tertile = Middle ] | Age 13 | 1.524 | -0.357 - 3.405 |
| SMFQ Score [IL-6 tertile = Top ] | Age 13 | 1.926 | 0.018 - 3.835 |
| SMFQ Score [IL-6 tertile = Bottom ] | Age 16 | 2.550 | 0.681 - 4.419 |
| SMFQ Score [IL-6 tertile = Middle ] | Age 16 | 2.667 | 0.77 - 4.564 |
| SMFQ Score [IL-6 tertile = Top ] | Age 16 | 3.148 | 1.223 - 5.074 |
| SMFQ Score [IL-6 tertile = Bottom ] | Age 19 | 3.339 | 1.445 - 5.233 |
| SMFQ Score [IL-6 tertile = Middle ] | Age 19 | 3.452 | 1.53 - 5.374 |
| SMFQ Score [IL-6 tertile = Top ] | Age 19 | 3.887 | 1.936 - 5.838 |
| SMFQ Score [IL-6 tertile = Bottom ] | Age 22 | 3.482 | 1.578 - 5.387 |
| SMFQ Score [IL-6 tertile = Middle ] | Age 22 | 3.494 | 1.561 - 5.427 |
| SMFQ Score [IL-6 tertile = Top ] | Age 22 | 3.924 | 1.961 - 5.886 |
| SMFQ Score [IL-6 tertile = Bottom ] | Age 25 | 3.271 | 1.35 - 5.192 |
| SMFQ Score [IL-6 tertile = Middle ] | Age 25 | 3.074 | 1.125 - 5.022 |
| SMFQ Score [IL-6 tertile = Top ] | Age 25 | 3.593 | 1.614 - 5.571 |
| SMFQ Score [IL-6 tertile = Bottom ] | Age 28 | 3.662 | 1.706 - 5.618 |
| SMFQ Score [IL-6 tertile = Middle ] | Age 28 | 3.139 | 1.156 - 5.122 |
| SMFQ Score [IL-6 tertile = Top ] | Age 28 | 3.782 | 1.769 - 5.796 |

Supplementary Table 10. Model estimates for main analysis in ALSPAC. IL-6 as a categorical variable (tertiles).

| | Unadjusted | | | | Sex Adjusted | | | | Fully Adjusted | | | |
|---|---|---|---|---|---|---|---|---|---|---|---|---|
| Parameter | Estimate | SE | p-value | N | Estimate | SE | p-value | N | Estimate | SE | p-value | N |
| Intercept | 5.8216 | 0.1303 | <0.0001 | 4835 | 5.4732 | 0.1331 | <0.0001 | 4835 | 3.1255 | 0.9109 | 0.0006 | 4264 |
| age | 0.2389 | 0.0221 | <0.0001 | 4835 | 0.2369 | 0.0221 | <0.0001 | 4835 | 0.2389 | 0.023 | <0.0001 | 4264 |
| age^2 (acceleration) | <0.0001 | 0.0047 | <0.0001 | 4835 | <0.0001 | 0.0047 | <0.0001 | 4835 | <0.0001 | 0.0049 | <0.0001 | 4264 |
| age^3 (cubic change) | <0.0001 | 0.0004 | 0.0001 | 4835 | <0.0001 | 0.0004 | 0.0001 | 4835 | <0.0001 | 0.0004 | 0.0001 | 4264 |
| age^4 (quartic change) | 0.0003 | <0.0001 | <0.0001 | 4835 | 0.0003 | <0.0001 | <0.0001 | 4835 | 0.0003 | <0.0001 | <0.0001 | 4264 |
| IL6_tertileMiddle | 0.1953 | 0.1848 | 0.2905 | 4835 | 0.1341 | 0.1811 | 0.459 | 4835 | 0.125 | 0.185 | 0.4994 | 4264 |
| IL6_tertileTop | 0.7869 | 0.1841 | <0.0001 | 4835 | 0.655 | 0.1809 | 0.0003 | 4835 | 0.5781 | 0.1874 | 0.002 | 4264 |
| age:IL6_tertileMiddle | <0.0001 | 0.0314 | 0.7965 | 4835 | <0.0001 | 0.0314 | 0.8218 | 4835 | <0.0001 | 0.0327 | 0.8672 | 4264 |
| age:IL6_tertileTop | <0.0001 | 0.0313 | 0.5888 | 4835 | <0.0001 | 0.0313 | 0.5887 | 4835 | <0.0001 | 0.0328 | 0.4683 | 4264 |
| age^2 (acceleration):IL6_tertileMiddle | <0.0001 | 0.0066 | 0.3732 | 4835 | <0.0001 | 0.0066 | 0.3658 | 4835 | <0.0001 | 0.0069 | 0.45 | 4264 |
| age^2 (acceleration):IL6_tertileTop | <0.0001 | 0.0066 | 0.4042 | 4835 | <0.0001 | 0.0066 | 0.4028 | 4835 | <0.0001 | 0.0069 | 0.4695 | 4264 |
| age^3 (cubic change):IL6_tertileMiddle | <0.0001 | 0.0006 | 0.8107 | 4835 | <0.0001 | 0.0006 | 0.7934 | 4835 | <0.0001 | 0.0006 | 0.9216 | 4264 |
| age^3 (cubic change):IL6_tertileTop | 0.0007 | 0.0006 | 0.2268 | 4835 | 0.0007 | 0.0006 | 0.2205 | 4835 | 0.0009 | 0.0006 | 0.1731 | 4264 |
| age^4 (quartic change):IL6_tertileMiddle | <0.0001 | <0.0001 | 0.8464 | 4835 | <0.0001 | <0.0001 | 0.8377 | 4835 | <0.0001 | <0.0001 | 0.9983 | 4264 |
| age^4 (quartic change):IL6_tertileTop | <0.0001 | <0.0001 | 0.6014 | 4835 | <0.0001 | <0.0001 | 0.5959 | 4835 | <0.0001 | <0.0001 | 0.5005 | 4264 |
| Sex1 | - | - | - | - | 0.791 | 0.0866 | <0.0001 | 4835 | 0.762 | 0.0904 | <0.0001 | 4264 |
| Maternal.education.at.birth1 | - | - | - | - | - | - | - | - | <0.0001 | 0.0895 | 0.2825 | 4264 |
| BMI_age9_log | - | - | - | - | - | - | - | - | 0.8442 | 0.3204 | 0.0084 | 4264 |
| Intercept variance | 15.2155 | 3.9007 | - | - | 14.1944 | 3.7675 | - | - | 12.9107 | 3.5932 | - | - |
| Age (slope) variance | 0.0764 | 0.2764 | - | - | 0.0753 | 0.2745 | - | - | 0.0719 | 0.2682 | - | - |
| Quadratic variance | 0.0011 | 0.0329 | - | - | 0.001 | 0.0321 | - | - | 0.001 | 0.031 | - | - |
| Intercept/age covariance | 0.7653 | 0.7099 | - | - | 0.7251 | 0.7012 | - | - | 0.6619 | 0.6868 | - | - |
| Intercept/Quadratic covariance | -0.1032 | -0.8048 | - | - | -0.0957 | -0.7927 | - | - | -0.0854 | -0.7664 | - | - |

| | Unadjusted | | | | Sex Adjusted | | | | Fully Adjusted | | | |
|---|---|---|---|---|---|---|---|---|---|---|---|---|
| Parameter | Estimate | SE | p-value | N | Estimate | SE | p-value | N | Estimate | SE | p-value | N |
| Age (slope)/Quadratic covariance | -0.0029 | -0.3198 | - | - | -0.0027 | -0.3093 | - | - | -0.0023 | -0.2732 | - | - |
| Residual variance | 13.251 | 3.6402 | - | - | 13.3118 | 3.6485 | - | - | 13.2588 | 3.6413 | - | - |
| Deviance | 170235.3316 | | | | 170144.0111 | | | | 152823.3672 | | | |
| AIC | 170279.3316 | | | | 170190.0111 | | | | 152873.3672 | | | |
| BIC | 170461.7982 | | | | 170380.7717 | | | | 153078.0876 | | | |

Supplementary Table 11. Model estimates for main analysis in ALSPAC split by sex. IL-6 as a categorical variable assigned to female and males separately.

| | Unadjusted | | | | Fully Adjusted | | | |
|---|---|---|---|---|---|---|---|---|
| Parameter | Estimate | SE | p-value | N | Estimate | SE | p-value | N |
| Intercept | 5.0663 | 0.1718 | <0.0001 | 4835 | 2.838 | 0.9237 | 0.0021 | 4264 |
| age | 0.2454 | 0.0305 | <0.0001 | 4835 | 0.2567 | 0.0316 | <0.0001 | 4264 |
| age^2 (acceleration) | <0.0001 | 0.0067 | 0.0002 | 4835 | <0.0001 | 0.007 | 0.0003 | 4264 |
| age^3 (cubic change) | <0.0001 | 0.0006 | <0.0001 | 4835 | <0.0001 | 0.0006 | <0.0001 | 4264 |
| age^4 (quartic change) | 0.0003 | <0.0001 | <0.0001 | 4835 | 0.0003 | <0.0001 | <0.0001 | 4264 |
| IL6_sex_tertileMale_Middle | <0.0001 | 0.2539 | 0.5215 | 4835 | <0.0001 | 0.2656 | 0.5684 | 4264 |
| IL6_sex_tertileMale_Top | 0.142 | 0.2699 | 0.5989 | 4835 | 0.1456 | 0.2836 | 0.6077 | 4264 |
| IL6_sex_tertileFemale_Bottom | 1.6241 | 0.2492 | <0.0001 | 4835 | 1.5583 | 0.2616 | <0.0001 | 4264 |
| IL6_sex_tertileFemale_Middle | 1.9246 | 0.2406 | <0.0001 | 4835 | 1.8389 | 0.2525 | <0.0001 | 4264 |
| IL6_sex_tertileFemale_Top | 2.3796 | 0.2313 | <0.0001 | 4835 | 2.2044 | 0.2462 | <0.0001 | 4264 |
| age:IL6_sex_tertileMale_Middle | 0.0469 | 0.0455 | 0.3026 | 4835 | 0.0334 | 0.0472 | 0.4791 | 4264 |
| age:IL6_sex_tertileMale_Top | 0.0247 | 0.0484 | 0.6094 | 4835 | 0.013 | 0.0502 | 0.7959 | 4264 |
| age:IL6_sex_tertileFemale_Bottom | <0.0001 | 0.044 | 0.4923 | 4835 | <0.0001 | 0.0458 | 0.2717 | 4264 |
| age:IL6_sex_tertileFemale_Middle | <0.0001 | 0.0426 | 0.0714 | 4835 | <0.0001 | 0.0443 | 0.0695 | 4264 |
| age:IL6_sex_tertileFemale_Top | <0.0001 | 0.041 | 0.1359 | 4835 | <0.0001 | 0.0429 | 0.0615 | 4264 |
| age^2 (acceleration):IL6_sex_tertileMale_Middle | <0.0001 | 0.01 | 0.6906 | 4835 | <0.0001 | 0.0103 | 0.5627 | 4264 |
| age^2 (acceleration):IL6_sex_tertileMale_Top | 0.0055 | 0.0106 | 0.6023 | 4835 | 0.0069 | 0.011 | 0.53 | 4264 |
| age^2 (acceleration):IL6_sex_tertileFemale_Bottom | <0.0001 | 0.0094 | 0.0343 | 4835 | <0.0001 | 0.0097 | 0.0484 | 4264 |
| age^2 (acceleration):IL6_sex_tertileFemale_Middle | <0.0001 | 0.0091 | 0.0093 | 4835 | <0.0001 | 0.0094 | 0.0258 | 4264 |

| | Unadjusted | | | | Fully Adjusted | | | |
|---|---|---|---|---|---|---|---|---|
| Parameter | Estimate | SE | p-value | N | Estimate | SE | p-value | N |
| age^2 (acceleration):IL6_sex_tertileFemale_Top | <0.0001 | 0.0088 | 0.0041 | 4835 | <0.0001 | 0.0092 | 0.0062 | 4264 |
| age^3 (cubic change):IL6_sex_tertileMale_Middle | <0.0001 | 0.0009 | 0.0986 | 4835 | <0.0001 | 0.0009 | 0.2237 | 4264 |
| age^3 (cubic change):IL6_sex_tertileMale_Top | <0.0001 | 0.0009 | 0.8366 | 4835 | 0.0002 | 0.001 | 0.866 | 4264 |
| age^3 (cubic change):IL6_sex_tertileFemale_Bottom | 0.0028 | 0.0009 | 0.0013 | 4835 | 0.0031 | 0.0009 | 0.0005 | 4264 |
| age^3 (cubic change):IL6_sex_tertileFemale_Middle | 0.0033 | 0.0008 | <0.0001 | 4835 | 0.0034 | 0.0009 | <0.0001 | 4264 |
| age^3 (cubic change):IL6_sex_tertileFemale_Top | 0.0033 | 0.0008 | <0.0001 | 4835 | 0.0036 | 0.0008 | <0.0001 | 4264 |
| age^4 (quartic change):IL6_sex_tertileMale_Middle | <0.0001 | 0.0001 | 0.6102 | 4835 | <0.0001 | 0.0001 | 0.6173 | 4264 |
| age^4 (quartic change):IL6_sex_tertileMale_Top | <0.0001 | 0.0001 | 0.5432 | 4835 | <0.0001 | 0.0001 | 0.3537 | 4264 |
| age^4 (quartic change):IL6_sex_tertileFemale_Bottom | <0.0001 | 0.0001 | 0.6751 | 4835 | <0.0001 | 0.0001 | 0.5402 | 4264 |
| age^4 (quartic change):IL6_sex_tertileFemale_Middle | <0.0001 | 0.0001 | 0.4591 | 4835 | <0.0001 | 0.0001 | 0.3102 | 4264 |
| age^4 (quartic change):IL6_sex_tertileFemale_Top | <0.0001 | 0.0001 | 0.4934 | 4835 | <0.0001 | 0.0001 | 0.4587 | 4264 |
| Maternal.education.at.birth1 | - | - | - | - | <0.0001 | 0.0899 | 0.3411 | 4264 |
| BMI_age9_log | - | - | - | - | 0.8086 | 0.3218 | 0.012 | 4264 |
| Intercept variance | 13.0356 | 3.6105 | - | - | 13.1192 | 3.622 | - | - |
| Age (slope) variance | 0.0702 | 0.265 | - | - | 0.0706 | 0.2658 | - | - |
| Quadratic variance | 0.0009 | 0.0297 | - | - | 0.001 | 0.0309 | - | - |
| Intercept/age covariance | 0.6543 | 0.6839 | - | - | 0.6572 | 0.6827 | - | - |
| Intercept/Quadratic covariance | -0.0832 | -0.7755 | - | - | -0.0863 | -0.7709 | - | - |
| Age (slope)/Quadratic covariance | -0.0019 | -0.2393 | - | - | -0.0021 | -0.2567 | - | - |
| Residual variance | 13.3568 | 3.6547 | - | - | 13.1276 | 3.6232 | - | - |
| Deviance | 169910.5598 | | | | 152628.5789 | | | |
| AIC | 169984.5598 | | | | 152706.5789 | | | |
| BIC | 170291.4356 | | | | 153025.9428 | | | |

Supplementary Table 12. Estimated depression scores for each IL-6 tertile trajectory at ages 10, 13, 16, 19, 22, 25 and 28 years, in ALSPAC, split by sex.

| IL-6 Tertile Group | age | estimate | 95% CI |
|---|---|---|---|
| SMFQ Score [IL6_sex_tertile, level = Male_Bottom ] | Age 10 | 2.062 | -0.003 - 4.128 |
| SMFQ Score [IL6_sex_tertile, level = Male_Middle ] | Age 10 | 2.067 | -0.036 - 4.17 |
| SMFQ Score [IL6_sex_tertile, level = Male_Top ] | Age 10 | 1.964 | -0.17 - 4.098 |
| SMFQ Score [IL6_sex_tertile, level = Female_Bottom ] | Age 10 | 0.956 | -1.125 - 3.038 |
| SMFQ Score [IL6_sex_tertile, level = Female_Middle ] | Age 10 | 1.034 | -1.074 - 3.141 |
| SMFQ Score [IL6_sex_tertile, level = Female_Top ] | Age 10 | 1.212 | -0.924 - 3.348 |
| SMFQ Score [IL6_sex_tertile, level = Male_Bottom ] | Age 13 | 1.517 | -0.438 - 3.472 |
| SMFQ Score [IL6_sex_tertile, level = Male_Middle ] | Age 13 | 1.224 | -0.761 - 3.21 |
| SMFQ Score [IL6_sex_tertile, level = Male_Top ] | Age 13 | 1.673 | -0.334 - 3.681 |
| SMFQ Score [IL6_sex_tertile, level = Female_Bottom ] | Age 13 | 2.439 | 0.477 - 4.4 |
| SMFQ Score [IL6_sex_tertile, level = Female_Middle ] | Age 13 | 2.761 | 0.772 - 4.749 |
| SMFQ Score [IL6_sex_tertile, level = Female_Top ] | Age 13 | 3.031 | 1.009 - 5.054 |
| SMFQ Score [IL6_sex_tertile, level = Male_Bottom ] | Age 16 | 2.275 | 0.288 - 4.263 |
| SMFQ Score [IL6_sex_tertile, level = Male_Middle ] | Age 16 | 2.046 | 0.026 - 4.066 |
| SMFQ Score [IL6_sex_tertile, level = Male_Top ] | Age 16 | 2.419 | 0.374 - 4.463 |
| SMFQ Score [IL6_sex_tertile, level = Female_Bottom ] | Age 16 | 3.836 | 1.841 - 5.831 |
| SMFQ Score [IL6_sex_tertile, level = Female_Middle ] | Age 16 | 4.165 | 2.145 - 6.185 |
| SMFQ Score [IL6_sex_tertile, level = Female_Top ] | Age 16 | 4.515 | 2.46 - 6.569 |
| SMFQ Score [IL6_sex_tertile, level = Male_Bottom ] | Age 19 | 3.081 | 1.04 - 5.123 |
| SMFQ Score [IL6_sex_tertile, level = Male_Middle ] | Age 19 | 2.957 | 0.88 - 5.035 |
| SMFQ Score [IL6_sex_tertile, level = Male_Top ] | Age 19 | 3.249 | 1.141 - 5.356 |
| SMFQ Score [IL6_sex_tertile, level = Female_Bottom ] | Age 19 | 4.567 | 2.519 - 6.615 |
| SMFQ Score [IL6_sex_tertile, level = Female_Middle ] | Age 19 | 4.814 | 2.742 - 6.885 |
| SMFQ Score [IL6_sex_tertile, level = Female_Top ] | Age 19 | 5.175 | 3.07 - 7.28 |
| SMFQ Score [IL6_sex_tertile, level = Male_Bottom ] | Age 22 | 3.356 | 1.301 - 5.411 |
| SMFQ Score [IL6_sex_tertile, level = Male_Middle ] | Age 22 | 3.185 | 1.092 - 5.277 |
| SMFQ Score [IL6_sex_tertile, level = Male_Top ] | Age 22 | 3.646 | 1.521 - 5.77 |
| SMFQ Score [IL6_sex_tertile, level = Female_Bottom ] | Age 22 | 4.581 | 2.521 - 6.641 |
| SMFQ Score [IL6_sex_tertile, level = Female_Middle ] | Age 22 | 4.718 | 2.635 - 6.801 |
| SMFQ Score [IL6_sex_tertile, level = Female_Top ] | Age 22 | 5.035 | 2.919 - 7.152 |
| SMFQ Score [IL6_sex_tertile, level = Male_Bottom ] | Age 25 | 3.197 | 1.099 - 5.296 |
| SMFQ Score [IL6_sex_tertile, level = Male_Middle ] | Age 25 | 2.753 | 0.612 - 4.894 |
| SMFQ Score [IL6_sex_tertile, level = Male_Top ] | Age 25 | 3.524 | 1.347 - 5.7 |
| SMFQ Score [IL6_sex_tertile, level = Female_Bottom ] | Age 25 | 4.357 | 2.257 - 6.456 |
| SMFQ Score [IL6_sex_tertile, level = Female_Middle ] | Age 25 | 4.332 | 2.211 - 6.452 |
| SMFQ Score [IL6_sex_tertile, level = Female_Top ] | Age 25 | 4.631 | 2.477 - 6.785 |
| SMFQ Score [IL6_sex_tertile, level = Male_Bottom ] | Age 28 | 3.378 | 1.208 - 5.547 |
| SMFQ Score [IL6_sex_tertile, level = Male_Middle ] | Age 28 | 2.487 | 0.267 - 4.708 |
| SMFQ Score [IL6_sex_tertile, level = Male_Top ] | Age 28 | 3.231 | 0.97 - 5.491 |
| SMFQ Score [IL6_sex_tertile, level = Female_Bottom ] | Age 28 | 4.905 | 2.743 - 7.067 |
| SMFQ Score [IL6_sex_tertile, level = Female_Middle ] | Age 28 | 4.551 | 2.372 - 6.73 |
| SMFQ Score [IL6_sex_tertile, level = Female_Top ] | Age 28 | 5.007 | 2.796 - 7.217 |

Supplementary Table 13. Model estimates for sensitivity analysis in ALSPAC. IL-6 as a continuous variable.

| | Unadjusted | | | Sex Adjusted | | | Fully Adjusted | | |
|---|---|---|---|---|---|---|---|---|---|
| Parameter | Estimate | SE | p-value | Estimate | SE | p-value | Estimate | SE | p-value |
| Intercept | 6.1499 | 0.0733 | <0.0001 | 5.7562 | 0.0892 | <0.0001 | 3.2696 | 0.9165 | 4e-04 |
| age | 0.2317 | 0.0128 | <0.0001 | 0.2291 | 0.0128 | <0.0001 | 0.2293 | 0.0134 | <0.0001 |
| age^2 (acceleration) | <0.0001 | 0.0027 | <0.0001 | <0.0001 | 0.0027 | <0.0001 | <0.0001 | 0.0028 | <0.0001 |
| age^3 (cubic change) | <0.0001 | 3e-04 | <0.0001 | <0.0001 | 2e-04 | <0.0001 | <0.0001 | 3e-04 | <0.0001 |
| age^4 (quartic change) | 3e-04 | <0.0001 | <0.0001 | 3e-04 | <0.0001 | <0.0001 | 3e-04 | <0.0001 | <0.0001 |
| IL6_log | 0.2998 | 0.0733 | <0.0001 | 0.2515 | 0.0769 | 0.0011 | 0.2073 | 0.0782 | 0.008 |
| age:IL6_log | <0.0001 | 0.0128 | 0.3912 | <0.0001 | 0.0128 | 0.3723 | <0.0001 | 0.0134 | 0.14 |
| age^2 (acceleration):IL6_log | <0.0001 | 0.0027 | 0.4018 | <0.0001 | 0.0027 | 0.4119 | <0.0001 | 0.0028 | 0.4358 |
| age^3 (cubic change):IL6_log | 4e-04 | 3e-04 | 0.1192 | 4e-04 | 2e-04 | 0.1042 | 5e-04 | 3e-04 | 0.0437 |
| age^4 (quartic change):IL6_log | <0.0001 | <0.0001 | 0.7535 | <0.0001 | <0.0001 | 0.7461 | <0.0001 | <0.0001 | 0.6743 |
| Sex1 | - | - | - | 0.7552 | 0.0878 | <0.0001 | 0.7423 | 0.0912 | <0.0001 |
| Maternal.education.at.birth1 | - | - | - | - | - | - | <0.0001 | 0.0905 | 0.3209 |
| BMI_age9_log | - | - | - | - | - | - | 0.8782 | 0.3198 | 0.006 |
| Intercept variance | 13.9376 | 3.7333 | - | 16.1267 | 4.0158 | - | 13.9625 | 3.7366 | - |
| Age (slope) variance | 0.0726 | 0.2695 | - | 0.0811 | 0.2847 | - | 0.0754 | 0.2747 | - |
| Quadratic variance | 9e-04 | 0.0307 | - | 0.0012 | 0.0353 | - | 0.0011 | 0.0329 | - |
| Intercept/age covariance | 0.7024 | 0.6981 | - | 0.8229 | 0.7196 | - | 0.7111 | 0.6928 | - |
| Intercept/Quadratic covariance | -0.0906 | -0.7895 | - | -0.1151 | -0.811 | - | -0.096 | -0.7808 | - |
| Age (slope)/Quadratic covariance | -0.0022 | -0.2702 | - | -0.0037 | -0.3672 | - | -0.0028 | -0.3067 | - |
| Residual variance | 13.4348 | 3.6654 | - | 13.0761 | 3.6161 | - | 13.0989 | 3.6192 | - |
| Deviance | 170506.3207 | | | 170191.0472 | | | 152836.3256 | | |
| AIC | 170540.3207 | | | 170227.0472 | | | 152876.3256 | | |
| BIC | 170681.3429 | | | 170376.3381 | | | 153040.1019 | | |

Supplementary Table 14. Model estimates for sensitivity analysis in ALSPAC. Individuals taking medication removed.

| | Unadjusted | | | | Sex Adjusted | | | | Fully Adjusted | | | |
|---|---|---|---|---|---|---|---|---|---|---|---|---|
| Parameter | Estimate | SE | p-value | N | Estimate | SE | p-value | N | Estimate | SE | p-value | N |
| Intercept | 5.7724 | 0.1373 | <0.0001 | 4140 | 5.4303 | 0.1472 | <0.0001 | 4140 | 3.4819 | 0.9964 | 0.0005 | 3655 |
| age | 0.2316 | 0.0235 | <0.0001 | 4140 | 0.2293 | 0.0235 | <0.0001 | 4140 | 0.2292 | 0.0244 | <0.0001 | 3655 |
| age^2 (acceleration) | <0.0001 | 0.005 | <0.0001 | 4140 | <0.0001 | 0.005 | <0.0001 | 4140 | <0.0001 | 0.0052 | <0.0001 | 3655 |
| age^3 (cubic change) | <0.0001 | 0.0005 | 0.0007 | 4140 | <0.0001 | 0.0005 | 0.0007 | 4140 | <0.0001 | 0.0005 | 0.0008 | 3655 |
| age^4 (quartic change) | 0.0003 | <0.0001 | <0.0001 | 4140 | 0.0003 | <0.0001 | <0.0001 | 4140 | 0.0004 | <0.0001 | <0.0001 | 3655 |
| IL6_tertileMiddle | 0.2617 | 0.1962 | 0.1824 | 4140 | 0.2002 | 0.2019 | 0.3215 | 4140 | 0.2173 | 0.2004 | 0.2782 | 3655 |
| IL6_tertileTop | 0.9103 | 0.1969 | <0.0001 | 4140 | 0.7727 | 0.2031 | 0.0001 | 4140 | 0.6522 | 0.2046 | 0.0014 | 3655 |
| age:IL6_tertileMiddle | <0.0001 | 0.0337 | 0.8322 | 4140 | <0.0001 | 0.0337 | 0.8629 | 4140 | 0.0031 | 0.0351 | 0.9305 | 3655 |
| age:IL6_tertileTop | <0.0001 | 0.0338 | 0.884 | 4140 | <0.0001 | 0.0338 | 0.8785 | 4140 | <0.0001 | 0.0354 | 0.8389 | 3655 |
| age^2 (acceleration):IL6_tertileMiddle | <0.0001 | 0.0071 | 0.5986 | 4140 | <0.0001 | 0.0071 | 0.5773 | 4140 | <0.0001 | 0.0074 | 0.5932 | 3655 |
| age^2 (acceleration):IL6_tertileTop | <0.0001 | 0.0071 | 0.5828 | 4140 | <0.0001 | 0.0071 | 0.5843 | 4140 | <0.0001 | 0.0075 | 0.6077 | 3655 |
| age^3 (cubic change):IL6_tertileMiddle | <0.0001 | 0.0007 | 0.8908 | 4140 | <0.0001 | 0.0007 | 0.8593 | 4140 | <0.0001 | 0.0007 | 0.8198 | 3655 |
| age^3 (cubic change):IL6_tertileTop | 0.0007 | 0.0007 | 0.3154 | 4140 | 0.0007 | 0.0007 | 0.3059 | 4140 | 0.0006 | 0.0007 | 0.3775 | 3655 |
| age^4 (quartic change):IL6_tertileMiddle | <0.0001 | <0.0001 | 0.9082 | 4140 | <0.0001 | <0.0001 | 0.9361 | 4140 | <0.0001 | <0.0001 | 0.8994 | 3655 |
| age^4 (quartic change):IL6_tertileTop | <0.0001 | <0.0001 | 0.4771 | 4140 | <0.0001 | <0.0001 | 0.4746 | 4140 | <0.0001 | <0.0001 | 0.4972 | 3655 |
| Sex1 | - | - | - | - | 0.7855 | 0.0953 | <0.0001 | 4140 | 0.7863 | 0.0977 | <0.0001 | 3655 |
| Maternal.education.at.birth1 | - | - | - | - | - | - | - | - | <0.0001 | 0.0969 | 0.3279 | 3655 |
| BMI_age9_log | - | - | - | - | - | - | - | - | 0.6993 | 0.3507 | 0.0462 | 3655 |
| Intercept variance | 14.6622 | 3.8291 | - | - | 16.1018 | 4.0127 | - | - | 13.3449 | 3.6531 | - | - |
| Age (slope) variance | 0.0725 | 0.2693 | - | - | 0.0791 | 0.2813 | - | - | 0.0709 | 0.2662 | - | - |
| Quadratic variance | 0.001 | 0.0318 | - | - | 0.0012 | 0.035 | - | - | 0.001 | 0.0318 | - | - |
| Intercept/age covariance | 0.7302 | 0.7081 | - | - | 0.804 | 0.7124 | - | - | 0.6687 | 0.6876 | - | - |
| Intercept/Quadratic covariance | -0.0984 | -0.809 | - | - | -0.1152 | -0.8207 | - | - | -0.0917 | -0.7887 | - | - |
| Age (slope)/Quadratic covariance | -0.0027 | -0.3102 | - | - | -0.0036 | -0.3697 | - | - | -0.0025 | -0.2904 | - | - |
| Residual variance | 13.4242 | 3.6639 | - | - | 13.1451 | 3.6256 | - | - | 13.271 | 3.6429 | - | - |
| Deviance | 145885.6449 | | | | 145834.2272 | | | | 130934.3575 | | | |
| AIC | 145929.6449 | | | | 145880.2272 | | | | 130984.3575 | | | |
| BIC | 146108.7028 | | | | 146067.424 | | | | 131185.2051 | | | |

Supplementary Table 15. Model estimates for sensitivity analysis in ALSPAC. Townsend quintiles used instead of maternal education.

| | Fully Adjusted | | | | Townsend Adjusted | | | |
|---|---|---|---|---|---|---|---|---|
| Parameter | Estimate | SE | p-value | N | Estimate | SE | p-value | N |
| Intercept | 3.1255 | 0.9109 | 0.0006 | 4264 | 3.5398 | 1.0159 | 0.0005 | 3572 |
| age | 0.2389 | 0.023 | <0.0001 | 4264 | 0.2337 | 0.0247 | <0.0001 | 3572 |
| age^2 (acceleration) | <0.0001 | 0.0049 | <0.0001 | 4264 | <0.0001 | 0.0052 | <0.0001 | 3572 |
| age^3 (cubic change) | <0.0001 | 0.0004 | 0.0001 | 4264 | <0.0001 | 0.0005 | 0.0008 | 3572 |
| age^4 (quartic change) | 0.0003 | <0.0001 | <0.0001 | 4264 | 0.0003 | <0.0001 | <0.0001 | 3572 |
| Sex1 | 0.762 | 0.0904 | <0.0001 | 4264 | 0.7325 | 0.1004 | <0.0001 | 3572 |
| Maternal.education.at.birth1 | <0.0001 | 0.0895 | 0.2825 | 4264 | - | - | - | - |
| BMI_age9_log | 0.8442 | 0.3204 | 0.0084 | 4264 | 0.6692 | 0.3575 | 0.0612 | 3572 |
| IL6_tertileMiddle | 0.125 | 0.185 | 0.4994 | 4264 | 0.0261 | 0.2107 | 0.9013 | 3572 |
| IL6_tertileTop | 0.5781 | 0.1874 | 0.002 | 4264 | 0.6222 | 0.2119 | 0.0033 | 3572 |
| age:IL6_tertileMiddle | <0.0001 | 0.0327 | 0.8672 | 4264 | <0.0001 | 0.0353 | 0.8139 | 3572 |
| age:IL6_tertileTop | <0.0001 | 0.0328 | 0.4683 | 4264 | <0.0001 | 0.0352 | 0.7956 | 3572 |
| age^2 (acceleration):IL6_tertileMiddle | <0.0001 | 0.0069 | 0.45 | 4264 | <0.0001 | 0.0074 | 0.2672 | 3572 |
| age^2 (acceleration):IL6_tertileTop | <0.0001 | 0.0069 | 0.4695 | 4264 | <0.0001 | 0.0074 | 0.5928 | 3572 |
| age^3 (cubic change):IL6_tertileMiddle | <0.0001 | 0.0006 | 0.9216 | 4264 | <0.0001 | 0.0007 | 0.5787 | 3572 |
| age^3 (cubic change):IL6_tertileTop | 0.0009 | 0.0006 | 0.1731 | 4264 | 0.0008 | 0.0007 | 0.2596 | 3572 |
| age^4 (quartic change):IL6_tertileMiddle | <0.0001 | <0.0001 | 0.9983 | 4264 | <0.0001 | <0.0001 | 0.5242 | 3572 |
| age^4 (quartic change):IL6_tertileTop | <0.0001 | <0.0001 | 0.5005 | 4264 | <0.0001 | <0.0001 | 0.5114 | 3572 |
| kqTownsendq52 | - | - | - | - | 0.1417 | 0.1459 | 0.3314 | 3572 |
| kqTownsendq53 | - | - | - | - | 0.0209 | 0.1401 | 0.8814 | 3572 |
| kqTownsendq54 | - | - | - | - | 0.1053 | 0.1374 | 0.4433 | 3572 |
| kqTownsendq55 | - | - | - | - | 0.3397 | 0.2037 | 0.0954 | 3572 |
| Intercept variance | 12.9107 | 3.5932 | - | - | 15.3406 | 3.9167 | - | - |
| Age (slope) variance | 0.0719 | 0.2682 | - | - | 0.0824 | 0.287 | - | - |
| Quadratic variance | 0.001 | 0.031 | - | - | 0.0012 | 0.0342 | - | - |
| Intercept/age covariance | 0.6619 | 0.6868 | - | - | 0.7975 | 0.7094 | - | - |
| Intercept/Quadratic covariance | -0.0854 | -0.7664 | - | - | -0.1084 | -0.8098 | - | - |
| Age (slope)/Quadratic covariance | -0.0023 | -0.2732 | - | - | -0.0037 | -0.3785 | - | - |

| | Fully Adjusted | | | | Townsend Adjusted | | | |
|---|---|---|---|---|---|---|---|---|
| Parameter | Estimate | SE | p-value | N | Estimate | SE | p-value | N |
| Residual variance | 13.2588 | 3.6413 | - | - | 12.9236 | 3.5949 | - | - |
| Deviance | 152823.3672 | | | | 131664.6607 | | | |
| AIC | 152873.3672 | | | | 131720.6607 | | | |
| BIC | 153078.0876 | | | | 131945.7851 | | | |

Supplementary Table 16. Model estimates from linear regression of number of SMFQ questionnaires participants completed on IL-6 tertile in ALSPAC. Total number of time points were 11.

| term | estimate | std.error | statistic | p.value |
|---|---|---|---|---|
| (Intercept) | 6.139 | 0.079 | 77.717 | p<0.0001 |
| IL6_tertileMiddle | -0.076 | 0.112 | -0.675 | 0.5 |
| IL6_tertileTop | 0.007 | 0.112 | 0.060 | 0.952 |

Supplementary Table 17. Estimated depression scores for each IL-6 tertile trajectory at ages 40, 50, 60, 70 and 80 years, in UK Biobank.

| IL-6 Tertile Group | age | estimate | 95% CI |
|---|---|---|---|
| PHQ-2 Score [IL-6 tertile = Bottom ] | Age 40 | 0.983 | 0.7 - 1.265 |
| PHQ-2 Score [IL-6 tertile = Middle ] | Age 40 | 1.112 | 0.827 - 1.397 |
| PHQ-2 Score [IL-6 tertile = Top ] | Age 40 | 1.170 | 0.882 - 1.458 |
| PHQ-2 Score [IL-6 tertile = Bottom ] | Age 50 | 0.921 | 0.651 - 1.19 |
| PHQ-2 Score [IL-6 tertile = Middle ] | Age 50 | 0.956 | 0.686 - 1.226 |
| PHQ-2 Score [IL-6 tertile = Top ] | Age 50 | 0.980 | 0.71 - 1.25 |
| PHQ-2 Score [IL-6 tertile = Bottom ] | Age 60 | 0.875 | 0.609 - 1.141 |
| PHQ-2 Score [IL-6 tertile = Middle ] | Age 60 | 0.863 | 0.597 - 1.129 |
| PHQ-2 Score [IL-6 tertile = Top ] | Age 60 | 0.874 | 0.609 - 1.14 |
| PHQ-2 Score [IL-6 tertile = Bottom ] | Age 70 | 0.846 | 0.579 - 1.112 |
| PHQ-2 Score [IL-6 tertile = Middle ] | Age 70 | 0.831 | 0.564 - 1.098 |
| PHQ-2 Score [IL-6 tertile = Top ] | Age 70 | 0.854 | 0.587 - 1.121 |
| PHQ-2 Score [IL-6 tertile = Bottom ] | Age 80 | 0.833 | 0.555 - 1.11 |
| PHQ-2 Score [IL-6 tertile = Middle ] | Age 80 | 0.862 | 0.585 - 1.139 |
| PHQ-2 Score [IL-6 tertile = Top ] | Age 80 | 0.919 | 0.641 - 1.197 |

Supplementary Table 18. Model estimates for main analysis in UK Biobank. IL-6 as a categorical variable (tertiles).

| | Unadjusted | | | | Sex Adjusted | | | | Fully Adjusted | | | |
|---|---|---|---|---|---|---|---|---|---|---|---|---|
| Parameter | Estimate | SE | p-value | N | Estimate | SE | p-value | N | Estimate | SE | p-value | N |
| Intercept | 0.6039 | 0.1323 | <0.0001 | 39613 | 0.5494 | 0.1323 | <0.0001 | 39613 | 0.7142 | 0.1318 | <0.0001 | 39285 |
| age | <0.0001 | 0.0007 | <0.0001 | 39613 | <0.0001 | 0.0007 | <0.0001 | 39613 | <0.0001 | 0.0007 | <0.0001 | 39285 |
| age^2 (acceleration) | <0.0001 | <0.0001 | 0.097 | 39613 | <0.0001 | <0.0001 | 0.0822 | 39613 | <0.0001 | <0.0001 | 0.096 | 39285 |
| Batch1 | 0.0243 | 0.1323 | 0.8542 | 39613 | 0.0263 | 0.1321 | 0.8423 | 39613 | <0.0001 | 0.1303 | 0.9233 | 39285 |
| Batch2 | 0.0131 | 0.1305 | 0.92 | 39613 | 0.0141 | 0.1303 | 0.914 | 39613 | <0.0001 | 0.1285 | 0.8529 | 39285 |
| Batch3 | 0.0419 | 0.1301 | 0.7474 | 39613 | 0.043 | 0.1299 | 0.7409 | 39613 | 0.0072 | 0.1281 | 0.9554 | 39285 |
| Batch4 | 0.0676 | 0.1303 | 0.6039 | 39613 | 0.0685 | 0.1301 | 0.5985 | 39613 | 0.0306 | 0.1283 | 0.8114 | 39285 |
| Batch5 | 0.0298 | 0.1303 | 0.8189 | 39613 | 0.0315 | 0.1301 | 0.8086 | 39613 | <0.0001 | 0.1283 | 0.9396 | 39285 |
| Batch6 | 0.0367 | 0.1303 | 0.7782 | 39613 | 0.0379 | 0.1301 | 0.7707 | 39613 | 0.0033 | 0.1283 | 0.9794 | 39285 |
| Batch7 | 0.0257 | 0.1305 | 0.8438 | 39613 | 0.0267 | 0.1303 | 0.8377 | 39613 | <0.0001 | 0.1285 | 0.9205 | 39285 |
| Assessment_centre_baseline Birmingham | <0.0001 | 0.0344 | 0.3281 | 39613 | <0.0001 | 0.0344 | 0.3899 | 39613 | 0.1492 | 0.0352 | <0.0001 | 39285 |
| Assessment_centre_baseline Bristol | <0.0001 | 0.0316 | <0.0001 | 39613 | <0.0001 | 0.0316 | <0.0001 | 39613 | 0.064 | 0.0332 | 0.0541 | 39285 |
| Assessment_centre_baseline Bury | <0.0001 | 0.033 | <0.0001 | 39613 | <0.0001 | 0.0329 | <0.0001 | 39613 | 0.0439 | 0.0343 | 0.201 | 39285 |
| Assessment_centre_baseline Cardiff | <0.0001 | 0.0361 | 0.0447 | 39613 | <0.0001 | 0.036 | 0.0531 | 39613 | 0.1604 | 0.0374 | <0.0001 | 39285 |
| Assessment_centre_baseline Croydon | <0.0001 | 0.0336 | 0.0762 | 39613 | <0.0001 | 0.0336 | 0.0771 | 39613 | 0.1229 | 0.0342 | 0.0003 | 39285 |
| Assessment_centre_baseline Edinburgh | <0.0001 | 0.0369 | <0.0001 | 39613 | <0.0001 | 0.0368 | <0.0001 | 39613 | 0.0332 | 0.038 | 0.3821 | 39285 |
| Assessment_centre_baseline Glasgow | <0.0001 | 0.0362 | 0.0031 | 39613 | <0.0001 | 0.0361 | 0.0036 | 39613 | 0.0652 | 0.0367 | 0.0758 | 39285 |
| Assessment_centre_baseline Hounslow | <0.0001 | 0.0332 | 0.0002 | 39613 | <0.0001 | 0.0331 | 0.0002 | 39613 | 0.0548 | 0.0337 | 0.1035 | 39285 |
| Assessment_centre_baseline Leeds | <0.0001 | 0.0307 | <0.0001 | 39613 | <0.0001 | 0.0307 | <0.0001 | 39613 | 0.0468 | 0.0322 | 0.1459 | 39285 |
| Assessment_centre_baseline Liverpool | <0.0001 | 0.0333 | 0.0002 | 39613 | <0.0001 | 0.0333 | 0.0003 | 39613 | 0.0892 | 0.0344 | 0.0095 | 39285 |

| | Unadjusted | | | | Sex Adjusted | | | | Fully Adjusted | | | |
|---|---|---|---|---|---|---|---|---|---|---|---|---|
| Parameter | Estimate | SE | p-value | N | Estimate | SE | p-value | N | Estimate | SE | p-value | N |
| Assessment_centre_baseline Manchester | <0.0001 | 0.0379 | 0.2771 | 39613 | <0.0001 | 0.0379 | 0.3268 | 39613 | 0.1497 | 0.0385 | 0.0001 | 39285 |
| Assessment_centre_baseline Middlesborough | <0.0001 | 0.0346 | <0.0001 | 39613 | <0.0001 | 0.0346 | <0.0001 | 39613 | 0.0843 | 0.0358 | 0.0185 | 39285 |
| Assessment_centre_baseline Newcastle | <0.0001 | 0.0323 | <0.0001 | 39613 | <0.0001 | 0.0322 | <0.0001 | 39613 | 0.0529 | 0.0333 | 0.1122 | 39285 |
| Assessment_centre_baseline Nottingham | <0.0001 | 0.0315 | <0.0001 | 39613 | <0.0001 | 0.0315 | <0.0001 | 39613 | 0.1048 | 0.033 | 0.0015 | 39285 |
| Assessment_centre_baseline Oxford | <0.0001 | 0.0395 | <0.0001 | 39613 | <0.0001 | 0.0395 | <0.0001 | 39613 | 0.0548 | 0.0405 | 0.1764 | 39285 |
| Assessment_centre_baseline Reading | <0.0001 | 0.0338 | <0.0001 | 39613 | <0.0001 | 0.0338 | <0.0001 | 39613 | 0.0665 | 0.0357 | 0.0625 | 39285 |
| Assessment_centre_baseline Sheffield | <0.0001 | 0.0327 | 0.0001 | 39613 | <0.0001 | 0.0326 | 0.0002 | 39613 | 0.1025 | 0.0339 | 0.0025 | 39285 |
| Assessment_centre_baseline Stoke | <0.0001 | 0.0363 | 0.0005 | 39613 | <0.0001 | 0.0363 | 0.0011 | 39613 | 0.1303 | 0.0377 | 0.0005 | 39285 |
| Assessment_centre_baseline Swansea | <0.0001 | 0.0933 | 0.7119 | 39613 | <0.0001 | 0.0932 | 0.7404 | 39613 | 0.1484 | 0.093 | 0.1105 | 39285 |
| Assessment_centre_baseline Wrexham | <0.0001 | 0.16 | 0.9902 | 39613 | <0.0001 | 0.1598 | 0.995 | 39613 | 0.2073 | 0.158 | 0.1896 | 39285 |
| IL6_tertileMiddle | 0.0263 | 0.0126 | 0.0362 | 39613 | 0.0277 | 0.0125 | 0.0273 | 39613 | <0.0001 | 0.0127 | 0.1631 | 39285 |
| IL6_tertileTop | 0.0969 | 0.0131 | <0.0001 | 39613 | 0.0995 | 0.013 | <0.0001 | 39613 | <0.0001 | 0.0138 | 0.7163 | 39285 |
| age:IL6_tertileMiddle | <0.0001 | 0.001 | 0.1028 | 39613 | <0.0001 | 0.001 | 0.1092 | 39613 | <0.0001 | 0.001 | 0.2842 | 39285 |
| age:IL6_tertileTop | <0.0001 | 0.0011 | 0.1408 | 39613 | <0.0001 | 0.0011 | 0.1699 | 39613 | <0.0001 | 0.0011 | 0.7042 | 39285 |
| age^2 (acceleration):IL6_tertileMiddle | 0.0002 | <0.0001 | 0.0007 | 39613 | 0.0002 | <0.0001 | 0.0007 | 39613 | 0.0002 | <0.0001 | 0.0015 | 39285 |
| age^2 (acceleration):IL6_tertileTop | 0.0003 | <0.0001 | <0.0001 | 39613 | 0.0003 | <0.0001 | <0.0001 | 39613 | 0.0003 | <0.0001 | <0.0001 | 39285 |
| SexFemale | - | - | - | - | 0.0907 | 0.0096 | <0.0001 | 39613 | 0.1218 | 0.0096 | <0.0001 | 39285 |
| smoking_statusNever | - | - | - | - | - | - | - | - | <0.0001 | 0.0169 | <0.0001 | 39285 |

| | Unadjusted | | | | Sex Adjusted | | | | Fully Adjusted | | | |
|---|---|---|---|---|---|---|---|---|---|---|---|---|
| Parameter | Estimate | SE | p-value | N | Estimate | SE | p-value | N | Estimate | SE | p-value | N |
| smoking_statusPrevious | - | - | - | - | - | - | - | - | <0.0001 | 0.0175 | <0.0001 | 39285 |
| Townsend | - | - | - | - | - | - | - | - | 0.1144 | 0.0054 | <0.0001 | 39285 |
| BMI | - | - | - | - | - | - | - | - | 0.072 | 0.0052 | <0.0001 | 39285 |
| Intercept variance | 0.5145 | 0.7173 | - | - | 0.5122 | 0.7157 | - | - | 0.4906 | 0.7004 | - | - |
| Age (slope) variance | 0.0011 | 0.0333 | - | - | 0.0011 | 0.0332 | - | - | 0.0011 | 0.0328 | - | - |
| Intercept/age covariance | -0.0025 | -0.1067 | - | - | -0.0026 | -0.1111 | - | - | -0.0021 | -0.0899 | - | - |
| Residual variance | 0.5893 | 0.7677 | - | - | 0.5895 | 0.7678 | - | - | 0.5885 | 0.7671 | - | - |
| Deviance | 283287.1176 | | | | 283197.3246 | | | | 280149.7478 | | | |
| AIC | 283367.1176 | | | | 283279.3246 | | | | 280239.7478 | | | |
| BIC | 283748.1402 | | | | 283669.8728 | | | | 280668.1232 | | | |

Supplementary Table 19. Model estimates for main analysis in UK Biobank split by sex. IL-6 as a categorical variable assigned to female and males separately.

| | Unadjusted | | | | Fully Adjusted | | | |
|---|---|---|---|---|---|---|---|---|
| Parameter | Estimate | SE | p-value | N | Estimate | SE | p-value | N |
| Intercept | 0.5794 | 0.0318 | <0.0001 | 39613 | 0.7051 | 0.0364 | <0.0001 | 39285 |
| age | <0.0001 | 0.0011 | <0.0001 | 39613 | <0.0001 | 0.0011 | <0.0001 | 39285 |
| age^2 (acceleration) | 0.0001 | <0.0001 | 0.0982 | 39613 | 0.0001 | <0.0001 | 0.0986 | 39285 |
| Batch | 0.0001 | 0.0028 | 0.9609 | 39613 | <0.0001 | 0.0028 | 0.9655 | 39285 |
| Assessment_centre_baseline Birmingham | <0.0001 | 0.0344 | 0.3988 | 39613 | 0.1492 | 0.0351 | <0.0001 | 39285 |
| Assessment_centre_baseline Bristol | <0.0001 | 0.0316 | <0.0001 | 39613 | 0.064 | 0.0332 | 0.0536 | 39285 |
| Assessment_centre_baseline Bury | <0.0001 | 0.0329 | <0.0001 | 39613 | 0.0431 | 0.0343 | 0.209 | 39285 |
| Assessment_centre_baseline Cardiff | <0.0001 | 0.0361 | 0.058 | 39613 | 0.1612 | 0.0374 | <0.0001 | 39285 |
| Assessment_centre_baseline Croydon | <0.0001 | 0.0336 | 0.0881 | 39613 | 0.1244 | 0.0342 | 0.0003 | 39285 |
| Assessment_centre_baseline Edinburgh | <0.0001 | 0.0369 | <0.0001 | 39613 | 0.0337 | 0.038 | 0.3744 | 39285 |
| Assessment_centre_baseline Glasgow | <0.0001 | 0.0361 | 0.0037 | 39613 | 0.0652 | 0.0367 | 0.0755 | 39285 |
| Assessment_centre_baseline Hounslow | <0.0001 | 0.0331 | 0.0003 | 39613 | 0.0556 | 0.0336 | 0.0984 | 39285 |
| Assessment_centre_baseline Leeds | <0.0001 | 0.0307 | <0.0001 | 39613 | 0.047 | 0.0322 | 0.144 | 39285 |
| Assessment_centre_baseline Liverpool | <0.0001 | 0.0333 | 0.0003 | 39613 | 0.0892 | 0.0343 | 0.0094 | 39285 |
| Assessment_centre_baseline Manchester | <0.0001 | 0.0379 | 0.3435 | 39613 | 0.1503 | 0.0385 | <0.0001 | 39285 |
| Assessment_centre_baseline Middlesborough | <0.0001 | 0.0346 | <0.0001 | 39613 | 0.0849 | 0.0357 | 0.0175 | 39285 |

| | Unadjusted | | | | Fully Adjusted | | | |
|---|---|---|---|---|---|---|---|---|
| Parameter | Estimate | SE | p-value | N | Estimate | SE | p-value | N |
| Assessment_centre_baseline Newcastle | <0.0001 | 0.0322 | <0.0001 | 39613 | 0.0525 | 0.0332 | 0.1142 | 39285 |
| Assessment_centre_baseline Nottingham | <0.0001 | 0.0315 | <0.0001 | 39613 | 0.1054 | 0.033 | 0.0014 | 39285 |
| Assessment_centre_baseline Oxford | <0.0001 | 0.0395 | <0.0001 | 39613 | 0.0558 | 0.0405 | 0.168 | 39285 |
| Assessment_centre_baseline Reading | <0.0001 | 0.0338 | <0.0001 | 39613 | 0.0663 | 0.0356 | 0.0628 | 39285 |
| Assessment_centre_baseline Sheffield | <0.0001 | 0.0327 | 0.0002 | 39613 | 0.1023 | 0.0338 | 0.0025 | 39285 |
| Assessment_centre_baseline Stoke | <0.0001 | 0.0363 | 0.0013 | 39613 | 0.1308 | 0.0376 | 0.0005 | 39285 |
| Assessment_centre_baseline Swansea | <0.0001 | 0.0932 | 0.7955 | 39613 | 0.1548 | 0.0928 | 0.0954 | 39285 |
| Assessment_centre_baseline Wrexham | <0.0001 | 0.1598 | 0.9571 | 39613 | 0.2002 | 0.1577 | 0.2043 | 39285 |
| IL6_sex_tertileMale_Middle | 0.0292 | 0.0189 | 0.1216 | 39613 | <0.0001 | 0.0188 | 0.5455 | 39285 |
| IL6_sex_tertileMale_Top | 0.1078 | 0.0194 | <0.0001 | 39613 | 0.0061 | 0.0197 | 0.7555 | 39285 |
| IL6_sex_tertileFemale_Bottom | 0.1026 | 0.0178 | <0.0001 | 39613 | 0.1377 | 0.0177 | <0.0001 | 39285 |
| IL6_sex_tertileFemale_Middle | 0.1299 | 0.018 | <0.0001 | 39613 | 0.1158 | 0.0178 | <0.0001 | 39285 |
| IL6_sex_tertileFemale_Top | 0.1968 | 0.0187 | <0.0001 | 39613 | 0.1249 | 0.0189 | <0.0001 | 39285 |
| age:IL6_sex_tertileMale_Middle | <0.0001 | 0.0015 | 0.1149 | 39613 | <0.0001 | 0.0015 | 0.165 | 39285 |
| age:IL6_sex_tertileMale_Top | <0.0001 | 0.0016 | 0.3424 | 39613 | <0.0001 | 0.0016 | 0.7049 | 39285 |
| age:IL6_sex_tertileFemale_Bottom | 0.0031 | 0.0015 | 0.0361 | 39613 | 0.0023 | 0.0015 | 0.1153 | 39285 |
| age:IL6_sex_tertileFemale_Middle | 0.0022 | 0.0015 | 0.1438 | 39613 | 0.0021 | 0.0015 | 0.1489 | 39285 |

| | Unadjusted | | | | Fully Adjusted | | | |
|---|---|---|---|---|---|---|---|---|
| Parameter | Estimate | SE | p-value | N | Estimate | SE | p-value | N |
| age:IL6_sex_tertileFemale_Top | 0.0016 | 0.0016 | 0.3016 | 39613 | 0.002 | 0.0016 | 0.1991 | 39285 |
| age^2 (acceleration):IL6_sex_tertile Male_Middle | 0.0002 | 0.0001 | 0.0221 | 39613 | 0.0002 | 0.0001 | 0.0541 | 39285 |
| age^2 (acceleration):IL6_sex_tertile Male_Top | 0.0004 | 0.0001 | 0.0026 | 39613 | 0.0004 | 0.0001 | 0.0017 | 39285 |
| age^2 (acceleration):IL6_sex_tertile Female_Bottom | <0.0001 | <0.0001 | 0.499 | 39613 | <0.0001 | <0.0001 | 0.4636 | 39285 |
| age^2 (acceleration):IL6_sex_tertile Female_Middle | 0.0002 | 0.0001 | 0.0761 | 39613 | 0.0002 | 0.0001 | 0.0799 | 39285 |
| age^2 (acceleration):IL6_sex_tertile Female_Top | 0.0003 | 0.0001 | 0.017 | 39613 | 0.0003 | 0.0001 | 0.02 | 39285 |
| smoking_statusNever | - | - | - | - | <0.0001 | 0.0169 | <0.0001 | 39285 |
| smoking_statusPrevious | - | - | - | - | <0.0001 | 0.0175 | <0.0001 | 39285 |
| Townsend | - | - | - | - | 0.1141 | 0.0054 | <0.0001 | 39285 |
| BMI | - | - | - | - | 0.0721 | 0.0052 | <0.0001 | 39285 |
| Intercept variance | 0.5128 | 0.7161 | - | - | 0.4881 | 0.6986 | - | - |
| Age (slope) variance | 0.0011 | 0.0332 | - | - | 0.0011 | 0.0327 | - | - |
| Intercept/age covariance | -0.0026 | -0.1114 | - | - | -0.0021 | -0.0913 | - | - |
| Residual variance | 0.5894 | 0.7677 | - | - | 0.5897 | 0.7679 | - | - |
| Deviance | 283186.0579 | | | | 280144.7292 | | | |
| AIC | 283272.0579 | | | | 280238.7292 | | | |
| BIC | 283681.6572 | | | | 280686.1435 | | | |

Supplementary Table 20. Estimated depression scores for each IL-6 tertile trajectory at ages 40, 50, 60, 70 and 80 years, in UK Biobank, split by sex.

| IL-6 Tertile Group | age | estimate | 95% CI |
|---|---|---|---|
| PHQ-2 Score [IL6_sex_tertile, level = Male_Bottom ] | Age 40 | 0.875 | 0.751 - 0.999 |
| PHQ-2 Score [IL6_sex_tertile, level = Male_Middle ] | Age 40 | 1.021 | 0.882 - 1.16 |
| PHQ-2 Score [IL6_sex_tertile, level = Male_Top ] | Age 40 | 1.090 | 0.93 - 1.25 |
| PHQ-2 Score [IL6_sex_tertile, level = Female_Bottom ] | Age 40 | 0.921 | 0.801 - 1.041 |
| PHQ-2 Score [IL6_sex_tertile, level = Female_Middle ] | Age 40 | 1.038 | 0.904 - 1.172 |
| PHQ-2 Score [IL6_sex_tertile, level = Female_Top ] | Age 40 | 1.092 | 0.943 - 1.24 |
| PHQ-2 Score [IL6_sex_tertile, level = Male_Bottom ] | Age 50 | 0.785 | 0.694 - 0.877 |
| PHQ-2 Score [IL6_sex_tertile, level = Male_Middle ] | Age 50 | 0.837 | 0.743 - 0.931 |
| PHQ-2 Score [IL6_sex_tertile, level = Male_Top ] | Age 50 | 0.862 | 0.764 - 0.961 |
| PHQ-2 Score [IL6_sex_tertile, level = Female_Bottom ] | Age 50 | 0.881 | 0.79 - 0.971 |
| PHQ-2 Score [IL6_sex_tertile, level = Female_Middle ] | Age 50 | 0.904 | 0.811 - 0.997 |
| PHQ-2 Score [IL6_sex_tertile, level = Female_Top ] | Age 50 | 0.928 | 0.832 - 1.025 |
| PHQ-2 Score [IL6_sex_tertile, level = Male_Bottom ] | Age 60 | 0.720 | 0.635 - 0.806 |
| PHQ-2 Score [IL6_sex_tertile, level = Male_Middle ] | Age 60 | 0.717 | 0.632 - 0.803 |
| PHQ-2 Score [IL6_sex_tertile, level = Male_Top ] | Age 60 | 0.732 | 0.646 - 0.817 |
| PHQ-2 Score [IL6_sex_tertile, level = Female_Bottom ] | Age 60 | 0.850 | 0.766 - 0.935 |
| PHQ-2 Score [IL6_sex_tertile, level = Female_Middle ] | Age 60 | 0.831 | 0.746 - 0.916 |
| PHQ-2 Score [IL6_sex_tertile, level = Female_Top ] | Age 60 | 0.842 | 0.756 - 0.927 |
| PHQ-2 Score [IL6_sex_tertile, level = Male_Bottom ] | Age 70 | 0.679 | 0.591 - 0.767 |
| PHQ-2 Score [IL6_sex_tertile, level = Male_Middle ] | Age 70 | 0.663 | 0.577 - 0.75 |
| PHQ-2 Score [IL6_sex_tertile, level = Male_Top ] | Age 70 | 0.699 | 0.612 - 0.786 |
| PHQ-2 Score [IL6_sex_tertile, level = Female_Bottom ] | Age 70 | 0.830 | 0.743 - 0.916 |
| PHQ-2 Score [IL6_sex_tertile, level = Female_Middle ] | Age 70 | 0.819 | 0.732 - 0.905 |
| PHQ-2 Score [IL6_sex_tertile, level = Female_Top ] | Age 70 | 0.831 | 0.744 - 0.918 |
| PHQ-2 Score [IL6_sex_tertile, level = Male_Bottom ] | Age 80 | 0.663 | 0.544 - 0.782 |
| PHQ-2 Score [IL6_sex_tertile, level = Male_Middle ] | Age 80 | 0.674 | 0.562 - 0.787 |
| PHQ-2 Score [IL6_sex_tertile, level = Male_Top ] | Age 80 | 0.763 | 0.646 - 0.881 |
| PHQ-2 Score [IL6_sex_tertile, level = Female_Bottom ] | Age 80 | 0.819 | 0.705 - 0.932 |
| PHQ-2 Score [IL6_sex_tertile, level = Female_Middle ] | Age 80 | 0.866 | 0.756 - 0.977 |
| PHQ-2 Score [IL6_sex_tertile, level = Female_Top ] | Age 80 | 0.896 | 0.78 - 1.012 |

Supplementary Table 21. Model estimates for sensitivity analysis in UK Biobank. IL-6 as a continuous variable.

| | Unadjusted | | | | Sex Adjusted | | | | Fully Adjusted | | | |
|---|---|---|---|---|---|---|---|---|---|---|---|---|
| Parameter | Estimate | SE | p-value | N | Estimate | SE | p-value | N | Estimate | SE | p-value | N |
| Intercept | 0.6483 | 0.1322 | <0.0001 | 39613 | 0.5947 | 0.1322 | <0.0001 | 39613 | 0.7076 | 0.1315 | <0.0001 | 39285 |
| age | <0.0001 | 0.0004 | <0.0001 | 39613 | <0.0001 | 0.0004 | <0.0001 | 39613 | <0.0001 | 0.0004 | <0.0001 | 39285 |
| age^2 (acceleration) | 0.0003 | <0.0001 | <0.0001 | 39613 | 0.0003 | <0.0001 | <0.0001 | 39613 | 0.0003 | <0.0001 | <0.0001 | 39285 |
| Batch1 | 0.0206 | 0.1323 | 0.8762 | 39613 | 0.0225 | 0.1322 | 0.8646 | 39613 | <0.0001 | 0.1303 | 0.917 | 39285 |
| Batch2 | 0.0105 | 0.1305 | 0.9358 | 39613 | 0.0115 | 0.1304 | 0.9299 | 39613 | <0.0001 | 0.1285 | 0.8473 | 39285 |
| Batch3 | 0.0387 | 0.1301 | 0.7662 | 39613 | 0.0397 | 0.13 | 0.7601 | 39613 | 0.0062 | 0.1281 | 0.9614 | 39285 |
| Batch4 | 0.0645 | 0.1303 | 0.6203 | 39613 | 0.0654 | 0.1302 | 0.6152 | 39613 | 0.0297 | 0.1283 | 0.8169 | 39285 |
| Batch5 | 0.0271 | 0.1303 | 0.8353 | 39613 | 0.0287 | 0.1302 | 0.8252 | 39613 | <0.0001 | 0.1283 | 0.9343 | 39285 |
| Batch6 | 0.0332 | 0.1303 | 0.7989 | 39613 | 0.0344 | 0.1302 | 0.7917 | 39613 | 0.0023 | 0.1283 | 0.9856 | 39285 |
| Batch7 | 0.0228 | 0.1305 | 0.8614 | 39613 | 0.0237 | 0.1304 | 0.8555 | 39613 | <0.0001 | 0.1285 | 0.9139 | 39285 |
| Assessment_centre_baseline Birmingham | <0.0001 | 0.0344 | 0.3223 | 39613 | <0.0001 | 0.0344 | 0.3839 | 39613 | 0.1493 | 0.0351 | <0.0001 | 39285 |
| Assessment_centre_baseline Bristol | <0.0001 | 0.0316 | <0.0001 | 39613 | <0.0001 | 0.0316 | <0.0001 | 39613 | 0.0638 | 0.0332 | 0.0547 | 39285 |
| Assessment_centre_baseline Bury | <0.0001 | 0.033 | <0.0001 | 39613 | <0.0001 | 0.0329 | <0.0001 | 39613 | 0.0439 | 0.0343 | 0.2013 | 39285 |
| Assessment_centre_baseline Cardiff | <0.0001 | 0.0361 | 0.0405 | 39613 | <0.0001 | 0.0361 | 0.0483 | 39613 | 0.1601 | 0.0374 | <0.0001 | 39285 |
| Assessment_centre_baseline Croydon | <0.0001 | 0.0336 | 0.0738 | 39613 | <0.0001 | 0.0336 | 0.075 | 39613 | 0.1228 | 0.0342 | 0.0003 | 39285 |
| Assessment_centre_baseline Edinburgh | <0.0001 | 0.0369 | <0.0001 | 39613 | <0.0001 | 0.0369 | <0.0001 | 39613 | 0.0334 | 0.038 | 0.3802 | 39285 |
| Assessment_centre_baseline Glasgow | <0.0001 | 0.0362 | 0.0029 | 39613 | <0.0001 | 0.0361 | 0.0034 | 39613 | 0.065 | 0.0367 | 0.0765 | 39285 |
| Assessment_centre_baseline Hounslow | <0.0001 | 0.0332 | 0.0003 | 39613 | <0.0001 | 0.0331 | 0.0003 | 39613 | 0.0552 | 0.0337 | 0.1015 | 39285 |
| Assessment_centre_baseline Leeds | <0.0001 | 0.0307 | <0.0001 | 39613 | <0.0001 | 0.0307 | <0.0001 | 39613 | 0.0469 | 0.0322 | 0.1451 | 39285 |
| Assessment_centre_baseline Liverpool | <0.0001 | 0.0333 | 0.0002 | 39613 | <0.0001 | 0.0333 | 0.0003 | 39613 | 0.0895 | 0.0344 | 0.0093 | 39285 |

| | Unadjusted | | | | Sex Adjusted | | | | Fully Adjusted | | | |
|---|---|---|---|---|---|---|---|---|---|---|---|---|
| Parameter | Estimate | SE | p-value | N | Estimate | SE | p-value | N | Estimate | SE | p-value | N |
| Assessment_centre_baseline Manchester | <0.0001 | 0.0379 | 0.283 | 39613 | <0.0001 | 0.0379 | 0.3342 | 39613 | 0.1502 | 0.0385 | <0.0001 | 39285 |
| Assessment_centre_baseline Middlesborough | <0.0001 | 0.0346 | <0.0001 | 39613 | <0.0001 | 0.0346 | <0.0001 | 39613 | 0.0842 | 0.0358 | 0.0187 | 39285 |
| Assessment_centre_baseline Newcastle | <0.0001 | 0.0323 | <0.0001 | 39613 | <0.0001 | 0.0323 | <0.0001 | 39613 | 0.0529 | 0.0333 | 0.1123 | 39285 |
| Assessment_centre_baseline Nottingham | <0.0001 | 0.0315 | <0.0001 | 39613 | <0.0001 | 0.0315 | <0.0001 | 39613 | 0.1049 | 0.033 | 0.0015 | 39285 |
| Assessment_centre_baseline Oxford | <0.0001 | 0.0395 | <0.0001 | 39613 | <0.0001 | 0.0395 | <0.0001 | 39613 | 0.0558 | 0.0405 | 0.1686 | 39285 |
| Assessment_centre_baseline Reading | <0.0001 | 0.0338 | <0.0001 | 39613 | <0.0001 | 0.0338 | <0.0001 | 39613 | 0.0669 | 0.0357 | 0.061 | 39285 |
| Assessment_centre_baseline Sheffield | <0.0001 | 0.0327 | 0.0001 | 39613 | <0.0001 | 0.0327 | 0.0002 | 39613 | 0.1025 | 0.0339 | 0.0025 | 39285 |
| Assessment_centre_baseline Stoke | <0.0001 | 0.0363 | 0.0005 | 39613 | <0.0001 | 0.0363 | 0.0012 | 39613 | 0.1305 | 0.0377 | 0.0005 | 39285 |
| Assessment_centre_baseline Swansea | <0.0001 | 0.0933 | 0.7241 | 39613 | <0.0001 | 0.0932 | 0.7531 | 39613 | 0.1489 | 0.093 | 0.1094 | 39285 |
| Assessment_centre_baseline Wrexham | <0.0001 | 0.16 | 0.9998 | 39613 | 0.0008 | 0.1599 | 0.9959 | 39613 | 0.2085 | 0.158 | 0.187 | 39285 |
| IL6_INT | 0.0422 | 0.0054 | <0.0001 | 39613 | 0.0436 | 0.0054 | <0.0001 | 39613 | <0.0001 | 0.0058 | 0.762 | 39285 |
| age:IL6_INT | <0.0001 | 0.0005 | 0.2828 | 39613 | <0.0001 | 0.0005 | 0.322 | 39613 | <0.0001 | 0.0005 | 0.8974 | 39285 |
| age^2 (acceleration):IL6_INT | 0.0001 | <0.0001 | <0.0001 | 39613 | 0.0001 | <0.0001 | <0.0001 | 39613 | 0.0001 | <0.0001 | <0.0001 | 39285 |
| SexFemale | - | - | - | - | 0.0916 | 0.0096 | <0.0001 | 39613 | 0.122 | 0.0096 | <0.0001 | 39285 |
| smoking_statusNever | - | - | - | - | - | - | - | - | <0.0001 | 0.0169 | <0.0001 | 39285 |
| smoking_statusPrevious | - | - | - | - | - | - | - | - | <0.0001 | 0.0175 | <0.0001 | 39285 |
| Townsend | - | - | - | - | - | - | - | - | 0.1146 | 0.0054 | <0.0001 | 39285 |
| BMI | - | - | - | - | - | - | - | - | 0.0722 | 0.0052 | <0.0001 | 39285 |
| Intercept variance | 0.5143 | 0.7171 | - | - | 0.5135 | 0.7166 | - | - | 0.4905 | 0.7003 | - | - |
| Age (slope) variance | 0.0011 | 0.0333 | - | - | 0.0011 | 0.0332 | - | - | 0.0011 | 0.0327 | - | - |
| Intercept/age covariance | -0.0026 | -0.1074 | - | - | -0.0027 | -0.1122 | - | - | -0.0021 | -0.0909 | - | - |

| | Unadjusted | | | | Sex Adjusted | | | | Fully Adjusted | | | |
|---|---|---|---|---|---|---|---|---|---|---|---|---|
| Parameter | Estimate | SE | p-value | N | Estimate | SE | p-value | N | Estimate | SE | p-value | N |
| Residual variance | 0.5894 | 0.7677 | - | - | 0.589 | 0.7675 | - | - | 0.5887 | 0.7673 | - | - |
| Deviance | 283295.7949 | | | | 283204.3155 | | | | 280158.84 | | | |
| AIC | 283369.7949 | | | | 283280.3155 | | | | 280242.84 | | | |
| BIC | 283722.2408 | | | | 283642.287 | | | | 280642.6571 | | | |

Supplementary Table 22. Model estimates for sensitivity analysis in UK Biobank. Individuals taking anti-inflammatory medication removed.

| | Unadjusted | | | | Sex Adjusted | | | | Fully Adjusted | | | |
|---|---|---|---|---|---|---|---|---|---|---|---|---|
| Parameter | Estimate | SE | p-value | N | Estimate | SE | p-value | N | Estimate | SE | p-value | N |
| Intercept | 0.5848 | 0.1348 | <0.0001 | 33271 | 0.5308 | 0.1347 | <0.0001 | 33271 | 0.699 | 0.1346 | <0.0001 | 33021 |
| age | <0.0001 | 0.0007 | <0.0001 | 33271 | <0.0001 | 0.0007 | <0.0001 | 33271 | <0.0001 | 0.0007 | <0.0001 | 33021 |
| age^2 (acceleration) | <0.0001 | <0.0001 | 0.054 | 33271 | 0.0001 | <0.0001 | 0.0436 | 33271 | <0.0001 | <0.0001 | 0.0583 | 33021 |
| Batch1 | 0.0201 | 0.1347 | 0.8813 | 33271 | 0.0198 | 0.1345 | 0.8828 | 33271 | <0.0001 | 0.1329 | 0.8754 | 33021 |
| Batch2 | 0.0006 | 0.1327 | 0.9964 | 33271 | <0.0001 | 0.1325 | 0.9921 | 33271 | <0.0001 | 0.1309 | 0.7555 | 33021 |
| Batch3 | 0.0324 | 0.1322 | 0.8067 | 33271 | 0.031 | 0.132 | 0.8146 | 33271 | <0.0001 | 0.1304 | 0.9618 | 33021 |
| Batch4 | 0.0631 | 0.1325 | 0.6339 | 33271 | 0.0618 | 0.1322 | 0.64 | 33271 | 0.0188 | 0.1307 | 0.8859 | 33021 |
| Batch5 | 0.0287 | 0.1324 | 0.8286 | 33271 | 0.0277 | 0.1322 | 0.8341 | 33271 | <0.0001 | 0.1307 | 0.8988 | 33021 |
| Batch6 | 0.0281 | 0.1325 | 0.8318 | 33271 | 0.0271 | 0.1322 | 0.8377 | 33271 | <0.0001 | 0.1307 | 0.935 | 33021 |
| Batch7 | 0.0149 | 0.1327 | 0.9104 | 33271 | 0.0136 | 0.1325 | 0.918 | 33271 | <0.0001 | 0.1309 | 0.8315 | 33021 |
| Assessment_centre_baseline Birmingham | <0.0001 | 0.036 | 0.2788 | 33271 | <0.0001 | 0.036 | 0.3451 | 33271 | 0.142 | 0.0369 | 0.0001 | 33021 |
| Assessment_centre_baseline Bristol | <0.0001 | 0.0329 | <0.0001 | 33271 | <0.0001 | 0.0329 | <0.0001 | 33271 | 0.0684 | 0.0348 | 0.0492 | 33021 |
| Assessment_centre_baseline Bury | <0.0001 | 0.0346 | <0.0001 | 33271 | <0.0001 | 0.0345 | <0.0001 | 33271 | 0.035 | 0.0361 | 0.3326 | 33021 |
| Assessment_centre_baseline Cardiff | <0.0001 | 0.0376 | 0.0609 | 33271 | <0.0001 | 0.0376 | 0.0729 | 33271 | 0.1532 | 0.0391 | <0.0001 | 33021 |

| | Unadjusted | | | | Sex Adjusted | | | | Fully Adjusted | | | |
|---|---|---|---|---|---|---|---|---|---|---|---|---|
| Parameter | Estimate | SE | p-value | N | Estimate | SE | p-value | N | Estimate | SE | p-value | N |
| Assessment_centre_baseline Croydon | <0.0001 | 0.0353 | 0.0679 | 33271 | <0.0001 | 0.0353 | 0.0701 | 33271 | 0.1116 | 0.036 | 0.0019 | 33021 |
| Assessment_centre_baseline Edinburgh | <0.0001 | 0.0382 | <0.0001 | 33271 | <0.0001 | 0.0381 | <0.0001 | 33271 | 0.0458 | 0.0395 | 0.2458 | 33021 |
| Assessment_centre_baseline Glasgow | <0.0001 | 0.0377 | 0.0047 | 33271 | <0.0001 | 0.0377 | 0.0057 | 33271 | 0.0642 | 0.0385 | 0.0952 | 33021 |
| Assessment_centre_baseline Hounslow | <0.0001 | 0.0346 | <0.0001 | 33271 | <0.0001 | 0.0346 | <0.0001 | 33271 | 0.0317 | 0.0353 | 0.3682 | 33021 |
| Assessment_centre_baseline Leeds | <0.0001 | 0.0321 | <0.0001 | 33271 | <0.0001 | 0.032 | <0.0001 | 33271 | 0.0597 | 0.0337 | 0.077 | 33021 |
| Assessment_centre_baseline Liverpool | <0.0001 | 0.035 | 0.0001 | 33271 | <0.0001 | 0.0349 | 0.0002 | 33271 | 0.0678 | 0.0362 | 0.0608 | 33021 |
| Assessment_centre_baseline Manchester | <0.0001 | 0.0398 | 0.0781 | 33271 | <0.0001 | 0.0397 | 0.1082 | 33271 | 0.1137 | 0.0405 | 0.005 | 33021 |
| Assessment_centre_baseline Middlesborough | <0.0001 | 0.0361 | <0.0001 | 33271 | <0.0001 | 0.0361 | 0.0001 | 33271 | 0.0787 | 0.0375 | 0.036 | 33021 |
| Assessment_centre_baseline Newcastle | <0.0001 | 0.0339 | <0.0001 | 33271 | <0.0001 | 0.0338 | <0.0001 | 33271 | 0.0592 | 0.0351 | 0.0914 | 33021 |
| Assessment_centre_baseline Nottingham | <0.0001 | 0.0329 | <0.0001 | 33271 | <0.0001 | 0.0329 | 0.0001 | 33271 | 0.1049 | 0.0346 | 0.0025 | 33021 |
| Assessment_centre_baseline Oxford | <0.0001 | 0.0413 | 0.0001 | 33271 | <0.0001 | 0.0412 | 0.0001 | 33271 | 0.0777 | 0.0425 | 0.0674 | 33021 |
| Assessment_centre_baseline Reading | <0.0001 | 0.0351 | <0.0001 | 33271 | <0.0001 | 0.0351 | <0.0001 | 33271 | 0.0683 | 0.0372 | 0.0668 | 33021 |
| Assessment_centre_baseline Sheffield | <0.0001 | 0.0344 | 0.0009 | 33271 | <0.0001 | 0.0343 | 0.0011 | 33271 | 0.1061 | 0.0357 | 0.003 | 33021 |
| Assessment_centre_baseline Stoke | <0.0001 | 0.0381 | 0.0011 | 33271 | <0.0001 | 0.0381 | 0.0024 | 33271 | 0.1215 | 0.0397 | 0.0022 | 33021 |
| Assessment_centre_baseline Swansea | 0.0098 | 0.0986 | 0.9212 | 33271 | 0.012 | 0.0984 | 0.9026 | 33271 | 0.1711 | 0.0978 | 0.0801 | 33021 |
| Assessment_centre_baseline Wrexham | 0.1149 | 0.1708 | 0.5012 | 33271 | 0.1167 | 0.1705 | 0.494 | 33271 | 0.3237 | 0.1689 | 0.0554 | 33021 |

| | Unadjusted | | | | Sex Adjusted | | | | Fully Adjusted | | | |
|---|---|---|---|---|---|---|---|---|---|---|---|---|
| Parameter | Estimate | SE | p-value | N | Estimate | SE | p-value | N | Estimate | SE | p-value | N |
| IL6_tertileMiddle | 0.0225 | 0.013 | 0.084 | 33271 | 0.0233 | 0.013 | 0.0729 | 33271 | <0.0001 | 0.0131 | 0.2293 | 33021 |
| IL6_tertileTop | 0.0761 | 0.0138 | <0.0001 | 33271 | 0.0767 | 0.0138 | <0.0001 | 33271 | <0.0001 | 0.0146 | 0.4511 | 33021 |
| age:IL6_tertileMiddle | <0.0001 | 0.0011 | 0.1268 | 33271 | <0.0001 | 0.0011 | 0.1336 | 33271 | <0.0001 | 0.0011 | 0.2885 | 33021 |
| age:IL6_tertileTop | <0.0001 | 0.0012 | 0.0407 | 33271 | <0.0001 | 0.0012 | 0.0534 | 33271 | <0.0001 | 0.0012 | 0.2987 | 33021 |
| age^2 (acceleration):IL6_tertileMiddle | 0.0002 | <0.0001 | 0.0035 | 33271 | 0.0002 | <0.0001 | 0.0034 | 33271 | 0.0002 | <0.0001 | 0.0048 | 33021 |
| age^2 (acceleration):IL6_tertileTop | 0.0003 | <0.0001 | 0.0003 | 33271 | 0.0003 | <0.0001 | 0.0003 | 33271 | 0.0003 | <0.0001 | 0.0002 | 33021 |
| SexFemale | - | - | - | - | 0.0927 | 0.01 | <0.0001 | 33271 | 0.1192 | 0.0101 | <0.0001 | 33021 |
| smoking_statusNever | - | - | - | - | - | - | - | - | <0.0001 | 0.0179 | <0.0001 | 33021 |
| smoking_statusPrevious | - | - | - | - | - | - | - | - | <0.0001 | 0.0187 | <0.0001 | 33021 |
| Townsend | - | - | - | - | - | - | - | - | 0.107 | 0.0058 | <0.0001 | 33021 |
| BMI | - | - | - | - | - | - | - | - | 0.0602 | 0.0056 | <0.0001 | 33021 |
| Intercept variance | 0.4667 | 0.6832 | - | - | 0.4644 | 0.6814 | - | - | 0.4471 | 0.6686 | - | - |
| Age (slope) variance | 0.001 | 0.0322 | - | - | 0.001 | 0.0321 | - | - | 0.001 | 0.0318 | - | - |
| Intercept/age covariance | -0.003 | -0.135 | - | - | -0.003 | -0.139 | - | - | -0.0025 | -0.1172 | - | - |
| Residual variance | 0.5668 | 0.7529 | - | - | 0.567 | 0.753 | - | - | 0.5656 | 0.7521 | - | - |
| Deviance | 238382.6179 | | | | 238297.5013 | | | | 235956.7178 | | | |
| AIC | 238462.6179 | | | | 238379.5013 | | | | 236046.7178 | | | |
| BIC | 238837.5373 | | | | 238763.7937 | | | | 236468.253 | | | |

Supplementary Table 23. Model estimates for sensitivity analysis in UK Biobank. Individuals with inflammatory conditions removed.

| | Unadjusted | | | | Sex Adjusted | | | | Fully Adjusted | | | |
|---|---|---|---|---|---|---|---|---|---|---|---|---|
| Parameter | Estimate | SE | p-value | N | Estimate | SE | p-value | N | Estimate | SE | p-value | N |
| Intercept | 0.5848 | 0.1348 | <0.0001 | 33271 | 0.5308 | 0.1347 | <0.0001 | 33271 | 0.699 | 0.1346 | <0.0001 | 33021 |

| | Unadjusted | | | | Sex Adjusted | | | | Fully Adjusted | | | |
|---|---|---|---|---|---|---|---|---|---|---|---|---|
| Parameter | Estimate | SE | p-value | N | Estimate | SE | p-value | N | Estimate | SE | p-value | N |
| age | <0.0001 | 0.0007 | <0.0001 | 33271 | <0.0001 | 0.0007 | <0.0001 | 33271 | <0.0001 | 0.0007 | <0.0001 | 33021 |
| age^2 (acceleration) | <0.0001 | <0.0001 | 0.054 | 33271 | 0.0001 | <0.0001 | 0.0436 | 33271 | <0.0001 | <0.0001 | 0.0583 | 33021 |
| Batch1 | 0.0201 | 0.1347 | 0.8813 | 33271 | 0.0198 | 0.1345 | 0.8828 | 33271 | <0.0001 | 0.1329 | 0.8754 | 33021 |
| Batch2 | 0.0006 | 0.1327 | 0.9964 | 33271 | <0.0001 | 0.1325 | 0.9921 | 33271 | <0.0001 | 0.1309 | 0.7555 | 33021 |
| Batch3 | 0.0324 | 0.1322 | 0.8067 | 33271 | 0.031 | 0.132 | 0.8146 | 33271 | <0.0001 | 0.1304 | 0.9618 | 33021 |
| Batch4 | 0.0631 | 0.1325 | 0.6339 | 33271 | 0.0618 | 0.1322 | 0.64 | 33271 | 0.0188 | 0.1307 | 0.8859 | 33021 |
| Batch5 | 0.0287 | 0.1324 | 0.8286 | 33271 | 0.0277 | 0.1322 | 0.8341 | 33271 | <0.0001 | 0.1307 | 0.8988 | 33021 |
| Batch6 | 0.0281 | 0.1325 | 0.8318 | 33271 | 0.0271 | 0.1322 | 0.8377 | 33271 | <0.0001 | 0.1307 | 0.935 | 33021 |
| Batch7 | 0.0149 | 0.1327 | 0.9104 | 33271 | 0.0136 | 0.1325 | 0.918 | 33271 | <0.0001 | 0.1309 | 0.8315 | 33021 |
| Assessment_centre_baseline Birmingham | <0.0001 | 0.036 | 0.2788 | 33271 | <0.0001 | 0.036 | 0.3451 | 33271 | 0.142 | 0.0369 | 0.0001 | 33021 |
| Assessment_centre_baseline Bristol | <0.0001 | 0.0329 | <0.0001 | 33271 | <0.0001 | 0.0329 | <0.0001 | 33271 | 0.0684 | 0.0348 | 0.0492 | 33021 |
| Assessment_centre_baseline Bury | <0.0001 | 0.0346 | <0.0001 | 33271 | <0.0001 | 0.0345 | <0.0001 | 33271 | 0.035 | 0.0361 | 0.3326 | 33021 |
| Assessment_centre_baseline Cardiff | <0.0001 | 0.0376 | 0.0609 | 33271 | <0.0001 | 0.0376 | 0.0729 | 33271 | 0.1532 | 0.0391 | <0.0001 | 33021 |
| Assessment_centre_baseline Croydon | <0.0001 | 0.0353 | 0.0679 | 33271 | <0.0001 | 0.0353 | 0.0701 | 33271 | 0.1116 | 0.036 | 0.0019 | 33021 |
| Assessment_centre_baseline Edinburgh | <0.0001 | 0.0382 | <0.0001 | 33271 | <0.0001 | 0.0381 | <0.0001 | 33271 | 0.0458 | 0.0395 | 0.2458 | 33021 |
| Assessment_centre_baseline Glasgow | <0.0001 | 0.0377 | 0.0047 | 33271 | <0.0001 | 0.0377 | 0.0057 | 33271 | 0.0642 | 0.0385 | 0.0952 | 33021 |
| Assessment_centre_baseline Hounslow | <0.0001 | 0.0346 | <0.0001 | 33271 | <0.0001 | 0.0346 | <0.0001 | 33271 | 0.0317 | 0.0353 | 0.3682 | 33021 |
| Assessment_centre_baseline Leeds | <0.0001 | 0.0321 | <0.0001 | 33271 | <0.0001 | 0.032 | <0.0001 | 33271 | 0.0597 | 0.0337 | 0.077 | 33021 |
| Assessment_centre_baseline Liverpool | <0.0001 | 0.035 | 0.0001 | 33271 | <0.0001 | 0.0349 | 0.0002 | 33271 | 0.0678 | 0.0362 | 0.0608 | 33021 |
| Assessment_centre_baseline Manchester | <0.0001 | 0.0398 | 0.0781 | 33271 | <0.0001 | 0.0397 | 0.1082 | 33271 | 0.1137 | 0.0405 | 0.005 | 33021 |

| | Unadjusted | | | | Sex Adjusted | | | | Fully Adjusted | | | |
|---|---|---|---|---|---|---|---|---|---|---|---|---|
| Parameter | Estimate | SE | p-value | N | Estimate | SE | p-value | N | Estimate | SE | p-value | N |
| Assessment_centre_baseline Middlesborough | <0.0001 | 0.0361 | <0.0001 | 33271 | <0.0001 | 0.0361 | 0.0001 | 33271 | 0.0787 | 0.0375 | 0.036 | 33021 |
| Assessment_centre_baseline Newcastle | <0.0001 | 0.0339 | <0.0001 | 33271 | <0.0001 | 0.0338 | <0.0001 | 33271 | 0.0592 | 0.0351 | 0.0914 | 33021 |
| Assessment_centre_baseline Nottingham | <0.0001 | 0.0329 | <0.0001 | 33271 | <0.0001 | 0.0329 | 0.0001 | 33271 | 0.1049 | 0.0346 | 0.0025 | 33021 |
| Assessment_centre_baseline Oxford | <0.0001 | 0.0413 | 0.0001 | 33271 | <0.0001 | 0.0412 | 0.0001 | 33271 | 0.0777 | 0.0425 | 0.0674 | 33021 |
| Assessment_centre_baseline Reading | <0.0001 | 0.0351 | <0.0001 | 33271 | <0.0001 | 0.0351 | <0.0001 | 33271 | 0.0683 | 0.0372 | 0.0668 | 33021 |
| Assessment_centre_baseline Sheffield | <0.0001 | 0.0344 | 0.0009 | 33271 | <0.0001 | 0.0343 | 0.0011 | 33271 | 0.1061 | 0.0357 | 0.003 | 33021 |
| Assessment_centre_baseline Stoke | <0.0001 | 0.0381 | 0.0011 | 33271 | <0.0001 | 0.0381 | 0.0024 | 33271 | 0.1215 | 0.0397 | 0.0022 | 33021 |
| Assessment_centre_baseline Swansea | 0.0098 | 0.0986 | 0.9212 | 33271 | 0.012 | 0.0984 | 0.9026 | 33271 | 0.1711 | 0.0978 | 0.0801 | 33021 |
| Assessment_centre_baseline Wrexham | 0.1149 | 0.1708 | 0.5012 | 33271 | 0.1167 | 0.1705 | 0.494 | 33271 | 0.3237 | 0.1689 | 0.0554 | 33021 |
| IL6_tertileMiddle | 0.0225 | 0.013 | 0.084 | 33271 | 0.0233 | 0.013 | 0.0729 | 33271 | <0.0001 | 0.0131 | 0.2293 | 33021 |
| IL6_tertileTop | 0.0761 | 0.0138 | <0.0001 | 33271 | 0.0767 | 0.0138 | <0.0001 | 33271 | <0.0001 | 0.0146 | 0.4511 | 33021 |
| age:IL6_tertileMiddle | <0.0001 | 0.0011 | 0.1268 | 33271 | <0.0001 | 0.0011 | 0.1336 | 33271 | <0.0001 | 0.0011 | 0.2885 | 33021 |
| age:IL6_tertileTop | <0.0001 | 0.0012 | 0.0407 | 33271 | <0.0001 | 0.0012 | 0.0534 | 33271 | <0.0001 | 0.0012 | 0.2987 | 33021 |
| age^2 (acceleration):IL6_tertileMiddl e | 0.0002 | <0.0001 | 0.0035 | 33271 | 0.0002 | <0.0001 | 0.0034 | 33271 | 0.0002 | <0.0001 | 0.0048 | 33021 |
| age^2 (acceleration):IL6_tertileTop | 0.0003 | <0.0001 | 0.0003 | 33271 | 0.0003 | <0.0001 | 0.0003 | 33271 | 0.0003 | <0.0001 | 0.0002 | 33021 |
| SexFemale | - | - | - | - | 0.0927 | 0.01 | <0.0001 | 33271 | 0.1192 | 0.0101 | <0.0001 | 33021 |
| smoking_statusNever | - | - | - | - | - | - | - | - | <0.0001 | 0.0179 | <0.0001 | 33021 |
| smoking_statusPrevious | - | - | - | - | - | - | - | - | <0.0001 | 0.0187 | <0.0001 | 33021 |
| Townsend | - | - | - | - | - | - | - | - | 0.107 | 0.0058 | <0.0001 | 33021 |

| | Unadjusted | | | | Sex Adjusted | | | | Fully Adjusted | | | |
|---|---|---|---|---|---|---|---|---|---|---|---|---|
| Parameter | Estimate | SE | p-value | N | Estimate | SE | p-value | N | Estimate | SE | p-value | N |
| BMI | - | - | - | - | - | - | - | - | 0.0602 | 0.0056 | <0.0001 | 33021 |
| Intercept variance | 0.4667 | 0.6832 | - | - | 0.4644 | 0.6814 | - | - | 0.4471 | 0.6686 | - | - |
| Age (slope) variance | 0.001 | 0.0322 | - | - | 0.001 | 0.0321 | - | - | 0.001 | 0.0318 | - | - |
| Intercept/age covariance | -0.003 | -0.135 | - | - | -0.003 | -0.139 | - | - | -0.0025 | -0.1172 | - | - |
| Residual variance | 0.5668 | 0.7529 | - | - | 0.567 | 0.753 | - | - | 0.5656 | 0.7521 | - | - |
| Deviance | 238382.<br>6179 | | | | 238297.<br>5013 | | | | 235956.<br>7178 | | | |
| AIC | 238462.<br>6179 | | | | 238379.<br>5013 | | | | 236046.<br>7178 | | | |
| BIC | 238837.<br>5373 | | | | 238763.<br>7937 | | | | 236468.<br>253 | | | |

Supplementary Table 24. Model estimates for sensitivity analysis in UK Biobank. Individuals with BMI >= 40 removed.

| | Unadjusted | | | | Sex Adjusted | | | | Fully Adjusted | | | |
|---|---|---|---|---|---|---|---|---|---|---|---|---|
| Parameter | Estimate | SE | p-value | N | Estimate | SE | p-value | N | Estimate | SE | p-value | N |
| Intercept | 0.6176 | 0.1309 | <0.0001 | 38862 | 0.5634 | 0.1308 | <0.0001 | 38862 | 0.7107 | 0.1306 | <0.0001 | 38691 |
| age | <0.0001 | 0.0007 | <0.0001 | 38862 | <0.0001 | 0.0007 | <0.0001 | 38862 | <0.0001 | 0.0007 | <0.0001 | 38691 |
| age^2 (acceleration) | <0.0001 | <0.0001 | 0.0772 | 38862 | <0.0001 | <0.0001 | 0.0647 | 38862 | <0.0001 | <0.0001 | 0.0903 | 38691 |
| Batch1 | 0.0215 | 0.1309 | 0.8692 | 38862 | 0.0235 | 0.1306 | 0.857 | 38862 | <0.0001 | 0.1292 | 0.9283 | 38691 |
| Batch2 | 0.0051 | 0.1291 | 0.9683 | 38862 | 0.0063 | 0.1288 | 0.9613 | 38862 | <0.0001 | 0.1274 | 0.8251 | 38691 |
| Batch3 | 0.033 | 0.1286 | 0.7976 | 38862 | 0.0343 | 0.1284 | 0.7895 | 38862 | 0.0021 | 0.1269 | 0.9866 | 38691 |
| Batch4 | 0.0592 | 0.1288 | 0.6459 | 38862 | 0.0602 | 0.1286 | 0.6395 | 38862 | 0.0262 | 0.1271 | 0.8366 | 38691 |
| Batch5 | 0.0176 | 0.1288 | 0.8911 | 38862 | 0.0195 | 0.1286 | 0.8793 | 38862 | <0.0001 | 0.1271 | 0.9004 | 38691 |
| Batch6 | 0.0301 | 0.1288 | 0.8155 | 38862 | 0.0314 | 0.1286 | 0.8069 | 38862 | <0.0001 | 0.1271 | 0.9967 | 38691 |
| Batch7 | 0.016 | 0.129 | 0.9014 | 38862 | 0.0171 | 0.1288 | 0.8944 | 38862 | <0.0001 | 0.1274 | 0.8868 | 38691 |
| Assessment_centre_baseline Birmingham | <0.0001 | 0.0343 | 0.1164 | 38862 | <0.0001 | 0.0343 | 0.1428 | 38862 | 0.1354 | 0.035 | 0.0001 | 38691 |
| Assessment_centre_baseline Bristol | <0.0001 | 0.0315 | <0.0001 | 38862 | <0.0001 | 0.0314 | <0.0001 | 38862 | 0.0618 | 0.0331 | 0.0621 | 38691 |

| | Unadjusted | | | | Sex Adjusted | | | | Fully Adjusted | | | |
|---|---|---|---|---|---|---|---|---|---|---|---|---|
| Parameter | Estimate | SE | p-value | N | Estimate | SE | p-value | N | Estimate | SE | p-value | N |
| Assessment_centre_baseline Bury | <0.0001 | 0.0329 | <0.0001 | 38862 | <0.0001 | 0.0328 | <0.0001 | 38862 | 0.0394 | 0.0343 | 0.2501 | 38691 |
| Assessment_centre_baseline Cardiff | <0.0001 | 0.036 | 0.0227 | 38862 | <0.0001 | 0.0359 | 0.027 | 38862 | 0.1535 | 0.0373 | <0.0001 | 38691 |
| Assessment_centre_baseline Croydon | <0.0001 | 0.0335 | 0.0378 | 38862 | <0.0001 | 0.0335 | 0.0379 | 38862 | 0.1123 | 0.0341 | 0.001 | 38691 |
| Assessment_centre_baseline Edinburgh | <0.0001 | 0.0368 | <0.0001 | 38862 | <0.0001 | 0.0367 | <0.0001 | 38862 | 0.0334 | 0.0379 | 0.3791 | 38691 |
| Assessment_centre_baseline Glasgow | <0.0001 | 0.0361 | 0.0003 | 38862 | <0.0001 | 0.036 | 0.0004 | 38862 | 0.05 | 0.0367 | 0.1732 | 38691 |
| Assessment_centre_baseline Hounslow | <0.0001 | 0.033 | <0.0001 | 38862 | <0.0001 | 0.033 | <0.0001 | 38862 | 0.0495 | 0.0336 | 0.1407 | 38691 |
| Assessment_centre_baseline Leeds | <0.0001 | 0.0306 | <0.0001 | 38862 | <0.0001 | 0.0306 | <0.0001 | 38862 | 0.0412 | 0.0321 | 0.1995 | 38691 |
| Assessment_centre_baseline Liverpool | <0.0001 | 0.0332 | 0.0002 | 38862 | <0.0001 | 0.0332 | 0.0002 | 38862 | 0.0857 | 0.0343 | 0.0126 | 38691 |
| Assessment_centre_baseline Manchester | <0.0001 | 0.0378 | 0.1427 | 38862 | <0.0001 | 0.0378 | 0.1739 | 38862 | 0.137 | 0.0385 | 0.0004 | 38691 |
| Assessment_centre_baseline Middlesborough | <0.0001 | 0.0345 | <0.0001 | 38862 | <0.0001 | 0.0345 | <0.0001 | 38862 | 0.0824 | 0.0357 | 0.021 | 38691 |
| Assessment_centre_baseline Newcastle | <0.0001 | 0.0322 | <0.0001 | 38862 | <0.0001 | 0.0321 | <0.0001 | 38862 | 0.052 | 0.0332 | 0.1173 | 38691 |
| Assessment_centre_baseline Nottingham | <0.0001 | 0.0314 | <0.0001 | 38862 | <0.0001 | 0.0314 | <0.0001 | 38862 | 0.0995 | 0.0329 | 0.0025 | 38691 |
| Assessment_centre_baseline Oxford | <0.0001 | 0.0393 | <0.0001 | 38862 | <0.0001 | 0.0393 | <0.0001 | 38862 | 0.0456 | 0.0404 | 0.2587 | 38691 |
| Assessment_centre_baseline Reading | <0.0001 | 0.0337 | <0.0001 | 38862 | <0.0001 | 0.0336 | <0.0001 | 38862 | 0.0599 | 0.0356 | 0.092 | 38691 |
| Assessment_centre_baseline Sheffield | <0.0001 | 0.0326 | <0.0001 | 38862 | <0.0001 | 0.0325 | <0.0001 | 38862 | 0.0906 | 0.0338 | 0.0073 | 38691 |
| Assessment_centre_baseline Stoke | <0.0001 | 0.0362 | 0.0004 | 38862 | <0.0001 | 0.0362 | 0.0009 | 38862 | 0.1296 | 0.0376 | 0.0006 | 38691 |

| | Unadjusted | | | | Sex Adjusted | | | | Fully Adjusted | | | |
|---|---|---|---|---|---|---|---|---|---|---|---|---|
| Parameter | Estimate | SE | p-value | N | Estimate | SE | p-value | N | Estimate | SE | p-value | N |
| Assessment_centre_baseline Swansea | <0.0001 | 0.094 | 0.3608 | 38862 | <0.0001 | 0.0938 | 0.3807 | 38862 | 0.096 | 0.0936 | 0.305 | 38691 |
| Assessment_centre_baseline Wrexham | <0.0001 | 0.1606 | 0.9942 | 38862 | <0.0001 | 0.1603 | 0.9897 | 38862 | 0.2125 | 0.1589 | 0.1811 | 38691 |
| IL6_tertileMiddle | 0.0245 | 0.0125 | 0.0492 | 38862 | 0.026 | 0.0124 | 0.0371 | 38862 | <0.0001 | 0.0126 | 0.32 | 38691 |
| IL6_tertileTop | 0.0797 | 0.0131 | <0.0001 | 38862 | 0.0825 | 0.0131 | <0.0001 | 38862 | <0.0001 | 0.0137 | 0.8479 | 38691 |
| age:IL6_tertileMiddle | <0.0001 | 0.001 | 0.1585 | 38862 | <0.0001 | 0.001 | 0.1667 | 38862 | <0.0001 | 0.001 | 0.3434 | 38691 |
| age:IL6_tertileTop | <0.0001 | 0.0011 | 0.2295 | 38862 | <0.0001 | 0.0011 | 0.2639 | 38862 | <0.0001 | 0.0011 | 0.788 | 38691 |
| age^2 (acceleration):IL6_tertileMiddle | 0.0002 | <0.0001 | 0.0012 | 38862 | 0.0002 | <0.0001 | 0.0012 | 38862 | 0.0002 | <0.0001 | 0.0016 | 38691 |
| age^2 (acceleration):IL6_tertileTop | 0.0003 | <0.0001 | <0.0001 | 38862 | 0.0003 | <0.0001 | <0.0001 | 38862 | 0.0003 | <0.0001 | <0.0001 | 38691 |
| SexFemale | - | - | - | - | 0.0903 | 0.0095 | <0.0001 | 38862 | 0.1191 | 0.0096 | <0.0001 | 38691 |
| smoking_statusNever | - | - | - | - | - | - | - | - | <0.0001 | 0.0168 | <0.0001 | 38691 |
| smoking_statusPrevious | - | - | - | - | - | - | - | - | <0.0001 | 0.0175 | <0.0001 | 38691 |
| Townsend | - | - | - | - | - | - | - | - | 0.1129 | 0.0054 | <0.0001 | 38691 |
| BMI | - | - | - | - | - | - | - | - | 0.0546 | 0.0051 | <0.0001 | 38691 |
| Intercept variance | 0.4997 | 0.7069 | - | - | 0.4966 | 0.7047 | - | - | 0.4796 | 0.6926 | - | - |
| Age (slope) variance | 0.0011 | 0.0331 | - | - | 0.0011 | 0.0331 | - | - | 0.0011 | 0.0326 | - | - |
| Intercept/age covariance | -0.0024 | -0.1006 | - | - | -0.0024 | -0.1052 | - | - | -0.0019 | -0.0843 | - | - |
| Residual variance | 0.582 | 0.7629 | - | - | 0.5824 | 0.7631 | - | - | 0.5816 | 0.7626 | - | - |
| Deviance | 277432.7901 | | | | 277343.2828 | | | | 275312.5407 | | | |
| AIC | 277512.7901 | | | | 277425.2828 | | | | 275402.5407 | | | |
| BIC | 277893.2206 | | | | 277815.2241 | | | | 275830.3696 | | | |

Supplementary Table 25. Model estimates for sensitivity analysis in UK Biobank. Individuals that remained alive after initial baseline appointment only.

| | Unadjusted | | | | Sex Adjusted | | | | Fully Adjusted | | | |
|---|---|---|---|---|---|---|---|---|---|---|---|---|
| Parameter | Estimate | SE | p-value | N | Estimate | SE | p-value | N | Estimate | SE | p-value | N |
| Intercept | 0.6219 | 0.1395 | <0.0001 | 36798 | 0.5653 | 0.1394 | <0.0001 | 36798 | 0.7435 | 0.1389 | <0.0001 | 36515 |
| age | <0.0001 | 0.0007 | <0.0001 | 36798 | <0.0001 | 0.0007 | <0.0001 | 36798 | <0.0001 | 0.0007 | <0.0001 | 36515 |
| age^2 (acceleration) | <0.0001 | <0.0001 | 0.1169 | 36798 | <0.0001 | <0.0001 | 0.0982 | 36798 | <0.0001 | <0.0001 | 0.1158 | 36515 |
| Batch1 | 0.0102 | 0.1396 | 0.9415 | 36798 | 0.0142 | 0.1394 | 0.919 | 36798 | <0.0001 | 0.1374 | 0.8082 | 36515 |
| Batch2 | <0.0001 | 0.1377 | 0.9497 | 36798 | <0.0001 | 0.1375 | 0.9702 | 36798 | <0.0001 | 0.1356 | 0.7014 | 36515 |
| Batch3 | 0.0208 | 0.1373 | 0.8794 | 36798 | 0.0244 | 0.1371 | 0.8589 | 36798 | <0.0001 | 0.1352 | 0.8861 | 36515 |
| Batch4 | 0.0521 | 0.1375 | 0.7045 | 36798 | 0.0553 | 0.1373 | 0.6869 | 36798 | 0.0089 | 0.1354 | 0.9476 | 36515 |
| Batch5 | 0.008 | 0.1375 | 0.9537 | 36798 | 0.0122 | 0.1373 | 0.9291 | 36798 | <0.0001 | 0.1354 | 0.7687 | 36515 |
| Batch6 | 0.0209 | 0.1375 | 0.879 | 36798 | 0.0246 | 0.1373 | 0.858 | 36798 | <0.0001 | 0.1354 | 0.8894 | 36515 |
| Batch7 | 0.0092 | 0.1377 | 0.947 | 36798 | 0.0127 | 0.1375 | 0.9267 | 36798 | <0.0001 | 0.1356 | 0.791 | 36515 |
| Assessment_centre_baseline Birmingham | <0.0001 | 0.0352 | 0.3531 | 36798 | <0.0001 | 0.0351 | 0.4153 | 36798 | 0.1491 | 0.0359 | <0.0001 | 36515 |
| Assessment_centre_baseline Bristol | <0.0001 | 0.0323 | <0.0001 | 36798 | <0.0001 | 0.0323 | <0.0001 | 36798 | 0.0621 | 0.034 | 0.0679 | 36515 |
| Assessment_centre_baseline Bury | <0.0001 | 0.0338 | <0.0001 | 36798 | <0.0001 | 0.0338 | <0.0001 | 36798 | 0.0466 | 0.0353 | 0.1864 | 36515 |
| Assessment_centre_baseline Cardiff | <0.0001 | 0.037 | 0.1603 | 36798 | <0.0001 | 0.0369 | 0.1785 | 36798 | 0.1765 | 0.0384 | <0.0001 | 36515 |
| Assessment_centre_baseline Croydon | <0.0001 | 0.0344 | 0.0721 | 36798 | <0.0001 | 0.0343 | 0.0722 | 36798 | 0.1188 | 0.035 | 0.0007 | 36515 |
| Assessment_centre_baseline Edinburgh | <0.0001 | 0.0378 | <0.0001 | 36798 | <0.0001 | 0.0378 | <0.0001 | 36798 | 0.0408 | 0.0391 | 0.2966 | 36515 |
| Assessment_centre_baseline Glasgow | <0.0001 | 0.0374 | 0.0099 | 36798 | <0.0001 | 0.0374 | 0.0103 | 36798 | 0.0807 | 0.0381 | 0.034 | 36515 |
| Assessment_centre_baseline Hounslow | <0.0001 | 0.0338 | 0.0003 | 36798 | <0.0001 | 0.0338 | 0.0003 | 36798 | 0.0528 | 0.0344 | 0.1245 | 36515 |
| Assessment_centre_baseline Leeds | <0.0001 | 0.0315 | <0.0001 | 36798 | <0.0001 | 0.0314 | <0.0001 | 36798 | 0.0494 | 0.033 | 0.1345 | 36515 |

| | Unadjusted | | | | Sex Adjusted | | | | Fully Adjusted | | | |
|---|---|---|---|---|---|---|---|---|---|---|---|---|
| Parameter | Estimate | SE | p-value | N | Estimate | SE | p-value | N | Estimate | SE | p-value | N |
| Assessment_centre_baseline Liverpool | <0.0001 | 0.0342 | 0.0002 | 36798 | <0.0001 | 0.0341 | 0.0003 | 36798 | 0.0868 | 0.0353 | 0.014 | 36515 |
| Assessment_centre_baseline Manchester | <0.0001 | 0.0389 | 0.2393 | 36798 | <0.0001 | 0.0389 | 0.2761 | 36798 | 0.1472 | 0.0396 | 0.0002 | 36515 |
| Assessment_centre_baseline Middlesborough | <0.0001 | 0.0354 | <0.0001 | 36798 | <0.0001 | 0.0354 | <0.0001 | 36798 | 0.0847 | 0.0366 | 0.0207 | 36515 |
| Assessment_centre_baseline Newcastle | <0.0001 | 0.033 | <0.0001 | 36798 | <0.0001 | 0.033 | <0.0001 | 36798 | 0.0552 | 0.0341 | 0.1058 | 36515 |
| Assessment_centre_baseline Nottingham | <0.0001 | 0.0323 | <0.0001 | 36798 | <0.0001 | 0.0322 | <0.0001 | 36798 | 0.1139 | 0.0339 | 0.0008 | 36515 |
| Assessment_centre_baseline Oxford | <0.0001 | 0.0405 | <0.0001 | 36798 | <0.0001 | 0.0404 | <0.0001 | 36798 | 0.0574 | 0.0416 | 0.1676 | 36515 |
| Assessment_centre_baseline Reading | <0.0001 | 0.0345 | <0.0001 | 36798 | <0.0001 | 0.0345 | <0.0001 | 36798 | 0.0612 | 0.0365 | 0.0934 | 36515 |
| Assessment_centre_baseline Sheffield | <0.0001 | 0.0334 | 0.0001 | 36798 | <0.0001 | 0.0334 | 0.0001 | 36798 | 0.0943 | 0.0346 | 0.0065 | 36515 |
| Assessment_centre_baseline Stoke | <0.0001 | 0.0373 | 0.0003 | 36798 | <0.0001 | 0.0373 | 0.0006 | 36798 | 0.1216 | 0.0388 | 0.0017 | 36515 |
| Assessment_centre_baseline Swansea | <0.0001 | 0.098 | 0.8877 | 36798 | <0.0001 | 0.0978 | 0.9047 | 36798 | 0.1547 | 0.097 | 0.1106 | 36515 |
| Assessment_centre_baseline Wrexham | <0.0001 | 0.1628 | 0.9694 | 36798 | <0.0001 | 0.1626 | 0.9865 | 36798 | 0.2041 | 0.1608 | 0.2044 | 36515 |
| IL6_tertileMiddle | 0.0186 | 0.0128 | 0.1469 | 36798 | 0.0196 | 0.0128 | 0.1255 | 36798 | <0.0001 | 0.0129 | 0.0823 | 36515 |
| IL6_tertileTop | 0.092 | 0.0135 | <0.0001 | 36798 | 0.0931 | 0.0135 | <0.0001 | 36798 | <0.0001 | 0.0143 | 0.9227 | 36515 |
| age:IL6_tertileMiddle | <0.0001 | 0.001 | 0.1095 | 36798 | <0.0001 | 0.001 | 0.1168 | 36798 | <0.0001 | 0.001 | 0.2756 | 36515 |
| age:IL6_tertileTop | <0.0001 | 0.0011 | 0.4309 | 36798 | <0.0001 | 0.0011 | 0.4832 | 36798 | 0.0002 | 0.0011 | 0.8794 | 36515 |
| age^2 (acceleration):IL6_tertileMiddle | 0.0002 | <0.0001 | 0.0009 | 36798 | 0.0002 | <0.0001 | 0.0008 | 36798 | 0.0002 | <0.0001 | 0.002 | 36515 |
| age^2 (acceleration):IL6_tertileTop | 0.0003 | <0.0001 | <0.0001 | 36798 | 0.0003 | <0.0001 | <0.0001 | 36798 | 0.0003 | <0.0001 | <0.0001 | 36515 |

| | Unadjusted | | | | Sex Adjusted | | | | Fully Adjusted | | | |
|---|---|---|---|---|---|---|---|---|---|---|---|---|
| Parameter | Estimate | SE | p-value | N | Estimate | SE | p-value | N | Estimate | SE | p-value | N |
| SexFemale | - | - | - | - | 0.0901 | 0.0098 | <0.0001 | 36798 | 0.1178 | 0.0099 | <0.0001 | 36515 |
| smoking_statusNever | - | - | - | - | - | - | - | - | <0.0001 | 0.0177 | <0.0001 | 36515 |
| smoking_statusPrevious | - | - | - | - | - | - | - | - | <0.0001 | 0.0184 | <0.0001 | 36515 |
| Townsend | - | - | - | - | - | - | - | - | 0.1146 | 0.0056 | <0.0001 | 36515 |
| BMI | - | - | - | - | - | - | - | - | 0.0672 | 0.0054 | <0.0001 | 36515 |
| Intercept variance | 0.5116 | 0.7153 | - | - | 0.5093 | 0.7136 | - | - | 0.4886 | 0.699 | - | - |
| Age (slope) variance | 0.0011 | 0.0327 | - | - | 0.0011 | 0.0326 | - | - | 0.001 | 0.0322 | - | - |
| Intercept/age covariance | -0.002 | -0.0863 | - | - | -0.0021 | -0.0906 | - | - | -0.0016 | -0.0706 | - | - |
| Residual variance | 0.5858 | 0.7654 | - | - | 0.586 | 0.7655 | - | - | 0.585 | 0.7648 | - | - |
| Deviance | 272455.4412 | | | | 272371.9245 | | | | 269594.9976 | | | |
| AIC | 272535.4412 | | | | 272453.9245 | | | | 269684.9976 | | | |
| BIC | 272915.086 | | | | 272843.0604 | | | | 270111.8392 | | | |

Supplementary Table 26. Model estimates from linear regression of number of PHQ-2 questionnaires participants completed on IL-6 tertile in UK Biobank. Participants subset to those that remained alive after the initial assessment. Excluding the two imaging appointments as only a subset of participants were invited to these appointments. Total number of time points were 6.

| term | estimate | SE | statistic | p.value |
|---|---|---|---|---|
| (Intercept) | 3.287 | 0.011 | 298.352 | p<0.0001 |
| IL6_tertileMiddle | -0.140 | 0.016 | -8.891 | p<0.0001 |
| IL6_tertileTop | -0.281 | 0.016 | -17.093 | p<0.0001 |

**Supplementary Material References**
